# Vitamin D Receptor Gene Polymorphisms in Type 1, Type 2, and Gestational Diabetes Mellitus: A Comprehensive Meta-Analysis and Meta-Regression of 154 Studies

**DOI:** 10.1101/2025.02.20.25322588

**Authors:** Haider Ali Alnaji, Al-Karrar Kais Abdul Jaleel, Muslimbek G. Normatov, Ali Abbas Abo Algon, Hanaa Addai Ali, Abbas F. Almulla

## Abstract

**Background:** Diabetes mellitus (DM) includes metabolic disorders marked by chronic hyperglycemia. Vitamin D and its receptor (VDR) play crucial roles in DM pathophysiology. Four single nucleotide polymorphisms (SNPs) in the VDR gene, namely FokI, TaqI, BsmI, and ApaI, have been implicated in DM risk. However, no prior meta-analysis has systematically assessed their associations across type 1 diabetes mellitus (T1DM), type 2 diabetes mellitus (T2DM), and gestational diabetes mellitus (GDM).

**Objectives:** To investigate the association of FokI, TaqI, BsmI, and ApaI polymorphisms with susceptibility to T1DM, T2DM, and GDM.

**Methods:** A systematic search of PubMed, Google Scholar, and SciFinder identified 154 studies (49,675 participants: 23,225 DM patients and 26,450 controls). Meta-analyses assessed genetic associations, and subgroup analyses were performed by ethnicity and DM subtype.

**Results:** Significant associations were observed between T1DM and FokI, BsmI, and ApaI polymorphisms, while TaqI showed no association. For T2DM, FokI, BsmI, and TaqI polymorphisms were associated with risk in specific ethnic groups. GDM analysis revealed no overall associations, though the FokI SNP showed significance in one ethnic subgroup. Comparative analysis across DM types revealed no differences in VDR polymorphisms except for the BsmI SNP, which increased T2DM risk in certain genetic models.

**Conclusion:** The G allele of the BsmI SNP significantly increases T2DM risk, while the T allele of the FokI SNP is protective in T1DM. These findings highlight the importance of VDR polymorphisms in DM susceptibility across diverse populations.

## Introduction

Diabetes mellitus (DM) encompasses a spectrum of metabolic disorders primarily defined by persistent hyperglycemia, which increases the risk for severe complications, heightened healthcare costs, diminished quality of life, and elevated mortality (Cho et al., 2018; Panahi et al., 2024). The major forms of DM include type 1 diabetes mellitus (T1DM), type 2 diabetes mellitus (T2DM), and gestational diabetes mellitus (GDM) (Ahmad, Lim, Lamptey, Webb, & Davies, 2022; McIntyre et al., 2019; Redondo & Morgan, 2023). The global burden of diabetes has surged, with approximately 415 million individuals affected in 2015, rising to 537 million by 2021 (Atlas, 2015; Magliano, Boyko, & Atlas, 2021). Projections from the International Diabetes Federation’s 10th edition suggest an alarming increase to 643 million by 2030 and 783 million by 2045 (Kumar, Gangwar, Zargar, Kumar, & Sharma, 2024). T2DM accounts for over 90% of all DM cases, imposing a considerable economic strain on global healthcare systems (Jia Liu, Zhang, Shi, He, & Xia, 2023), while T1DM constitutes about 5–10% of cases. The rise in diabetes prevalence aligns with accelerated economic growth, urbanization, and the adoption of modern lifestyles.

The underlying mechanisms contributing to diabetes differ by type. T1DM results from autoimmune destruction of insulin-producing β-cells, leading to an absolute insulin deficiency. This process involves immune-mediated β-cell damage exacerbated by stress-induced dysfunction, triggering further immune responses (Galicia-Garcia et al., 2020; Roep, Thomaidou, van Tienhoven, & Zaldumbide, 2021). In contrast, T2DM is characterized by a complex interplay of insulin resistance and β-cell dysfunction. Peripheral tissues such as muscle, liver, and adipose tissue become resistant to insulin, driven by factors including obesity, chronic inflammation, and oxidative stress. As insulin resistance advances, β-cell performance declines, contributing to insufficient insulin secretion and hyperglycemia (Ortega-Contreras et al., 2022). GDM, characterized by glucose intolerance manifesting during pregnancy, is influenced by genetic and environmental factors, compounded by pregnancy-related metabolic and hormonal changes.

Vitamin D has been implicated in the pathogenesis of diabetes, supported by evidence linking vitamin D deficiency to increased diabetes risk (Lips et al., 2017; Sergeev, 2016; Wacker & Holick, 2013). The active form of vitamin D exerts its physiological effects through the vitamin D receptor (VDR), expressed in various tissues, including the pancreas (Y. Wang, Zhu, & DeLuca, 2012). In pancreatic β-cells, vitamin D, via nuclear VDR (nVDR), influences insulin synthesis through interaction with vitamin D response elements (VDREs) in insulin gene promoters (Maestro, Dávila, Carranza, & Calle, 2003). This regulation may enhance insulin production, protect β-cells, and reduce muscle insulin resistance (Cândido & Bressan, 2014).

Polymorphisms in the VDR gene may affect its function, influencing the risk of diabetes (Oh & Barrett-Connor, 2002). Over 25 polymorphisms have been identified, with some evidence supporting their association with T1DM and T2DM development (Palomer, González Clemente, Blanco Vaca, & Mauricio, 2008; Penna-Martinez & Badenhoop, 2017; Qin et al., 2014). These genetic variations may alter immune function and calcium metabolism, potentially contributing to β-cell autoimmunity in T1DM and impairing insulin secretion in T2DM (Lemire, 2000; Mathieu et al., 2004). The role of VDR polymorphisms in GDM has also been explored, though results remain varied (Mao, Li, & Gao, 2012). Key VDR polymorphisms such as BsmI, ApaI, TaqI, and FokI have been studied for their potential role in diabetes susceptibility and related metabolic effects (Z. Liu, Liu, Chen, He, & Yu, 2014; Rahmannezhad, Mashayekhi, Goodarzi, Rezvanfar, & Sadeghi, 2016).

Previous meta-analyses have provided mixed conclusions regarding the significance of VDR polymorphisms in diabetes. For instance, Zeng et al. (2022) found that the VDR rs739837 polymorphism is significantly associated with T2DM risk but not GDM (Zeng et al., 2022). Wang and Xue (2020) reported that the FokI rs2228570 polymorphism increases susceptibility to GDM, particularly in South Asian populations (B. Wang & Xue, 2020). Shahmoradi et al. (2021) highlighted a protective effect of the BsmI SNP in T1DM under specific genetic models (Shahmoradi, Ghaderi, Aghaei, & Azarnezhad, 2021). However, some meta-analyses, such as Zhai et al. (2020) and Tizaoui et al. (2014), emphasize ethnic-specific associations and interactions with environmental factors rather than significant global associations (Tizaoui, Kaabachi, Hamzaoui, & Hamzaoui, 2014; Zhai et al., 2020b). These studies did not perform a meta-regression to examine the impact of some factors on the heterogeneity in the results of VDR gene polymorphism.

Given these discrepancies and lack of examination of various phenotypic factors that may influence the appearance of these SNPs, this comprehensive meta-analysis and meta-regression aim to synthesize existing evidence to clarify the role of VDR polymorphisms namely FokI, TaqI, BsmI and ApaI in the susceptibility to T1DM, T2DM, and GDM. This study will examine five genetic models (allelic, recessive, dominant, overdominant, homozygous, and heterozygous) to provide a more nuanced understanding of these associations.

## Materials and method

In this study, we adhered to several key methodological frameworks, including PRISMA 2020 guidelines, the Cochrane Handbook for Systematic Reviews of Interventions, and the Meta-Analyses of Observational Studies in Epidemiology (MOOSE) guidelines. Our analysis focused on patients with different types of DM (T1DM, T2DM, and GDM) and healthy controls, examining gene polymorphism in various SNPs including FokI (rs2228570), TaqI (rs731236), BsmI (rs1544410), and ApaI (rs7975232).

### Search strategy

To obtain comprehensive data on the FokI (rs2228570), TaqI (rs731236), BsmI (rs1544410), and ApaI (rs7975232) polymorphisms in T1DM, T2DM, and GDM, we performed a systematic search across multiple electronic databases, including PubMed/MEDLINE, Google Scholar, and SCOPUS. The search spanned from June 15 to the end of September 2024, utilizing predefined keywords and MeSH terms (refer to S1, Table 1). To ensure comprehensive coverage, we also examined the reference lists of relevant studies and prior meta-analyses to identify any potentially overlooked significant research.

### Eligibility criteria

We carried out a thorough search of relevant studies, giving priority to peer-reviewed articles published in English. To broaden the scope of data collection, we also included grey literature and research papers in China, and Arabic languages. Eligible studies were those with an observational design, either case-control or cohort, that incorporated control groups and examined at least one polymorphism of the vitamin D receptor (VDR) gene, such as FokI (rs2228570), TaqI (rs731236), BsmI (rs1544410), or ApaI (rs7975232), in individuals with T1DM, T2DM, or GDM, as diagnosed based on the World Health Organization (WHO) criteria (1999) (Organization, 1999). Studies also had to provide sufficient data to calculate odds ratios (OR) and 95% confidence intervals (CIs) and report detailed genotype frequencies, along with the total number of cases and controls. Studies were excluded if they were reviews, book chapters, duplicates, any study didn’t mention sufficient data or otherwise deemed irrelevant.

### Screening and data extraction

The initial phase of this meta-analysis involved two researchers, HA and AQ, who independently screened study titles and abstracts against the predefined inclusion criteria. After this preliminary review, the full texts of studies deemed potentially relevant were retrieved for further evaluation, while those not meeting the exclusion criteria were discarded. HA, AQ, and AA then systematically extracted key information into a custom Excel sheet, which included details such as the authors, publication dates, SNP frequency data, the number of participants in both patient and control groups, and the overall sample size for each study. Additional recorded variables included the study design, types of biological samples used, key biomarkers for DM (such as HbA1c and fasting or random blood glucose), as well as demographic data like participant age, gender, ethnicity, latitude and geographical location. Any inconsistencies in data extraction or interpretation were resolved by consulting with the senior author, AA. The included studies were assessed for methodological quality using the Newcastle-Ottawa Scale (NOS) following the guidelines of (Stang, 2010). This tool evaluates studies in three categories: Selection, Comparability, and Exposure, with a total of nine criteria. Based on their scores, studies were classified as low (0–3 stars), moderate (4–6 stars), or high quality (7–9 stars), as shown in S1 Table 2 of the supplementary file 1.

### Data analysis

In this meta-analysis, we utilized SPSS version 30, adhering to PRISMA guidelines (see ESF-1, Table 5). For each single nucleotide polymorphism (SNP) examined, at least two studies were required for inclusion. The Hardy-Weinberg equilibrium (HWE) in the control groups was assessed using the Chi-squared (χ²) test. The associations between VDR gene polymorphisms and various types of DM (T1DM, T2DM, and GDM) were analyzed using OR and their respective 95% CI.

The relationships between VDR gene polymorphisms FokI, TaqI, BsmI, and ApaI and the three types of diabetes (T1DM, T2DM and GDM) were explored through various genetic models. The specified models for FokI, TaqI, BsmI, and ApaI SNPs are included the allelic, recessive, dominant, overdominant, homozygous and heterozygous. The symbols for each model are as follows: For FokI; the allelic (T vs. C), Recessive model (TT vs. TC + CC), Dominant model (TT+TC vs. CC), Overdominant model (TC vs. TT+CC), homozygous (TT vs. CC) and heterozygous (TC vs. CC), in addition to the (TT vs. TC) model. For TaqI; the Allelic (C vs. T), Recessive model (CC vs. CT+TT), Dominant model (CC+CT vs. TT), Overdominant model (CT vs. CC+TT), homozygous (CC vs. TT), heterozygous (CT vs. TT), in addition to (CC vs. CT) model. For BsmI; Allelic (G vs. A), Recessive model (GG vs. GA+AA), Dominant model (GG+GA vs. AA), Overdominant model (GA vs. GG+AA), homozygous (GG vs. AA), heterozygous (GA vs. AA), in addition to (GG vs. GA) model. The last models is for ApaI is as follows: Allelic (G vs. T), Recessive model (GG vs. GT+TT), Dominant model (GG+GT vs. TT), Overdominant model (GT vs. GG+TT), homozygous (GG vs. TT), heterozygous (GT vs. TT), in addition to the (GG vs. GT) model.

We used Cochran’s Q-statistic (p < 0.10 indicating statistical significance) and the I-squared (I²) test to assess heterogeneity across studies (Huedo-Medina, Sánchez-Meca, Marín-Martínez, & Botella, 2006). A fixed-effects model (FEM) was applied when the Q-statistic p-value exceeded 0.10 or I² was below 50%, indicating low heterogeneity. Otherwise, a random-effects model (REM) was used (DerSimonian & Laird, 1986; Mantel & Haenszel, 1959). Subgroup analyses and meta-regressions were conducted based on variables such as study year, ethnicity, age, HbA1c, duration of illness, BMI and sample size to explore sources of heterogeneity.

Furthermore, we performed sensitivity analyses by sequentially excluding individual studies to assess the stability of the pooled estimates. This allowed us to determine whether any specific study disproportionately affected the results. To examine publication bias and the impact of small-study effects, we employed Begg’s funnel plots and Egger’s regression test, considering p-values less than 0.05 as statistically significant (Begg & Mazumdar, 1994; Egger, Smith, Schneider, & Minder, 1997).

## Results

### Study characteristics

In our study, we utilized specific keywords and MeSH terms (refer to ESF-1, Table 2) to conduct a systematic search across several literature databases, including PubMed, Google Scholar, and SciFinder. This initial search retrieved a total of 5331 studies, as illustrated in the PRISMA flow diagram (**Figure 1**), which outlines the process of study selection and exclusion. After an exhaustive screening to remove irrelevant and duplicate records, we refined our selection to 174 articles. Of these, only 154 studies met the stringent inclusion criteria for our systematic review. Ultimately, a total of 154 studies were included in the final meta-analysis, adhering to our established inclusion and exclusion criteria. However, two of the included studies featured cohorts of both T1DM and T2DM patients within the same study, so each was treated as two separate studies: one for T1DM and one for T2DM. Additionally, one study included two distinct T1DM cohorts from different cities, which were also considered as two separate studies. Therefore, the meta-analysis incorporated data from 157 studies, including 57 on T1DM, 85 on T2DM, and 15 on GDM, all examining various models of VDR gene polymorphisms such as FokI (rs2228570), TaqI (rs731236), BsmI (rs1544410), and ApaI (rs7975232). Overall, 49,675 participants were included: 26,450 healthy controls and 23,225 patients with different types of diabetes (T1DM: 8,013, T2DM: 12,206, GDM: 2,306).

**Figure 1.**
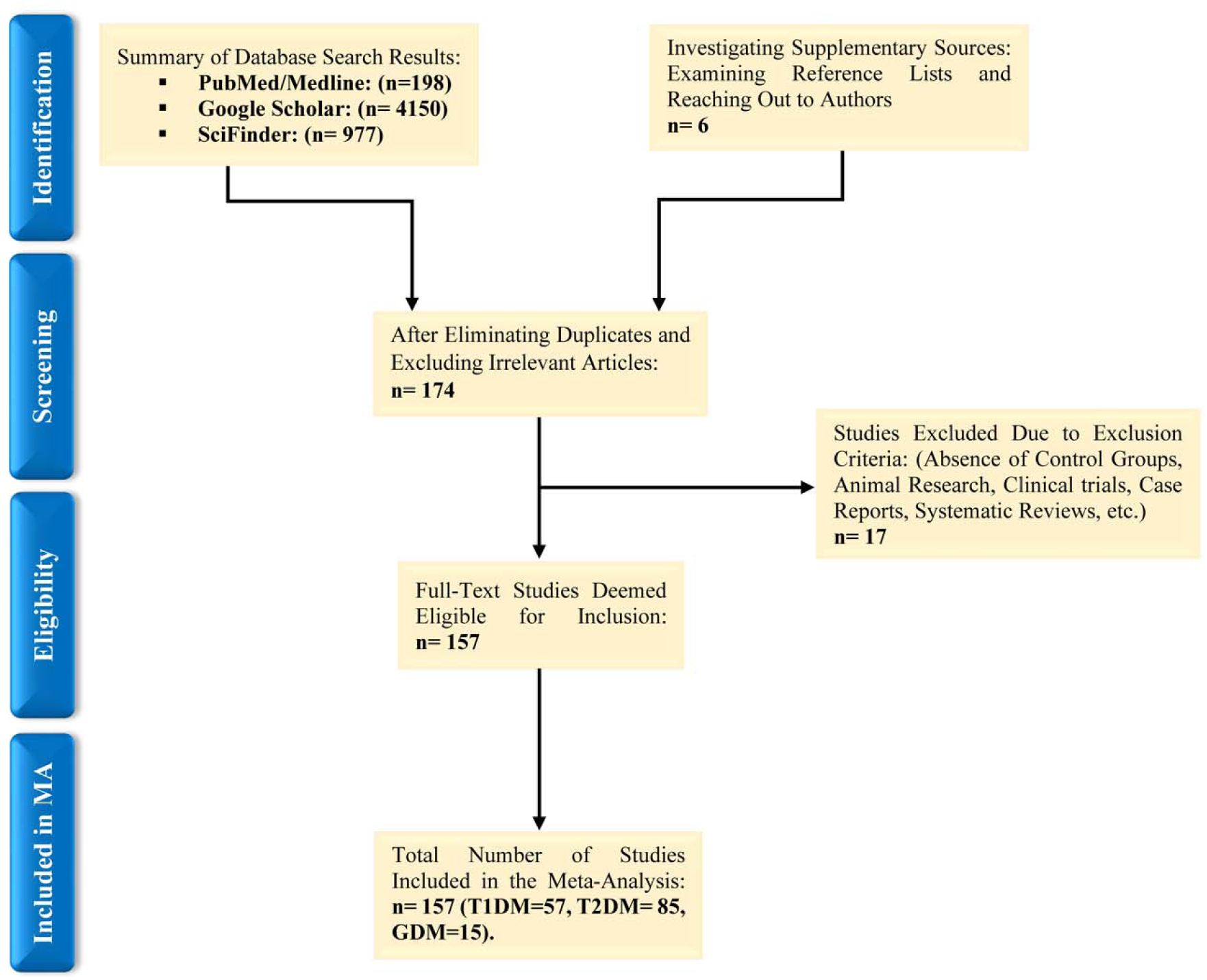
The PRISMA flow diagram.

Participants ranged in age from 5.8 to 80 years (Abd-Allah, Pasha, Hagrass, & Alghobashy, 2014; ABD EL-SALAM, GABER, & EMAM; AbdulKhaliq¹, Farhan, & AlKhateeb, 2020; A. E.-A. Ahmed, Sakhr, Hassan, El-Amir, & Ameen, 2019; S. H. Ahmed, Foad, Ahmed, Mohamed, & Sabry, 2021; Al-Daghri et al., 2012; S. Al-Darraji, H. Al-Azzawie, & A. Al-Kharsani, 2017; Al-Kashwan, Algenabi, Omara, & Kaftan, 2021; Al-Mawla, Ali, & Al-Ani, 2020; Al-Moubarak & Haddad, 2013; AL-TIMIMI & MPHIL; Al Safar et al., 2018; Alfaqih et al., 2022; Ali Khan, Alhaizan, Neyazi, Al-Hakeem, & Alshammary, 2023; Ali, Fawzy, Mohsen, & Settin, 2018; Alkhedaide et al., 2021; Alzaim et al., 2024; Angel, Lera, Márquez, & Albala, 2018; Angel, Lera, Sánchez, Oyarzún, & Albala, 2016; Apaydın et al., 2019a; Aslani et al., 2011; Ata, 2024; Audi et al., 2004; Azizi, Behzadi Andohjerdi, & Mohajerani, 2020; Ban et al., 2001; Batool, Mukhtar, Qayyum, & Shakeel, 2016; Berca et al., 2021; Bertoccini et al., 2017; Beysel et al., 2019; Bid et al., 2009; Bonakdaran, Abbaszadegan, Dadkhah, & Dalouie, 2012; Boullu-Sanchis et al., 1999; Capoluongo et al., 2006; Chang et al., 2000; Chen et al., 2022; Cheon et al., 2015; Denisova, Ivaschenko, Yarmolinskaya, Shagina, & Misharina, 2021; Dilmec, Uzer, Akkafa, Kose, & van Kuilenburg, 2010; Dong et al., 2018; Durgawale et al., 2022; Eissa et al., 2021; El-Beshbishy et al., 1969; El-Kafoury, Haroun, Embaby, & Dawoods, 2014; El Gendy, Sadik, Helmy, & Rashed, 2019; Elfasakhany, Alqahtani, Elguindy, & El-Damarawi, 2022; Elgazzaz, Mohammed, Atwa, & Badr, 2016; Elhuseiny, Ezz, Mohamed, & Hassan, 2024; Errouagui et al., 2014; Eweida et al., 2021; Ezhilarasi, Dhamodharan, & Vijay, 2018; Fassbender et al., 2002; Fatma & Abdul, 2019; Ferraz et al.; García et al., 2007; GEHAN et al., 2018; Gnanaprakash et al., 2019; Gogas Yavuz et al., 2011; Gomaa, Shehata, Ooda, & Eldeeb, 2022; Györffy et al., 2002; Hadi & Al-Zubaidy, 2019; Hamed, Abdel-Aal, Din, & Atia, 2013; Hatmal et al., 2020; Hauache et al., 1998; Hesham A et al., 2015; Israni, Goswami, Kumar, & Rani, 2009; Iyer, New, Khoja, Al-Ghamdi, & Bahlas, 2017; Jia et al., 2015; Kaftan, Hussain, Algenabi, Omara, & Al-Kashwan, 2021; Kandala & Abdul Ridha, 2016; Khan et al., 2019; Khdair, Jarrar, & Jarrar, 2021; Kirac et al., 2018; Klahold et al., 2020; Kocabaş, Karagüzel, İmir, Yavuzer, & Akcurin, 2010; Konečná et al., 2023; Lemos et al., 2008; Jianqiong Liu et al., 2021; López et al., 2008; Ma et al., 2020; Mackawy & Badawi, 2014; Mahjoubi et al., 2016; Maia et al., 2016; Malecki et al., 2003; Malik et al., 2018; Maulood, 2021; Memon, Baig, & Siddiqui, 2022; Miettinen et al., 2015; Mohamed Abd El-Maksoud, Abulsoud, Abulsoud, & Elshaer, 2022; Mohamed, Shipl, Sarhan, Sakar, & Arfa, 2020; Mohammadnejad et al., 2012; A. A. Mohammed et al., 2023; Mohammed, Jaff, & Aziz, 2023; Mohyi, El-Abd, Rashed, Omar, & A AbdulLatif, 2022; Morán-Auth, Penna-Martinez, & Badenhoop, 2015; Mory et al., 2009; Mostafa et al., 2024; Motohashi et al., 2003; Mukhopadhyaya, Acharya, Chavan, Purohit, & Mutha, 2010; Mukhtar, Batool, Wajid, & Qayyum, 2017; Mutar, Allwsh, Yassin, Al-Tu’ma, & Taghi, 2023; Najjar, Jwad, & Alkhafaji, 2020; Nasreen, Lone, Khaliq, & Khaliq, 2016; Reza Nosratabadi, Arababadi, & Salehabad, 2011; R. Nosratabadi et al., 2010; Oh & Barrett-Connor, 2002; Galawezh Obaid Othman, 2022; GALAWEZH O OTHMAN, AALI, SAEED, & NAJEEB, 2023; Panierakis et al., 2009; Paul, 2010; Pedro et al., 2005; Pinho et al., 2019; Radwan, Taha, ElSayed, & Omar, 2021; Rahmannezhad et al., 2016; Rasheed et al., 2017; Rasoul, Haider, Al-Mahdi, Al-Kandari, & Dhaunsi, 2019; Raya Hatem, Ali, & Al-Ani, 2020; RAYIMOVA et al., 2024; Razi et al., 2019; Ridha & Kandala, 2016; Rivera-Leon et al., 2015; Rodrigues et al., 2019; Sah et al., 2024; Sahin, Cetinkalp, Erdogan, Yilmaz, & Berdeli, 2012; Salehizadeh et al., 2024; Sarma, Chauhan, Saikia, Sarma, & Nath, 2018; Sattar & Hussain, 2020; Sattar, Shaheen, Hussain, & Jamil, 2021; Sattar, Subhani, Jabeen, Yaqub, & Akhtar, 2020; Saxena et al., 2020; Selvarajan, Srinivasan, Surendran, Mathaiyan, & Kamalanathan, 2021; Shafie et al., 2022; Sharkawy, Abdel-Ghaffar, Gharib, & El; Shimada et al., 2008; Siqueira, Araujo, Mattar, & Daher, 2019; Škrabić, Zemunik, Šitum, & Terzić, 2003; TANEJA, KHADAGAWAT, & MANI, 2017; Taneja, Khadgawat, & Mani, 2016; Tangjittipokin et al., 2021; A. R. Tawfeek, Tawfeek, Aboelyazeid Ellayen, & Fathy, 2018; M. Tawfeek, Habib, & Saultan, 2011; Thirunavukkarasu, Chitra, Asirvatham, & Jayalakshmi, 2024; Turpeinen et al., 2003; Vedralová et al., 2013; Vural & Maltas, 2012; Xi et al., 2017; Xia et al., 2017; Yavuz et al., 2022; Yildiz, Üstündağ, Tiber, & Doğan, 2022; Youssef, El-Dosoky, El-Shafey, & Abed, 2019; Zakaria et al., 2021; Zemunik et al., 2005; Zhang et al., 2012; Zhong et al., 2015; Zhu et al., 2019). SNP frequencies in the VDR gene were primarily determined using PCR-Restriction Fragment Length Polymorphism (RFLP) and TaqMan techniques, while other methods were less frequently used, as detailed in ESF-1, Table 5. Egypt led in contributions with 26 studies, followed by Iraq (16), and India (13). China, Turkey, and Saudi Arabia each contributed 10 studies, while Iran (9), Pakistan (8), Brazil (7), and Chile (4) also made notable contributions. Spain, Germany, Japan, and Jordan each conducted 3 studies, with Romania, Croatia, Italy, the Czech Republic, and Finland contributing 2 studies each. Various countries including Tunisia, Guadeloupe, England, the USA, and others contributed one study each. A full list is available in S2, Tables 1,2, and 3. The quality of these studies was evaluated using the Newcastle-Ottawa Scale (NOS)(Wells et al., 2000), yielding scores that ranged from 6 to 9 (Median = 8). Study characteristics and genotype frequencies are summarized in S2, Tables 1, 2, and 3.

*The results of FokI (rs2228570) polymorphism in association with T1DM, T2DM and GDM*.

#### T1DM

An extensive analysis of the FokI gene SNP was performed, incorporating data from 38 studies in total. These included 13 studies from European countries, 17 from Asian countries, 5 from African nations, and 3 from American countries, as presented in **Table 1**. The pooled results demonstrated a significant association between FokI polymorphism and T1DM under two genotype models. Specifically, the allelic model (T vs. C) yielded an OR of 0.81 (95% CI = 0.67–0.98, p = 0.029) as shown in **Figure 2**, and the heterozygous model (TC vs. CC) showed an OR of 0.83 (95% CI = 0.71–0.97, p = 0.019, see **Figure 3**) suggesting the association of FokI SNP with decreased risk of T1DM in these models. Egger’s test, outlined in Table 1, showed no evidence of significant bias across these models. Additionally, ethnicity-based subgroup analysis revealed a reduced susceptibility to T1DM among the Asian population, particularly in the heterozygous (TC vs. CC) model (see **Figure 3**) and TT vs. TC model (S1, Figure 1). No significant association was detected for others population.

**Figure 2.**
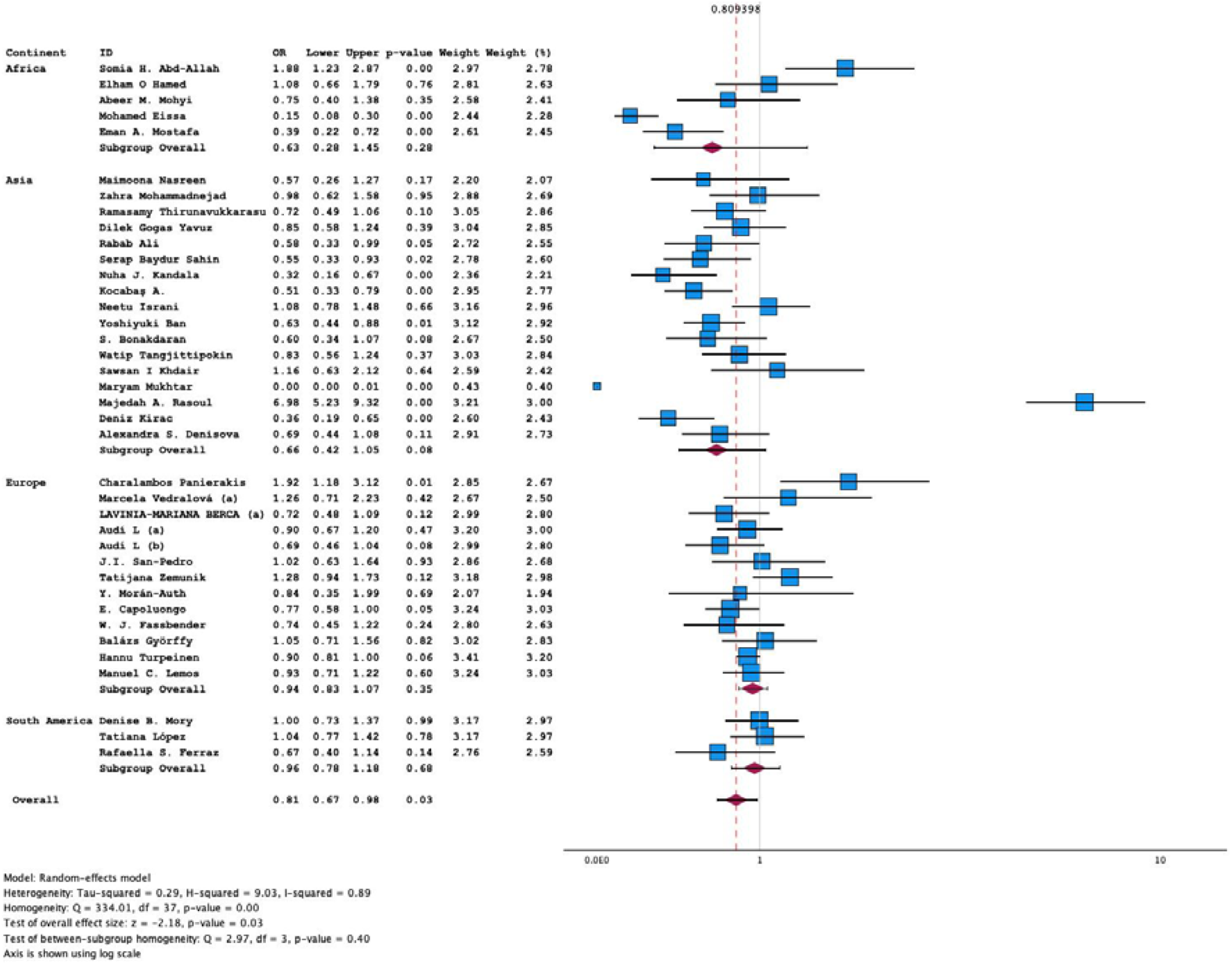
Forest plot of FokI SNP allelic (T vs C) model in T1DM

**Figure 3.**
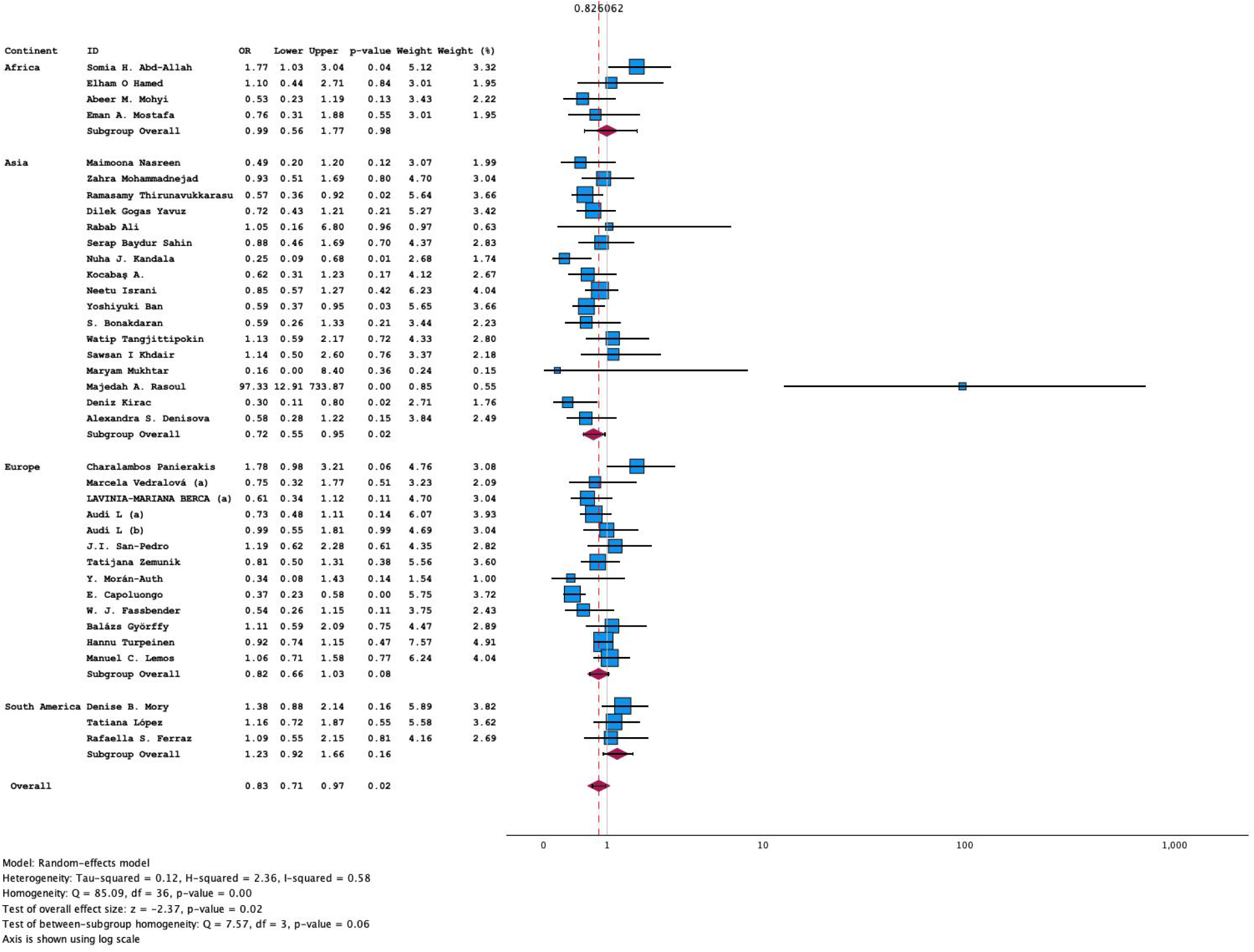
Forest plot of FokI SNP heterozygous (TC vs CC) model in T1DM

**Table 1.**
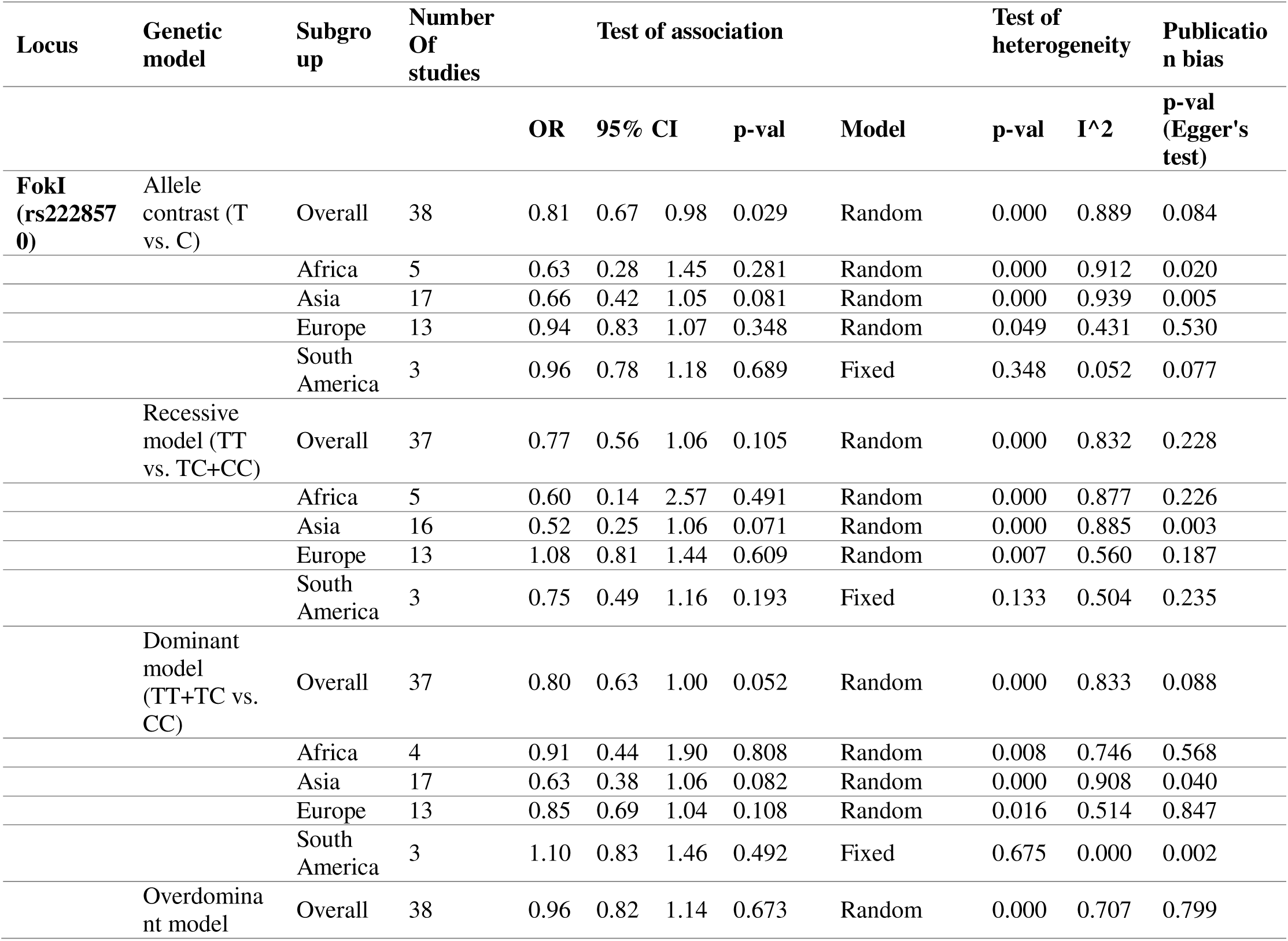
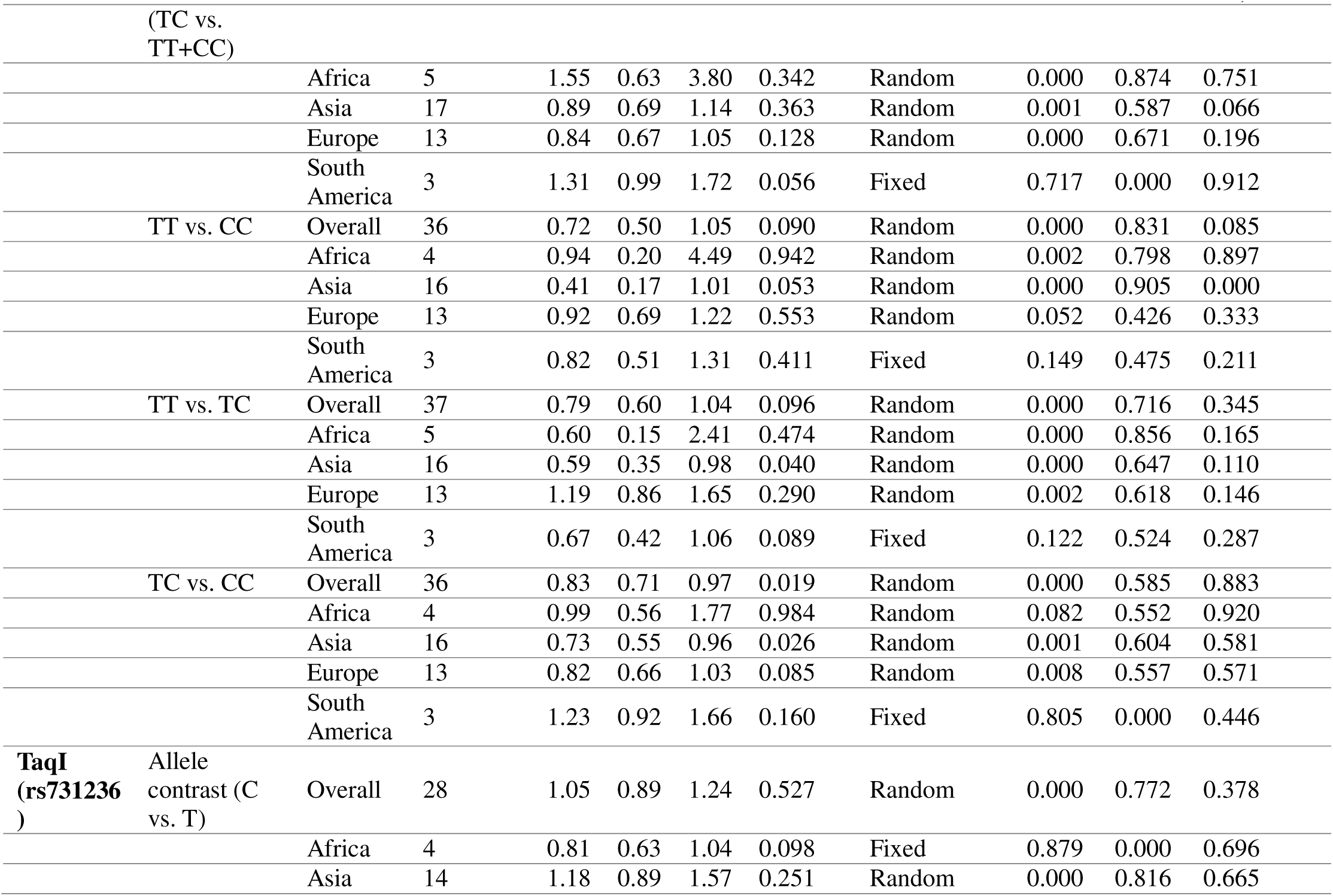
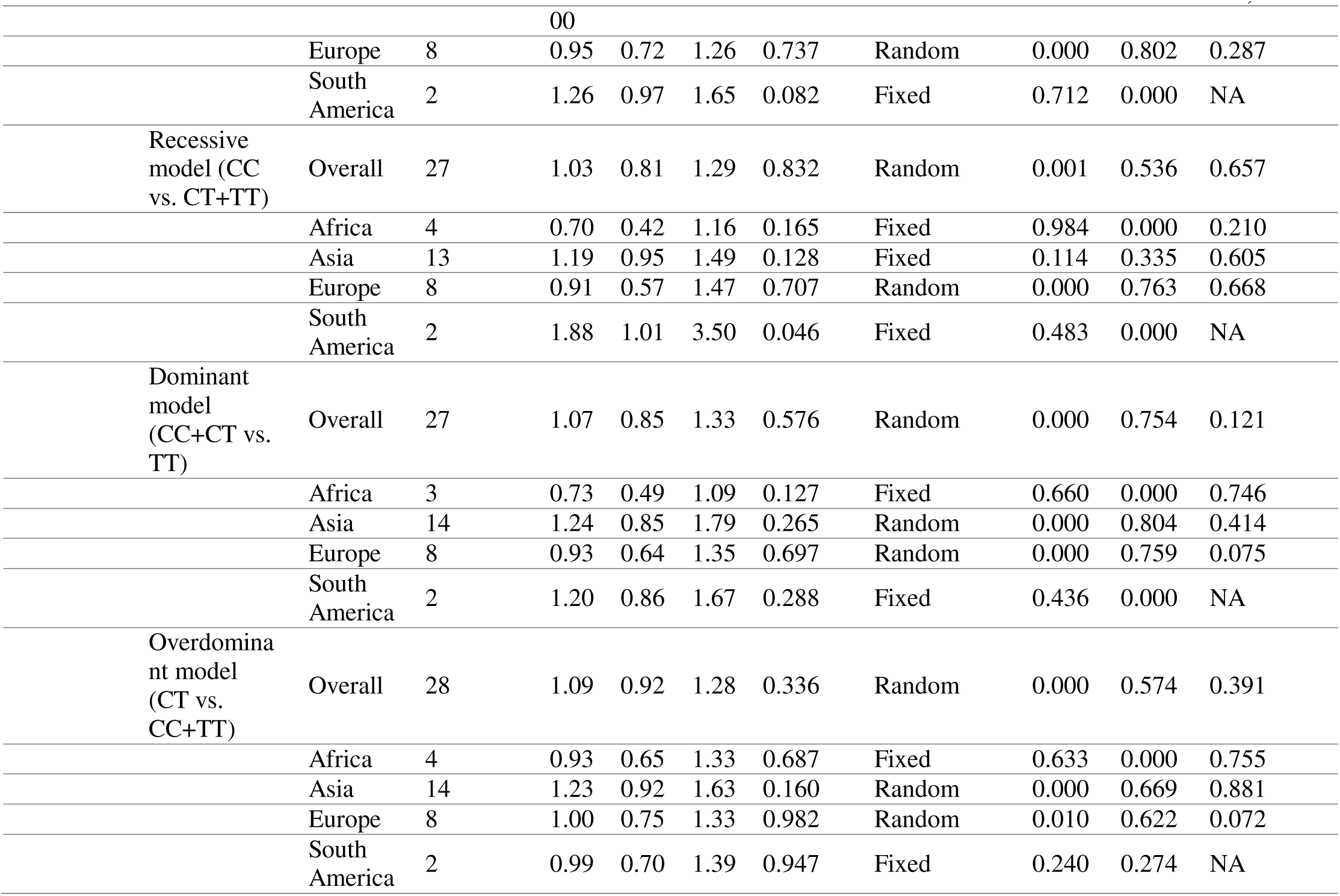
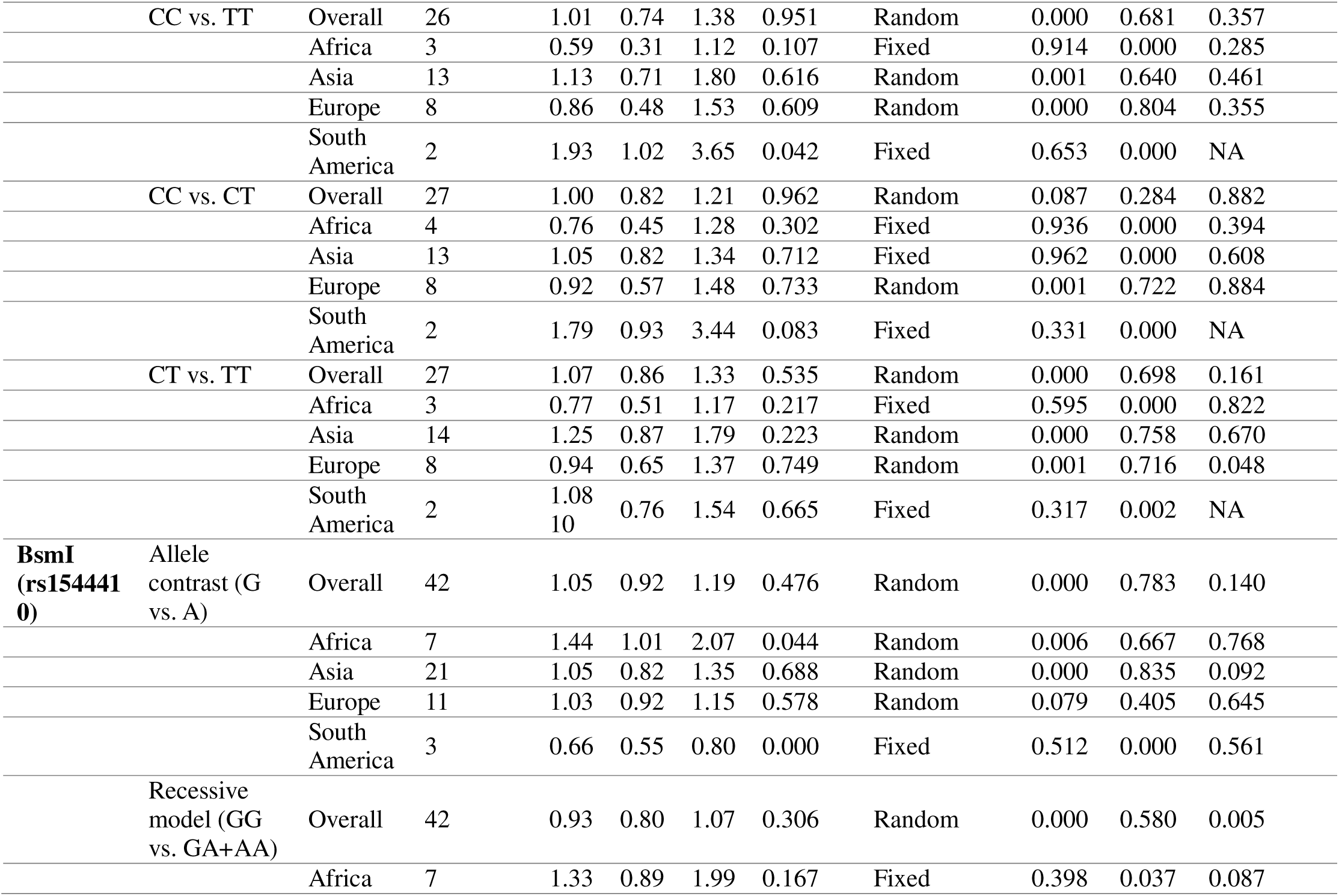
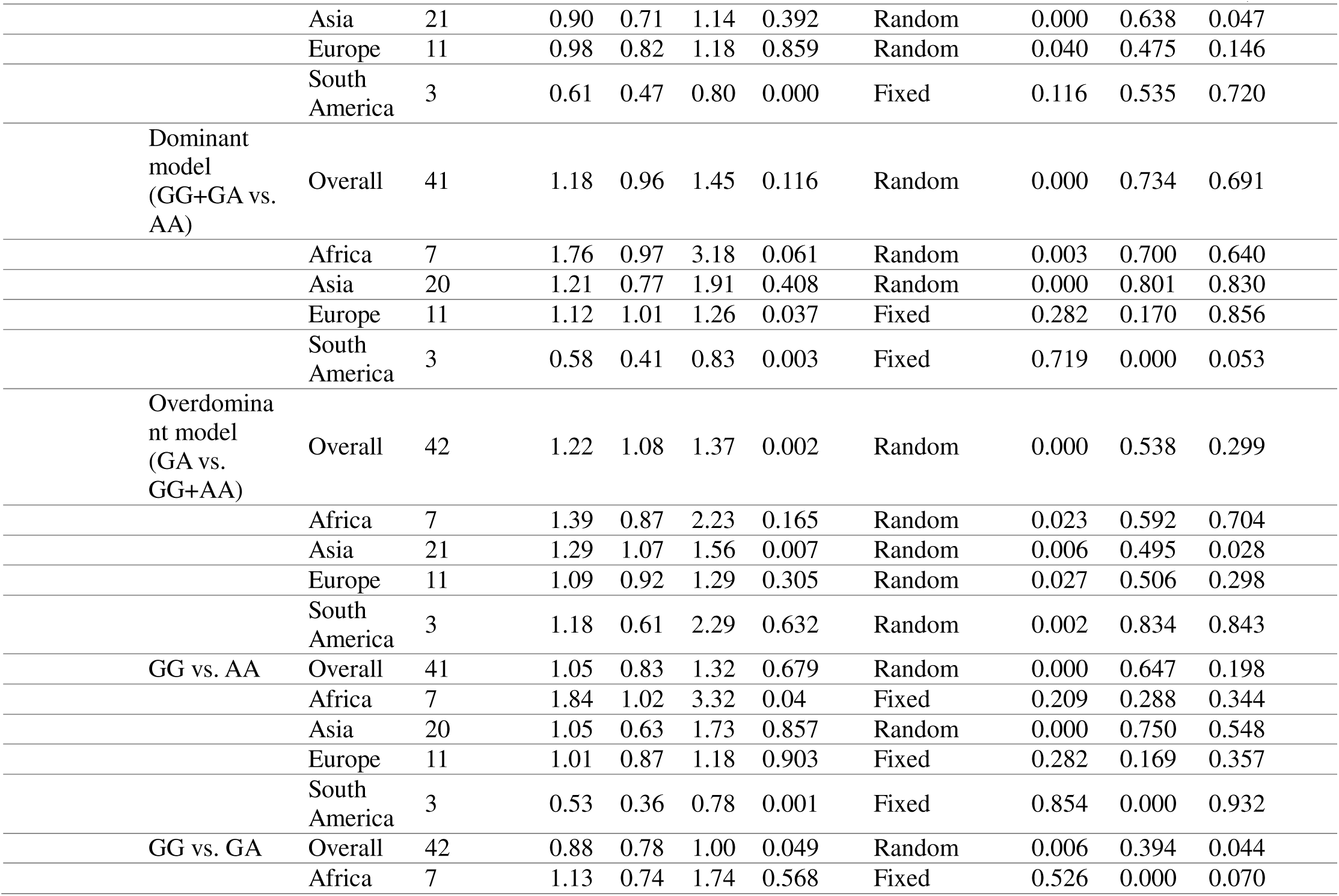
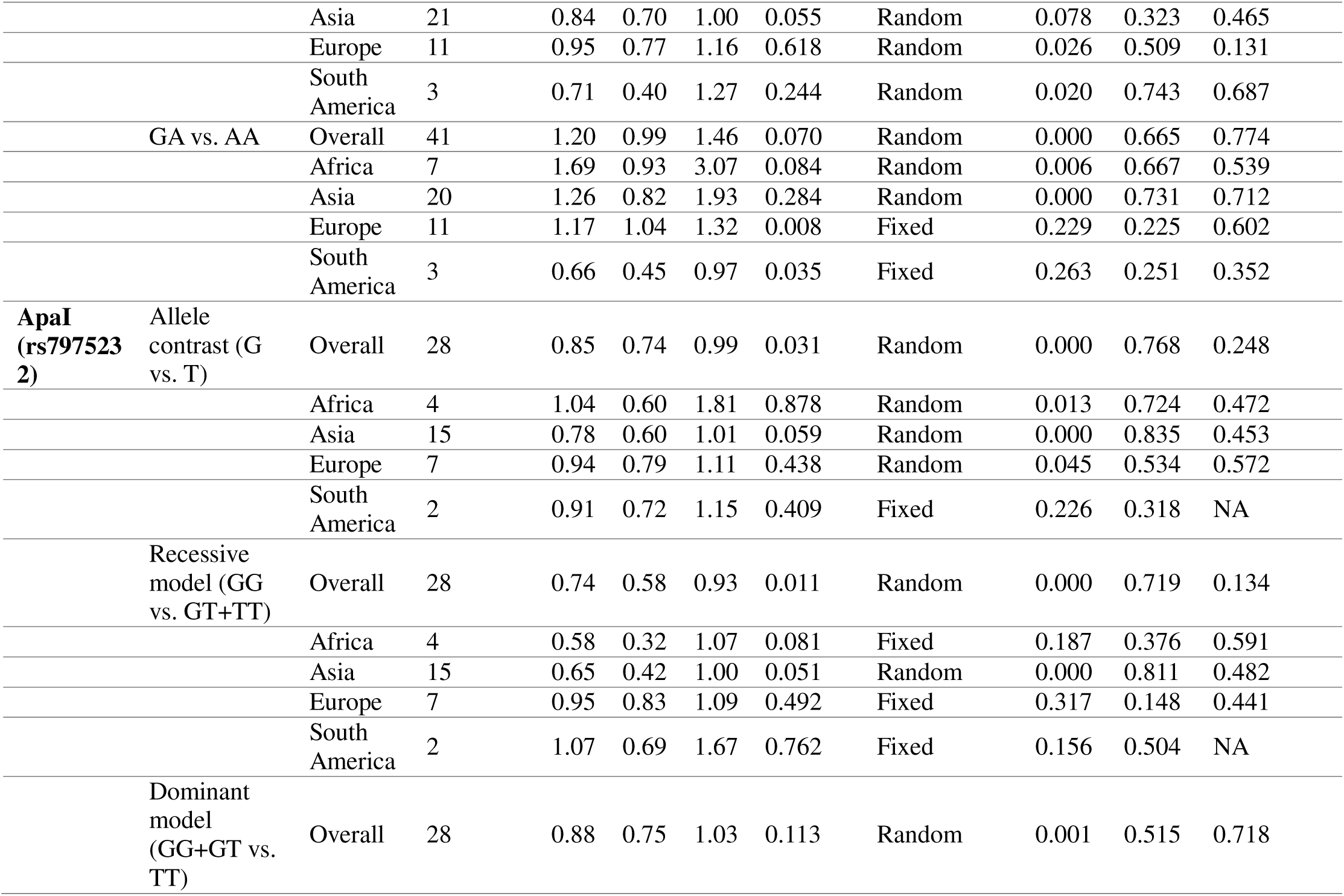
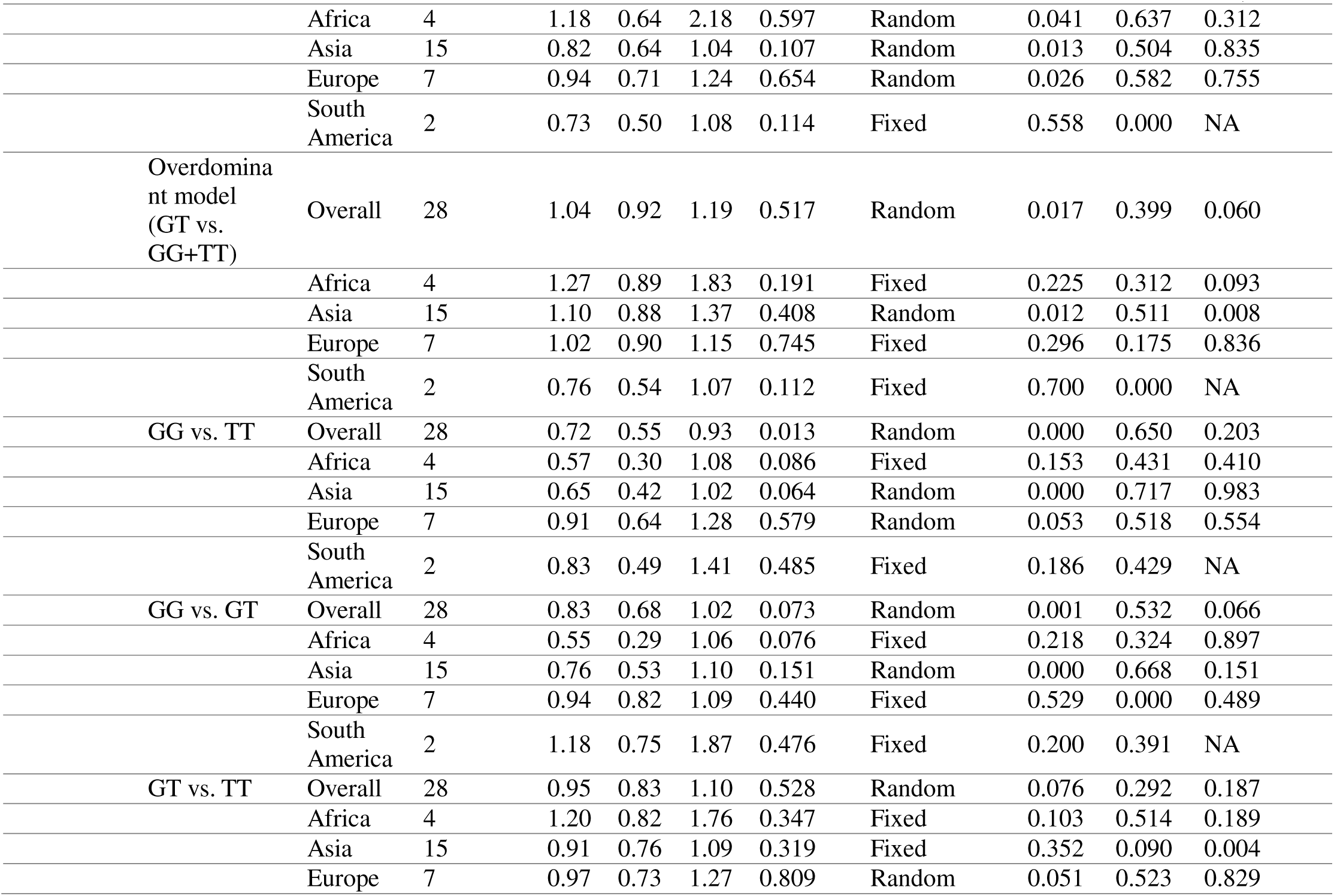
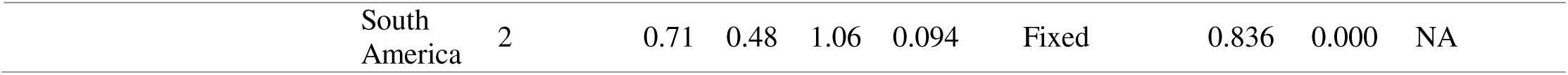
Main results of pooled odd ratios (ORs) in meta-analysis of vitamin D receptor (VDR) gene polymorphisms in association with type 1 diabetes mellitus (T1DM) risk.

#### T2DM

**Table 2** presents the findings from an extensive analysis of the FokI SNP in relation to T2DM, which utilized data from 55 studies. These studies included 7 from European countries, 30 from Asian countries, 13 from African nations, and 5 from South American countries. The overall analysis revealed no significant association between the FokI polymorphism and T2DM across all genotype models. Publication bias analysis, evaluated using Egger’s test, confirmed the absence of bias, as detailed in Table 1. However, subgroup analysis by ethnicity suggested an increased risk of T2DM in the African population across four genotype models: allelic (T vs. C, see S1, Figure 2), the recessive model (TT vs. TC+CC, see S1, Figure 3), the dominant model (TT+TC vs. CC, see S1, Figure 4), and the homozygous model (TT vs. CC, see S1, Figure 5). In contrast, among the Asian population, the (TT vs. TC) model showed a significantly reduced susceptibility to T2DM, see S1, Figure 7).

**Table 2.**
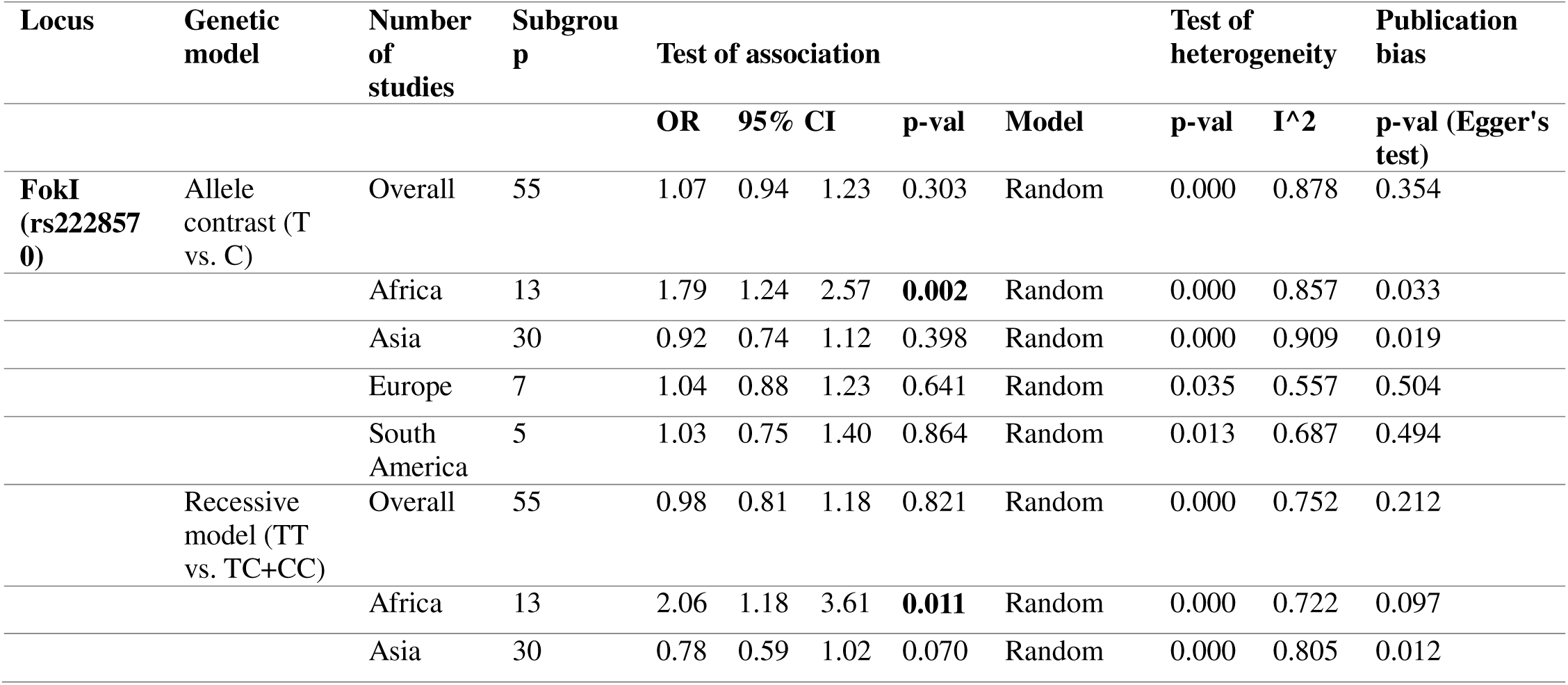
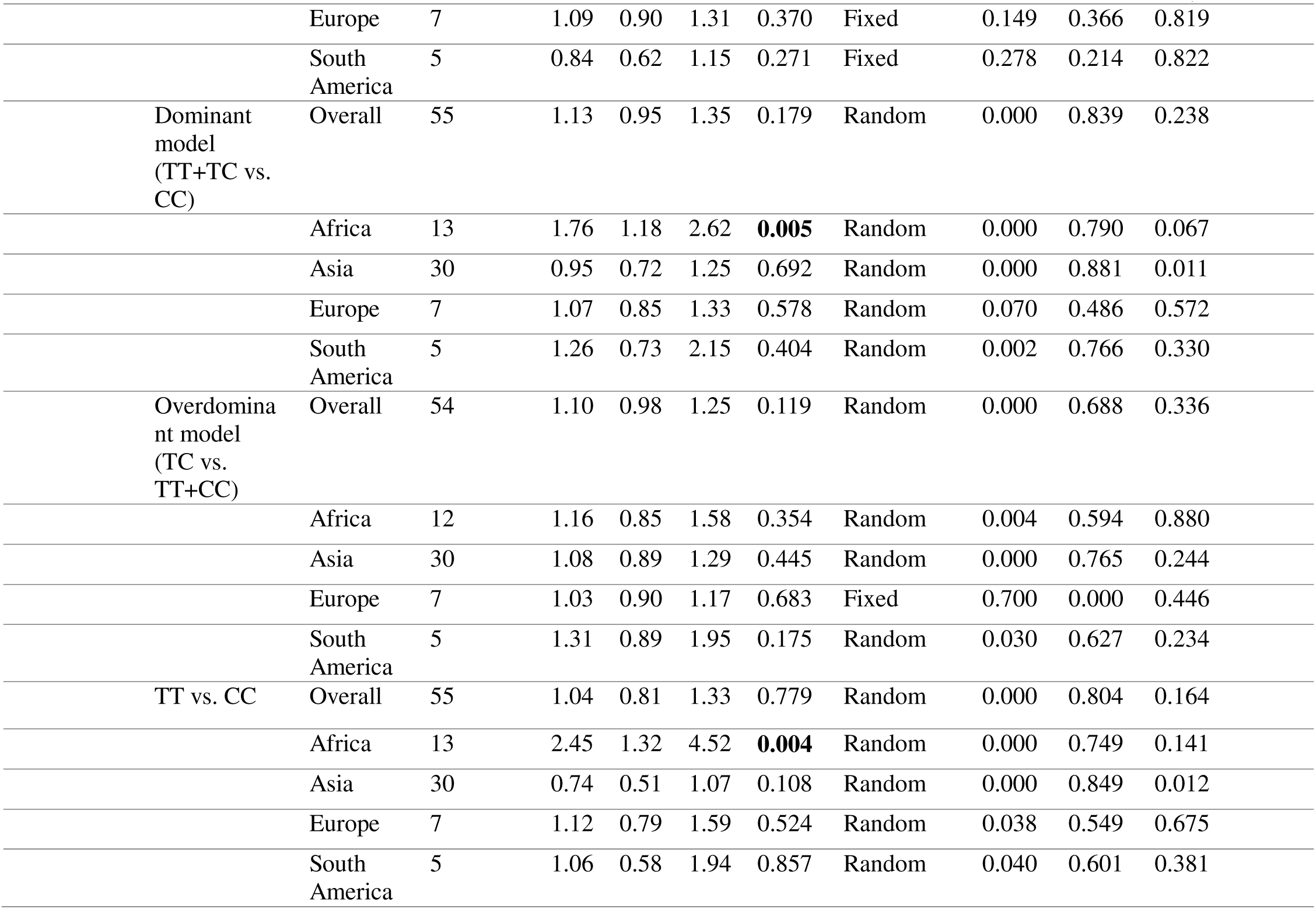
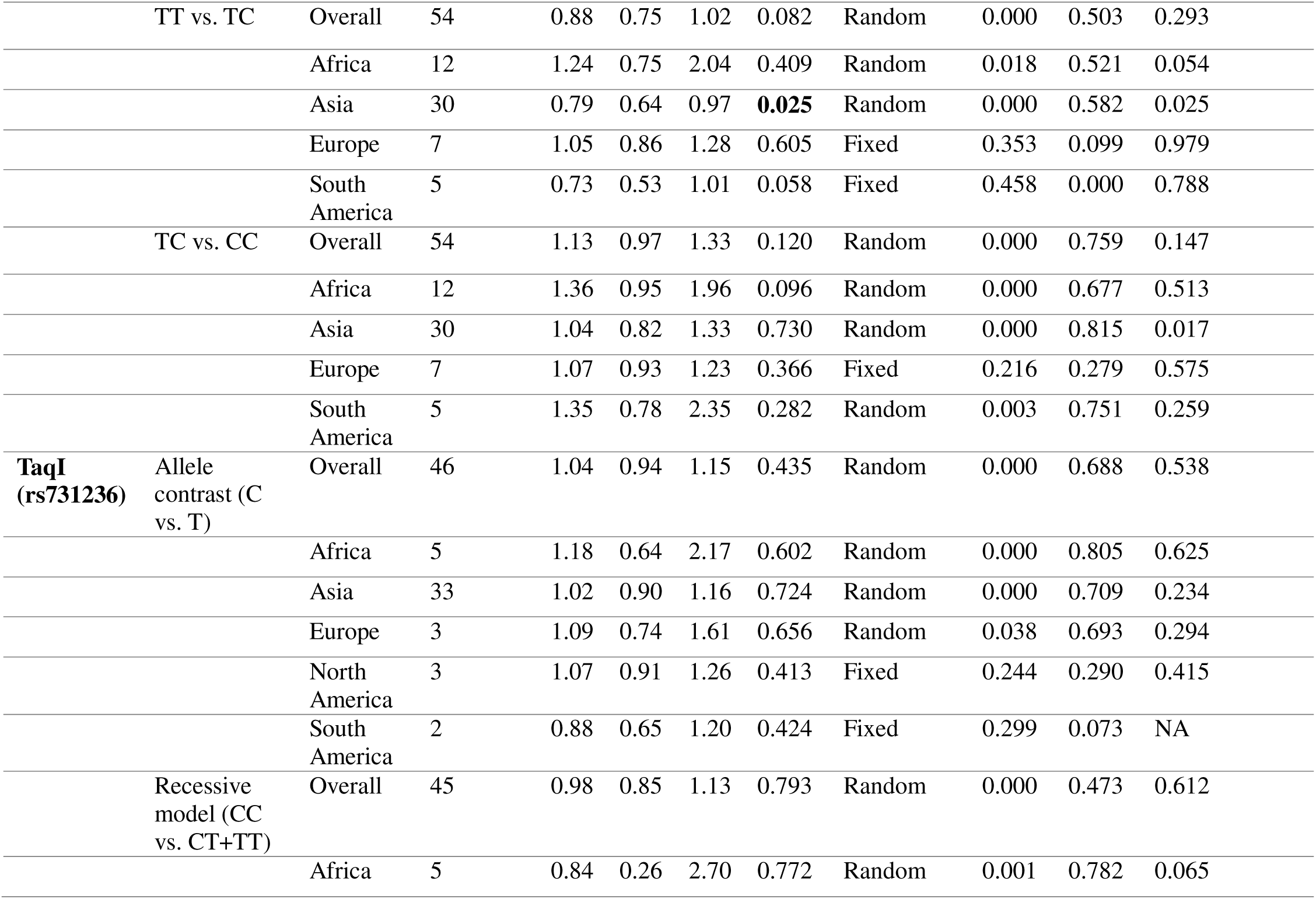
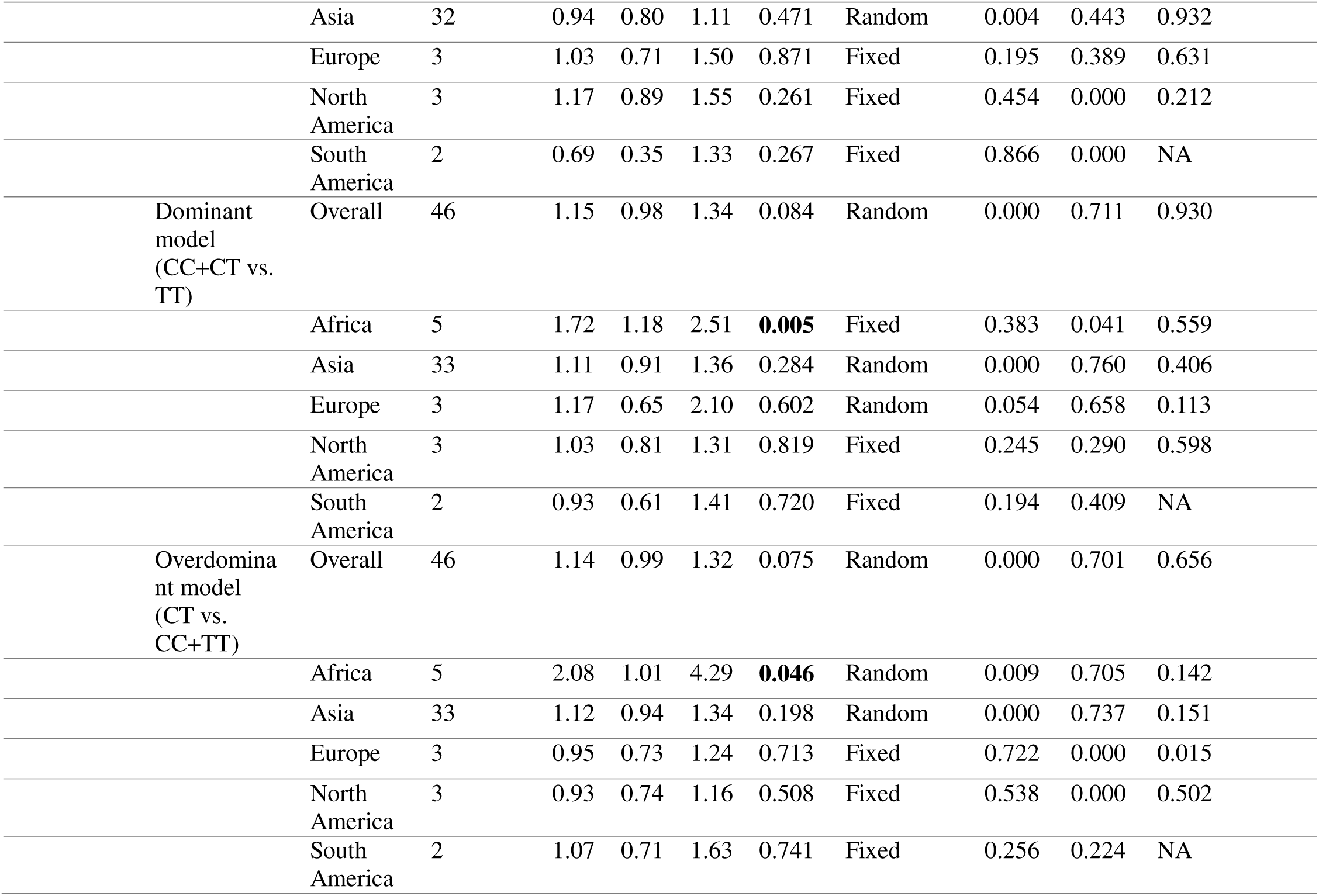
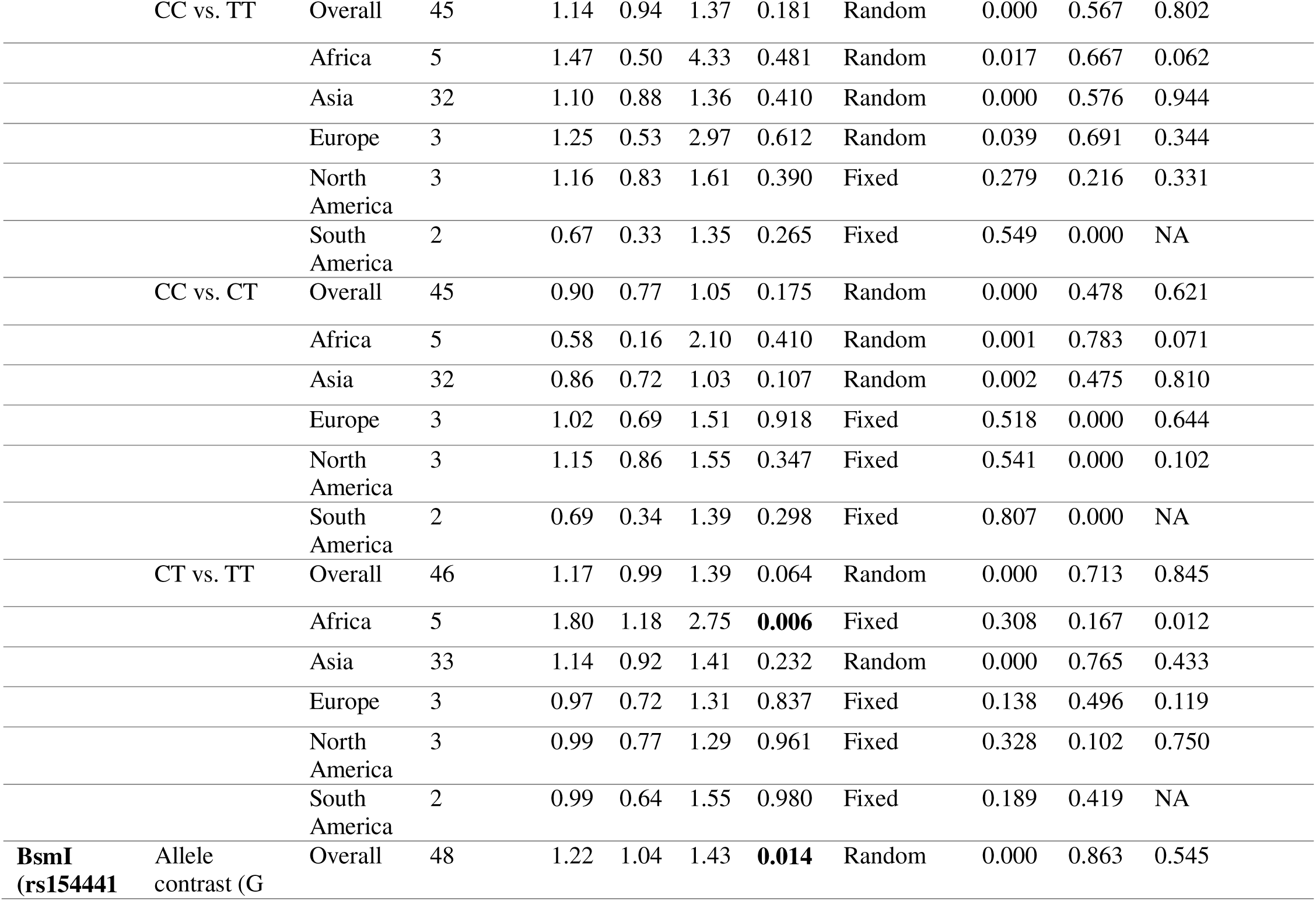
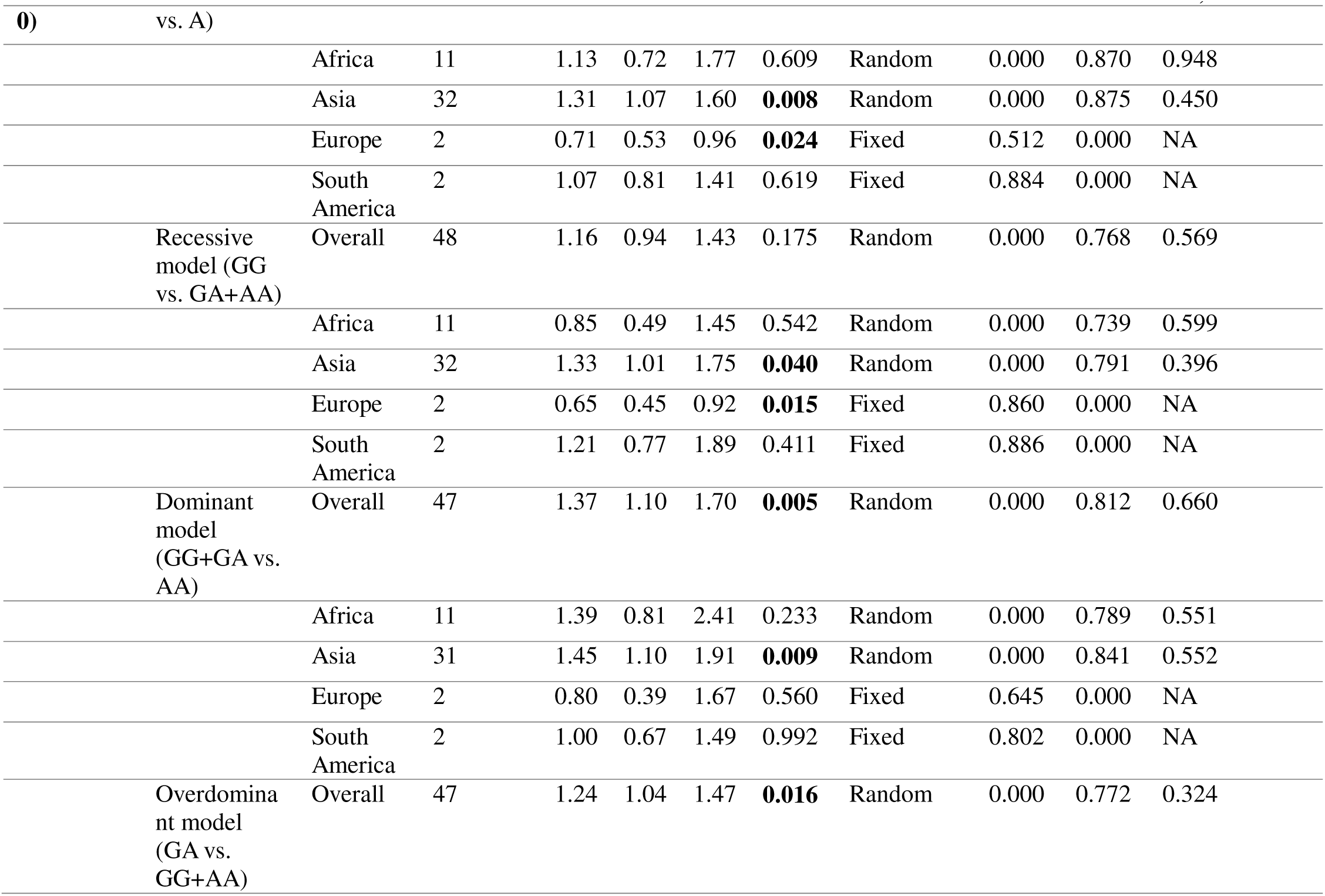
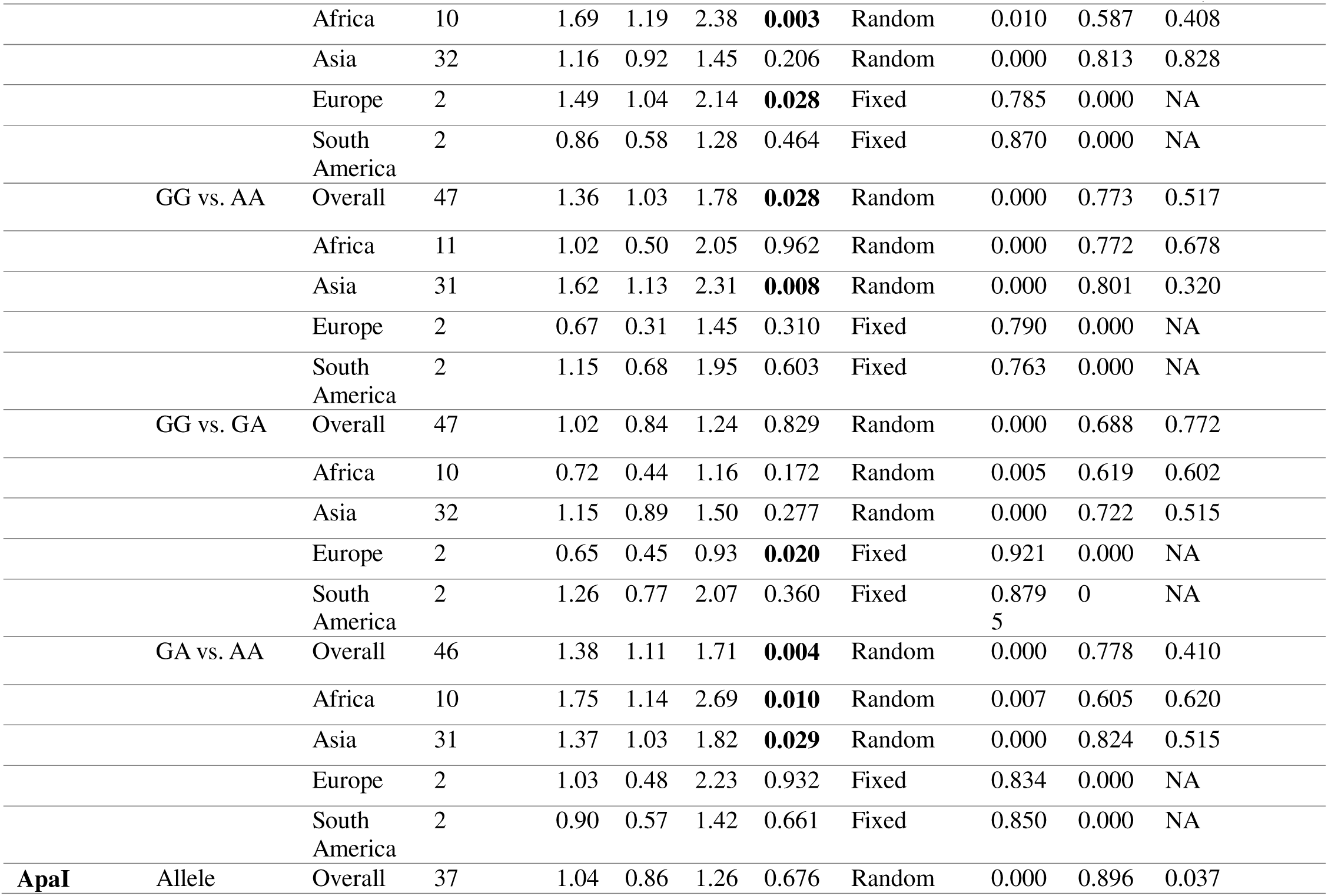
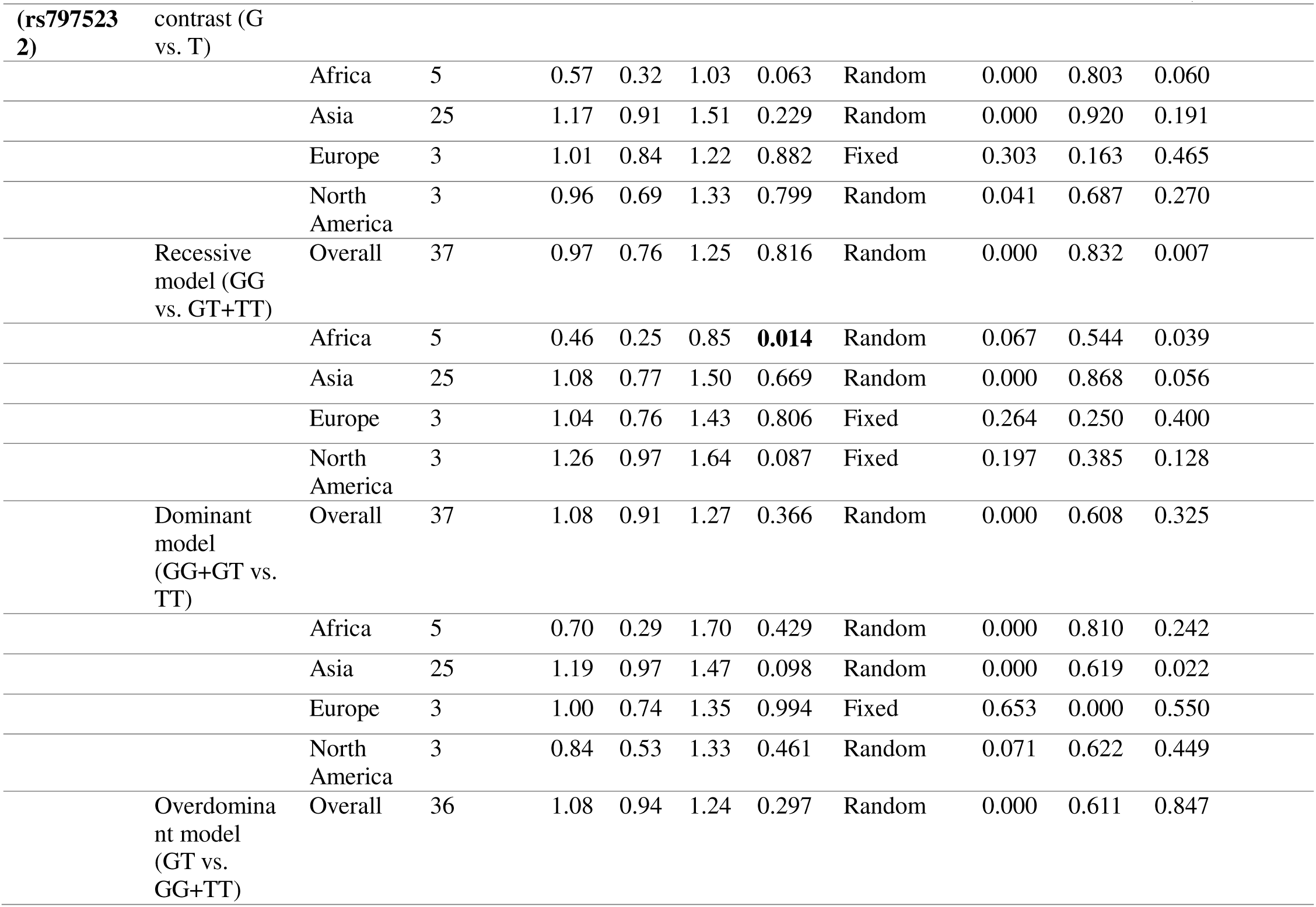
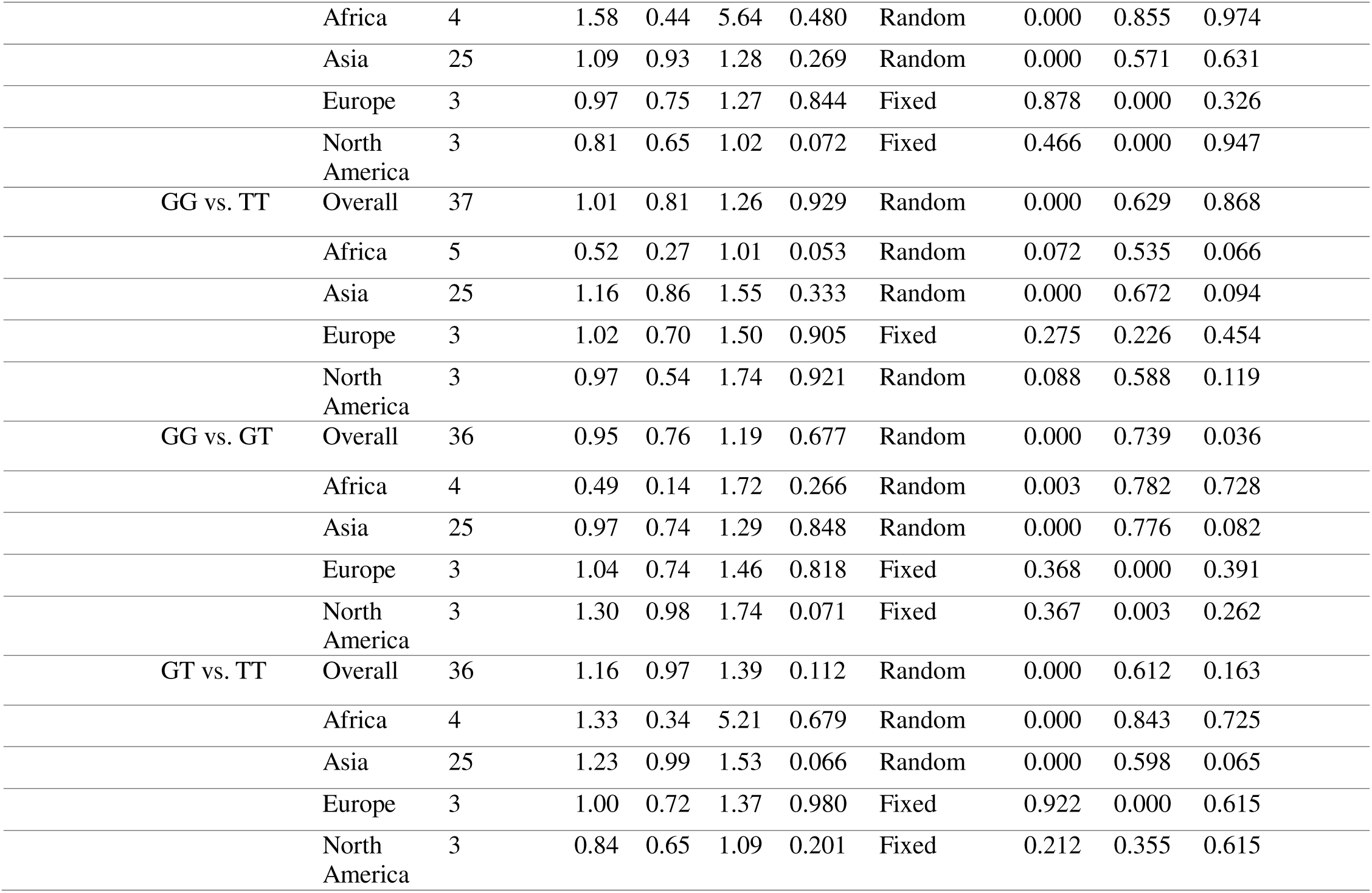
Main results of pooled odd ratios (ORs) in meta-analysis of vitamin D receptors (VDR) gene polymorphisms in association with type 2 diabetes mellitus (T2DM) risk.

#### GDM

A detailed analysis of the FokI SNP was conducted using data from 10 studies, as presented in Table 3. Of these, 8 studies were conducted in Asian countries, 1 in an African nation, and 1 in a South American. The overall pooled analysis found no significant association between the FokI polymorphism and GDM across all genotype models. Egger’s test showed no evidence of significant bias. However, subgroup analysis by ethnicity revealed a probable increased risk to GDM in the Asian population under the allelic (T vs. C) model as shown in S1, Figure 7.

**Table 3.**
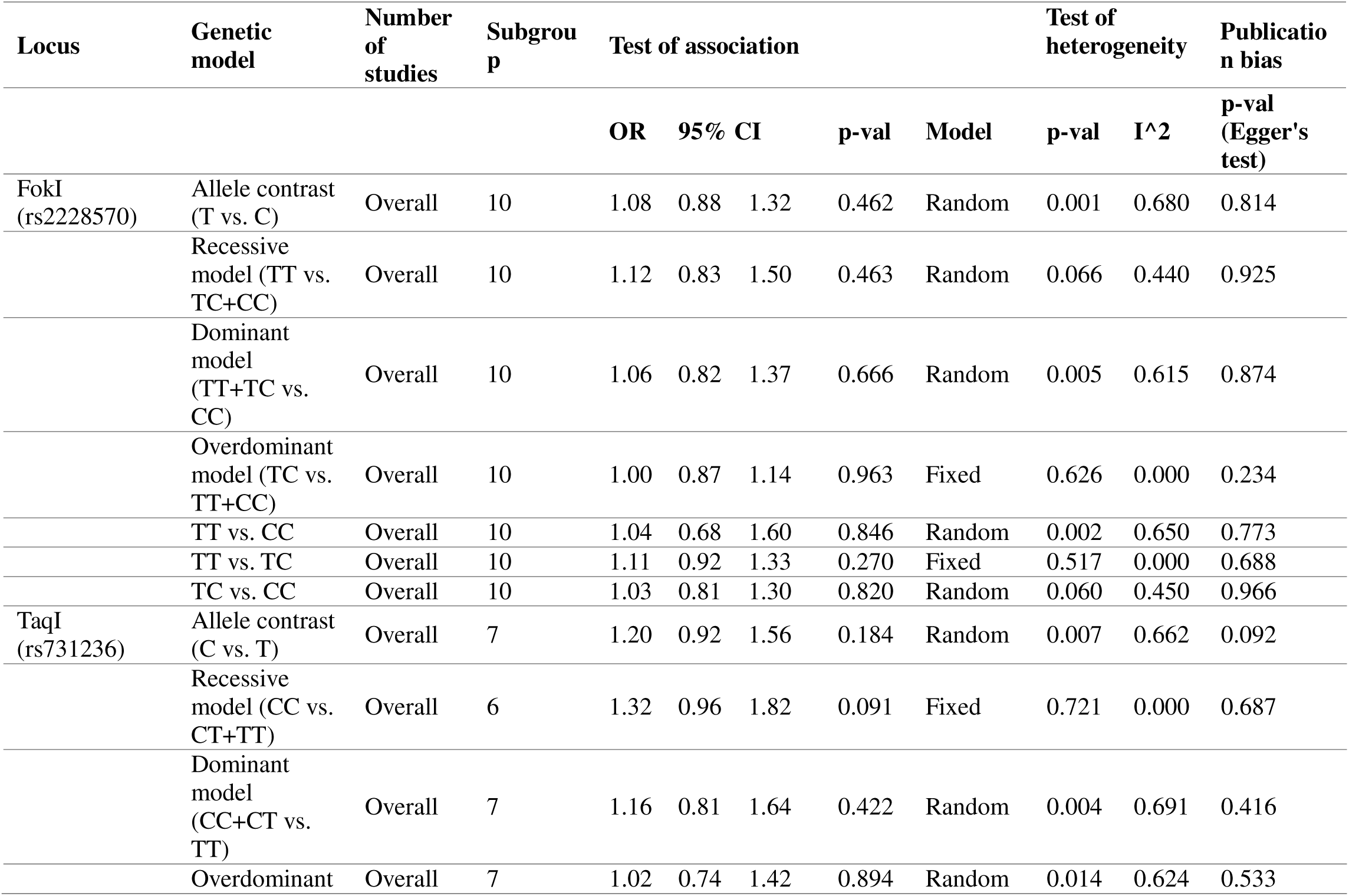
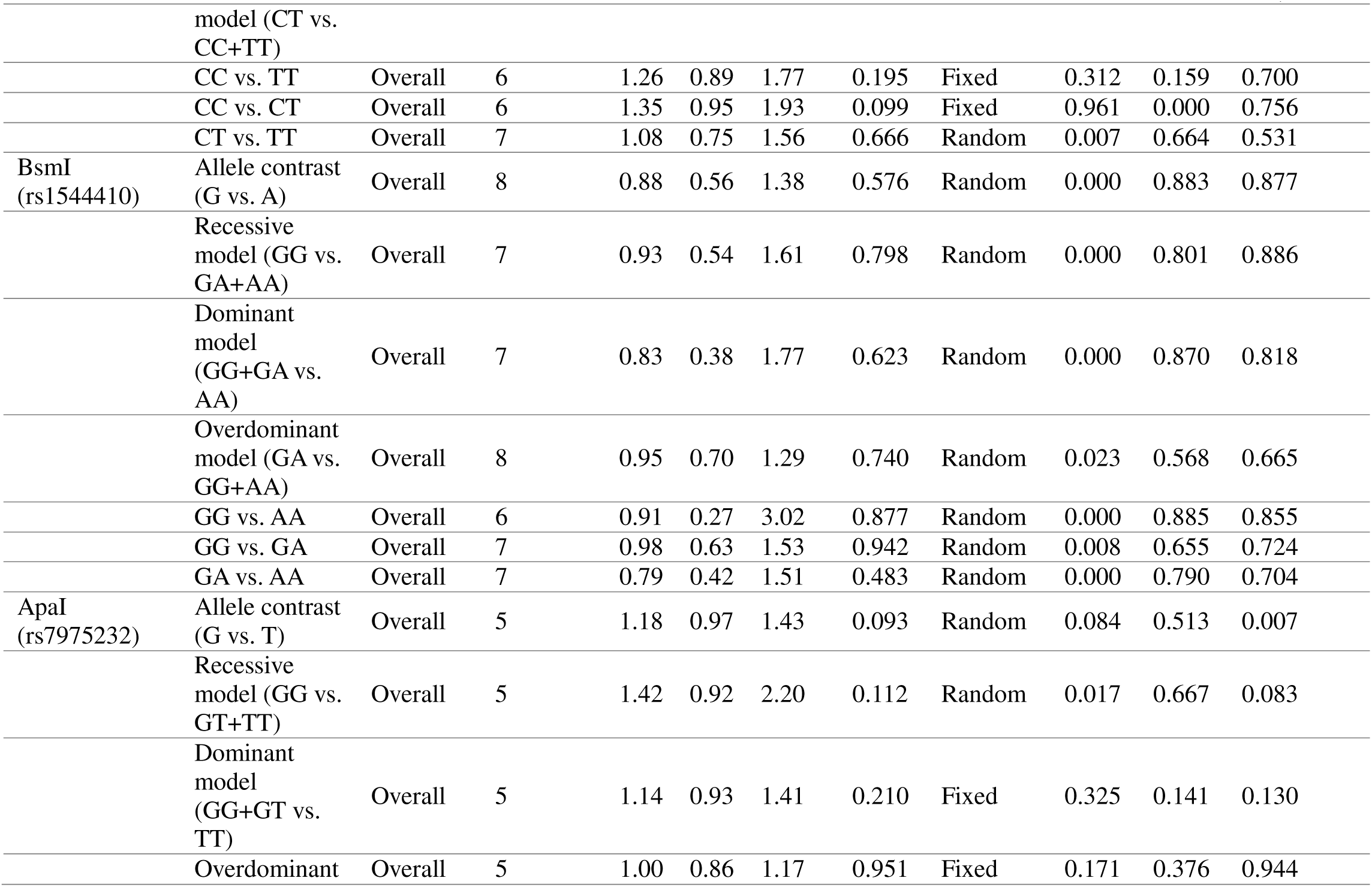
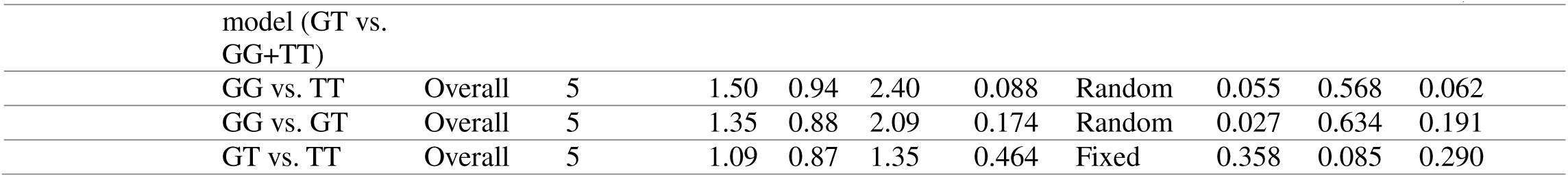
Main results of pooled odd ratios (ORs) in meta-analysis of vitamin D receptors (VDR) gene polymorphisms in association with gestational diabetes mellitus (GDM) risk.

| Locus | Genetic model | Number of studies | Subgroup | Test of association |  |  |  |  | Test of heterogeneity |  | Publication bias |
| --- | --- | --- | --- | --- | --- | --- | --- | --- | --- | --- | --- |
|  |  |  |  | OR | 95% CI |  | p-val | Model | p-val | I^2 | p-val (Egger's test) |
| FokI (rs2228570) | Allele contrast (T vs. C) | Overall | 10 | 1.08 | 0.88 | 1.32 | 0.462 | Random | 0.001 | 0.680 | 0.814 |
|  | Recessive model (TT vs. TC+CC) | Overall | 10 | 1.12 | 0.83 | 1.50 | 0.463 | Random | 0.066 | 0.440 | 0.925 |
|  | Dominant model (TT+TC vs. CC) | Overall | 10 | 1.06 | 0.82 | 1.37 | 0.666 | Random | 0.005 | 0.615 | 0.874 |
|  | Overdominant model (TC vs. TT+CC) | Overall | 10 | 1.00 | 0.87 | 1.14 | 0.963 | Fixed | 0.626 | 0.000 | 0.234 |
|  | TT vs. CC | Overall | 10 | 1.04 | 0.68 | 1.60 | 0.846 | Random | 0.002 | 0.650 | 0.773 |
|  | TT vs. TC | Overall | 10 | 1.11 | 0.92 | 1.33 | 0.270 | Fixed | 0.517 | 0.000 | 0.688 |
|  | TC vs. CC | Overall | 10 | 1.03 | 0.81 | 1.30 | 0.820 | Random | 0.060 | 0.450 | 0.966 |
| TaqI (rs731236) | Allele contrast (C vs. T) | Overall | 7 | 1.20 | 0.92 | 1.56 | 0.184 | Random | 0.007 | 0.662 | 0.092 |
|  | Recessive model (CC vs. CT+TT) | Overall | 6 | 1.32 | 0.96 | 1.82 | 0.091 | Fixed | 0.721 | 0.000 | 0.687 |
|  | Dominant model (CC+CT vs. TT) | Overall | 7 | 1.16 | 0.81 | 1.64 | 0.422 | Random | 0.004 | 0.691 | 0.416 |
|  | Overdominant | Overall | 7 | 1.02 | 0.74 | 1.42 | 0.894 | Random | 0.014 | 0.624 | 0.533 |

| Locus | Genetic model | Number of studies | Subgroup | Test of association |  |  |  | Test of heterogeneity |  | Publication bias |
| --- | --- | --- | --- | --- | --- | --- | --- | --- | --- | --- |
|  |  |  |  | OR | 95% CI | p-val | Model | p-val | I^2 | p-val (Egger's test) |
|  | model (CT vs. CC+TT) |  |  |  |  |  |  |  |  |  |
|  | CC vs. TT | Overall | 6 | 1.26 | 0.89 1.77 | 0.195 | Fixed | 0.312 | 0.159 | 0.700 |
|  | CC vs. CT | Overall | 6 | 1.35 | 0.95 1.93 | 0.099 | Fixed | 0.961 | 0.000 | 0.756 |
|  | CT vs. TT | Overall | 7 | 1.08 | 0.75 1.56 | 0.666 | Random | 0.007 | 0.664 | 0.531 |
| BsmI (rs1544410) | Allele contrast (G vs. A) | Overall | 8 | 0.88 | 0.56 1.38 | 0.576 | Random | 0.000 | 0.883 | 0.877 |
|  | Recessive model (GG vs. GA+AA) | Overall | 7 | 0.93 | 0.54 1.61 | 0.798 | Random | 0.000 | 0.801 | 0.886 |
|  | Dominant model (GG+GA vs. AA) | Overall | 7 | 0.83 | 0.38 1.77 | 0.623 | Random | 0.000 | 0.870 | 0.818 |
|  | Overdominant model (GA vs. GG+AA) | Overall | 8 | 0.95 | 0.70 1.29 | 0.740 | Random | 0.023 | 0.568 | 0.665 |
|  | GG vs. AA | Overall | 6 | 0.91 | 0.27 3.02 | 0.877 | Random | 0.000 | 0.885 | 0.855 |
|  | GG vs. GA | Overall | 7 | 0.98 | 0.63 1.53 | 0.942 | Random | 0.008 | 0.655 | 0.724 |
|  | GA vs. AA | Overall | 7 | 0.79 | 0.42 1.51 | 0.483 | Random | 0.000 | 0.790 | 0.704 |
| ApaI (rs7975232) | Allele contrast (G vs. T) | Overall | 5 | 1.18 | 0.97 1.43 | 0.093 | Random | 0.084 | 0.513 | 0.007 |
|  | Recessive model (GG vs. GT+TT) | Overall | 5 | 1.42 | 0.92 2.20 | 0.112 | Random | 0.017 | 0.667 | 0.083 |
|  | Dominant model (GG+GT vs. TT) | Overall | 5 | 1.14 | 0.93 1.41 | 0.210 | Fixed | 0.325 | 0.141 | 0.130 |
|  | Overdominant | Overall | 5 | 1.00 | 0.86 1.17 | 0.951 | Fixed | 0.171 | 0.376 | 0.944 |
|  |  |  |  | OR | 95% CI |  | p-val | Model | p-val | I^2 | p-val (Egger's test) |
|  | model (GT vs. GG+TT) |  |  |  |  |  |  |  |  |  |  |
|  | GG vs. TT | Overall | 5 | 1.50 | 0.94 | 2.40 | 0.088 | Random | 0.055 | 0.568 | 0.062 |
|  | GG vs. GT | Overall | 5 | 1.35 | 0.88 | 2.09 | 0.174 | Random | 0.027 | 0.634 | 0.191 |
|  | GT vs. TT | Overall | 5 | 1.09 | 0.87 | 1.35 | 0.464 | Fixed | 0.358 | 0.085 | 0.290 |

*The results of TaqI (rs731236) polymorphism in association with T1DM, T2DM and GDM*.

#### T1DM

An analysis of the TaqI SNP was conducted using data from 28 studies. As shown in Table 1, these included 8 studies from European countries, 14 from Asian countries, 4 from African nations, and 2 from American countries. The pooled analysis revealed no significant association between the TaqI polymorphism and T1DM across seven genotype models. Publication bias analysis showed no significant bias in any of these models, as presented in Table 1. However, a subgroup analysis by ethnicity identified an increased risk of T1DM in the South American population, specifically in two genotype models: the recessive (CC vs. CT+TT) model (see S1, Figure 8) and the homozygous (CC vs. TT) model (see S1, Figure 9).

#### T2DM

The TaqI SNP analysis included data from 46 studies, as summarized in Table 2. Among these, 3 studies were conducted in European countries, 33 in Asian countries, 5 in African countries, and 5 across North and South America (3 in North America and 2 in South America). The pooled analysis found no significant association between the FokI gene polymorphism and T2DM across any of the genotype models. Egger’s test showed no evidence of publication bias. However, ethnic subgroup analysis revealed a higher susceptibility to T2DM in the African population under the dominant (CC+CT vs. TT) model (see S1, Figure 10) and the overdominant (CT vs. CC+TT) model (see S1, Figure 11) and heterozygous (CT vs. TT) model (see S1, Figure 12).

#### GDM

The TaqI SNP analysis, covering over seven models, incorporated data from 6 to 7 studies from various countries, as outlined in Table 3. The pooled results indicated no significant association between the FokI polymorphism and GDM across multiple genotype models. Bias analysis confirmed no evidence of publication bias.

*The results of BsmI (rs1544410) polymorphism in association with T1DM, T2DM and GDM*.

#### T1DM

A comprehensive analysis of the BsmI SNP was conducted, incorporating data from 42 studies, as shown in Table 1. These included 11 studies from European countries, 21 from Asian countries, 7 from African nations, and 3 from American countries. Table 1 and **Figure 4** illustrates that the pooled analysis reveals a significant association between the BsmI polymorphism and increased risk for T1DM under the overdominant model (GA vs. GG+AA), with an OR of 1.22 (95% CI = 1.08–1.37, p = 0.002), suggesting a heightened susceptibility to T1DM. Conversely, a protective association was observed under the GG vs. GA model (OR = 0.88, 95% CI = 0.78–1.00, p = 0.049), indicating a potential reduced risk for T1DM associated with the GG genotype. Egger’s test revealed significant bias in the GG vs. GA model (p = 0.049), which suggests the possibility of small-study effects. However, the symmetrical appearance of the funnel plot and no clear indication of missing studies imply that this finding could be attributed to study heterogeneity rather than publication bias.

**Figure 4.**
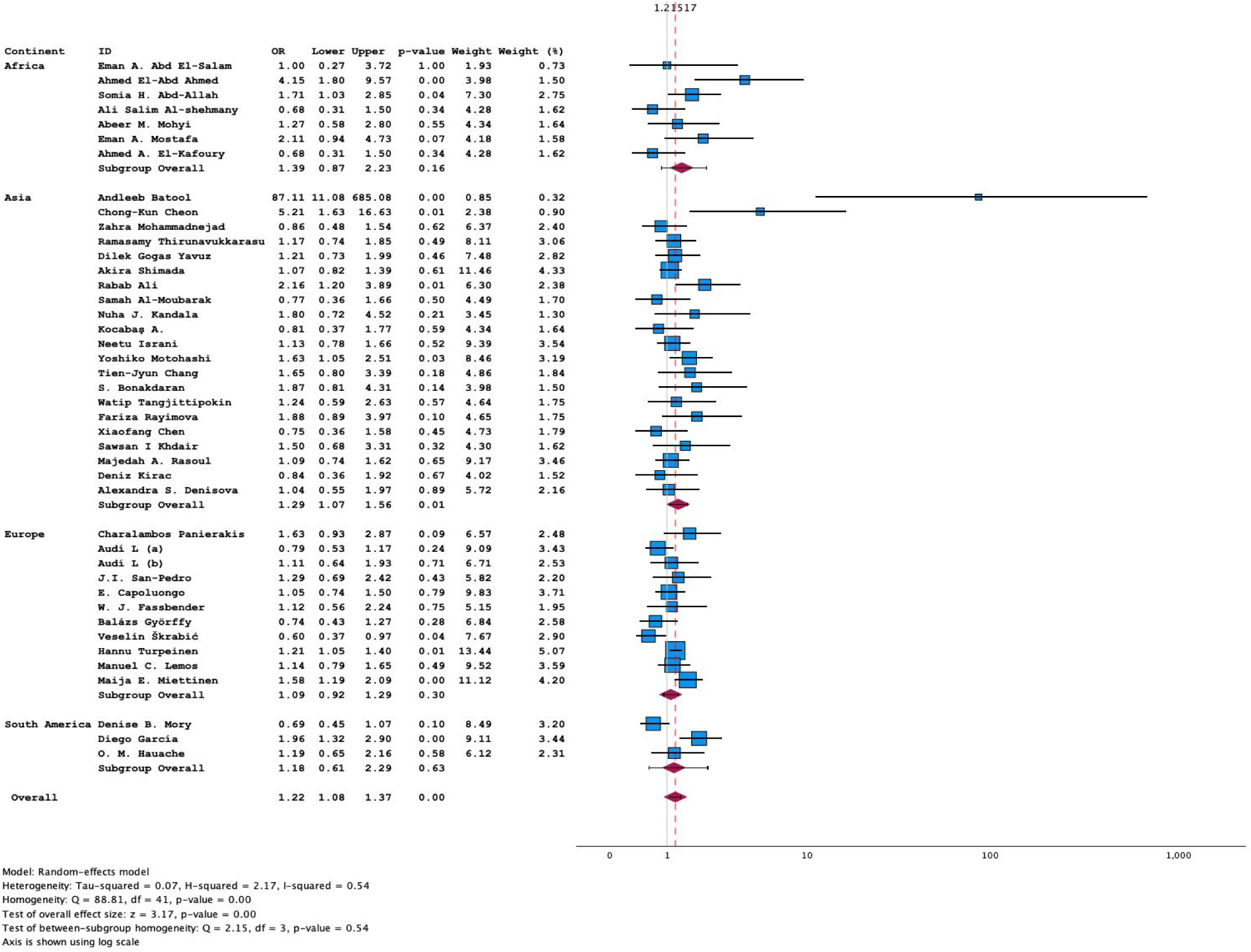
Forest plot of BsmI SNP overdominant (GA vs. GG+AA) model in T1DM

Additionally, subgroup analysis by ethnicity identified an increased risk of T1DM in across different genetic models; two genetic models allelic (G vs A, see S1, Figure 13) and homozygous (GG vs AA, see S1, Figure 14) in African, two genetic models in European population dominant (GG+GA vs. AA, see S1, Figure 15) and heterozygous GA vs. AA, see S1, Figure 16), and single model (overdominant, see **Figure 4**) in Asian. While a decreased susceptibility and protective role was observed in the South American population in almost all genetic models. These models are allelic (G vs. A, see S1, Figure 13), recessive model (GG vs. GA+AA see S1, Figure 17), dominant model (GG+GA vs. AA, see S1, Figure 15), homozygous (GG vs. AA, see S1, Figure 14), and heterozygous (GA vs. AA, see S1, Figure 16) models.

**Figure 5.**
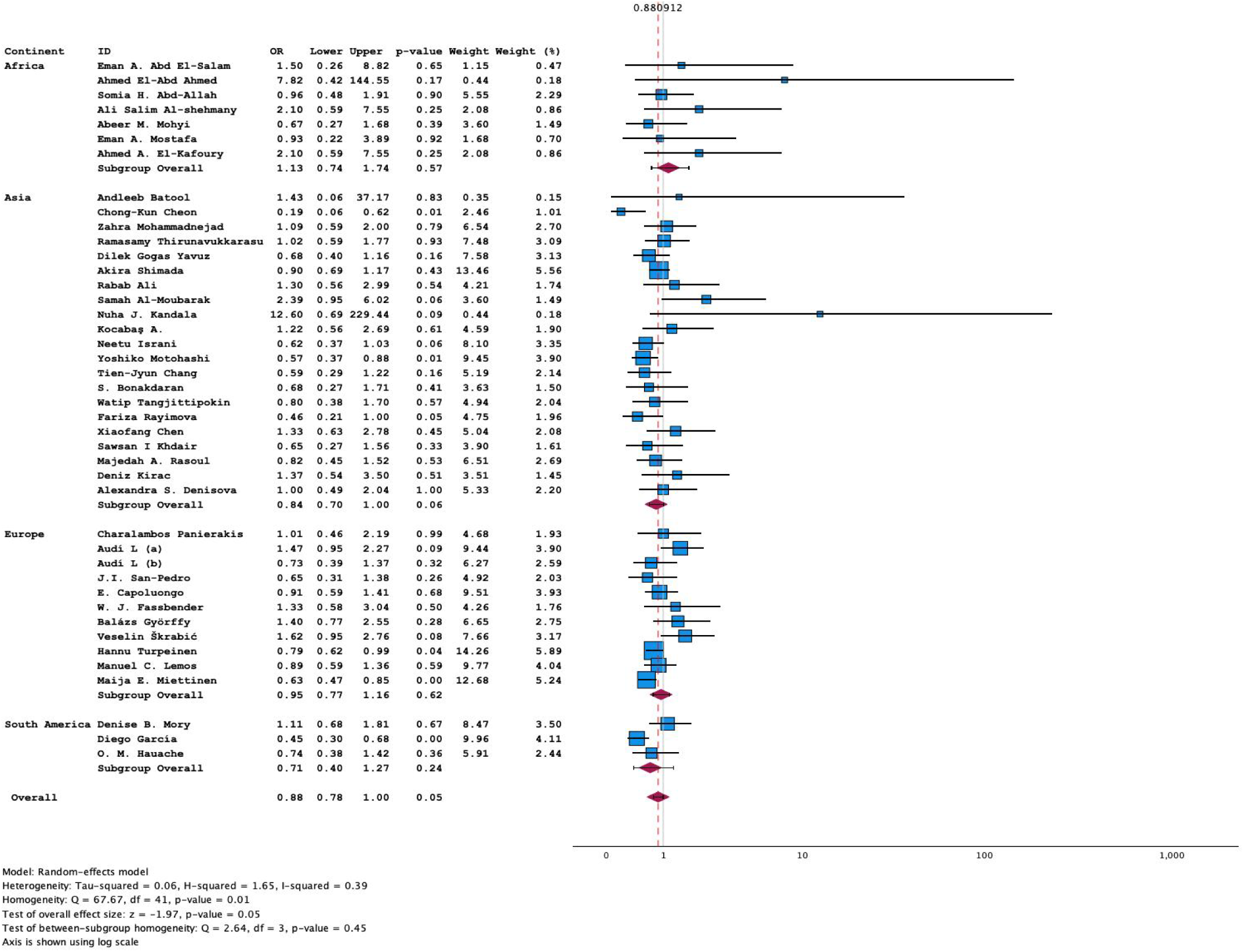
Forest plot of BsmI SNP (GG vs GA) model in T1DM

#### T2DM

A comprehensive analysis of the BsmI SNP was performed, incorporating data from 48 studies, as detailed in Table 2. Among these, 2 studies were conducted in European countries, 32 in Asian countries, 11 in African nations, and 2 in South American countries. The pooled analysis demonstrated a significant positive association between the BsmI gene polymorphism and T2DM under all models expect recessive and (GG vs GA) models. The risk association models are as follows: allelic model (G vs. A) (OR = 1.22, 95% CI = 1.04–1.43, p = 0.014) (see **Figure 6A)** and the dominant model (GG+GA vs. AA) (OR = 1.37, 95% CI = 1.10–1.70, p = 0.005) as shown in **Figure 6B**, overdominant (GA vs. GG+AA) model (OR = 1.24, 95% CI = 1.04–1.47, p = 0.016) as shown in **Figure 6D**, homozygous (GG vs. AA) model (OR = 1.36, 95% CI = 1.03–1.78, p = 0.028) as shown in **Figure 6C**, and heterozygous (GA vs. AA) model (OR = 1.38, 95% CI = 1.11–1.71, p = 0.004) as shown in **Figure 7**. No evidence of bias was detected across these models. Moreover, subgroup analysis by ethnicity revealed a higher susceptibility to T2DM within the Asian population across multiple genotype models, including allelic (G vs. A) (see **Figure 6A)**, recessive (GG vs. GA+AA) (see S1, Figure 18), dominant (GG+GA vs. AA) (see **Figure 6B**), homozygous (GG vs. AA) (see **Figure 6C**), and heterozygous (GA vs. AA) (see **Figure 7)** models.

**Figure 6.**
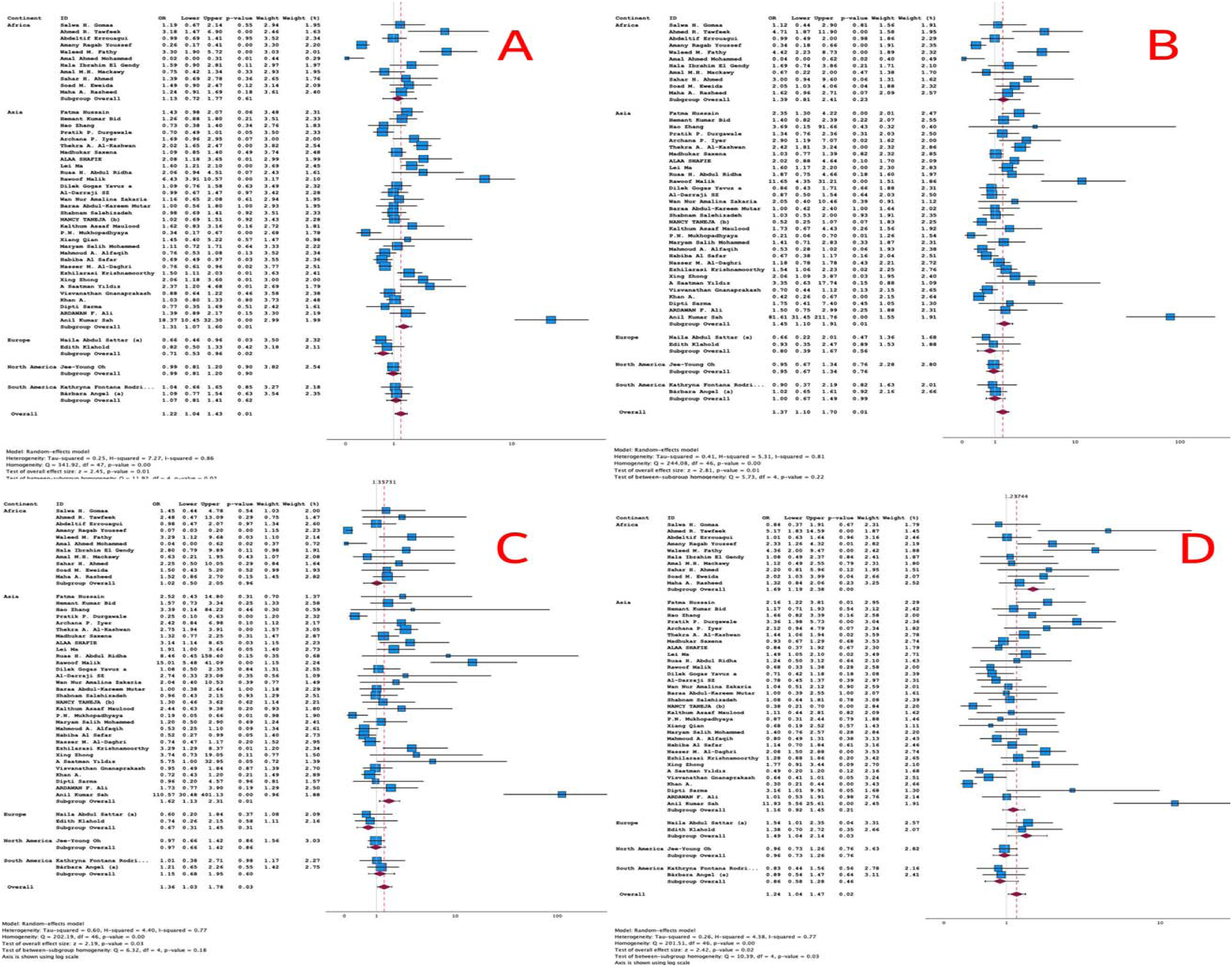
Forest plot of BsmI SNP in T2DM; A: Allelic (G vs. A) model, B: Dominant model (GG+GA vs. AA), C: Homozygous (GG vs. AA), D: Overdominant model (GA vs. GG+AA).

**Figure 7.**
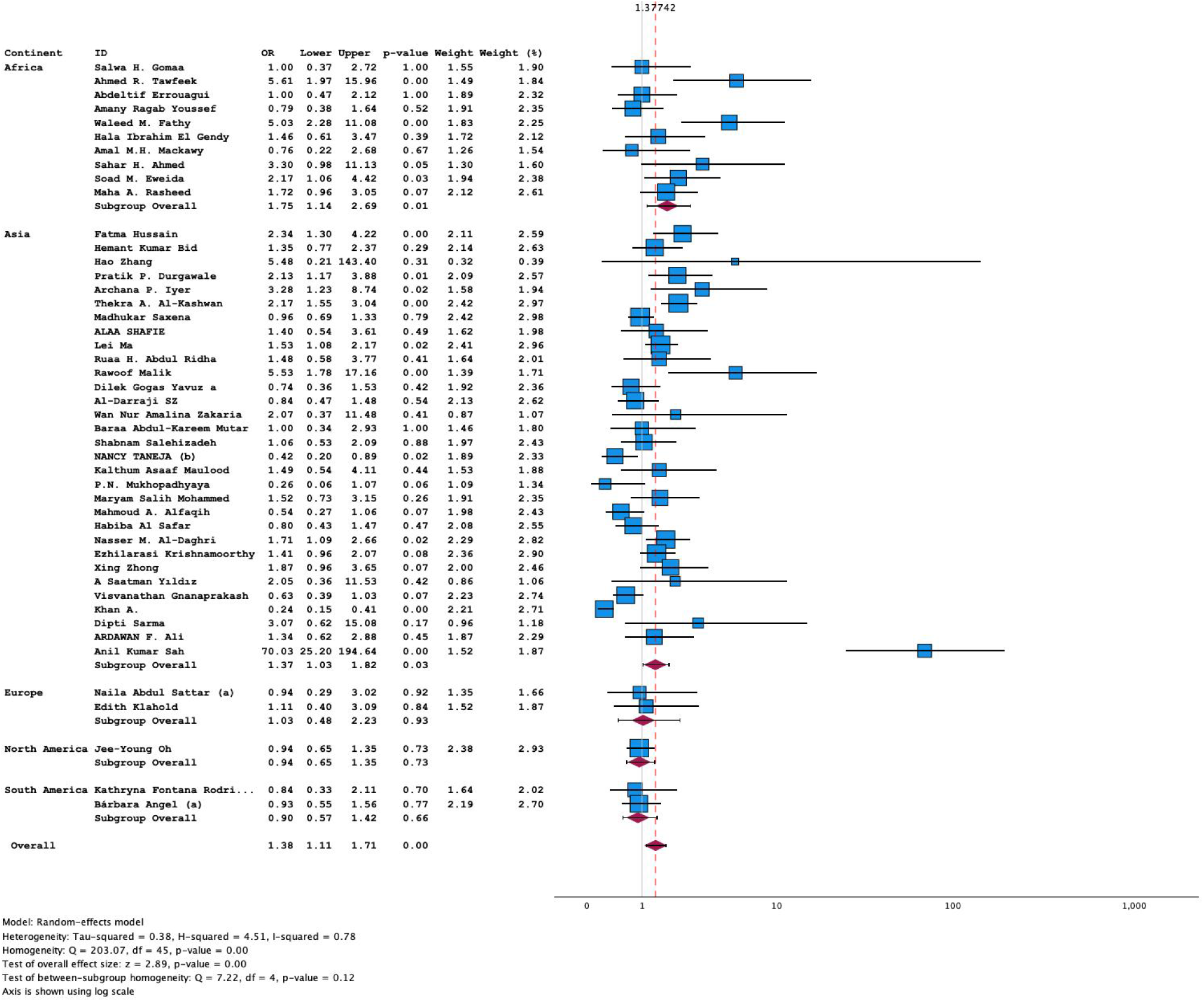
Forest plot of BsmI SNP heterozygous (GA vs. AA) model in T2DM

The European population exhibited increased susceptibility under the overdominant model (GA vs. GG+AA, **see Figure 6D**), while the African population showed similar results under the overdominant (GA vs. GG+AA, **see Figure 6D**) and heterozygous (GA vs. AA, see **Figure 7**). In contrast, protective association toward T2DM was observed in the European population under the allelic (G vs. A) model (see **Figure 6A)**, recessive (GG vs. GA+AA) model (see **S1, Figure 18**), and (GG vs. GA) models (**see S1, Figure 19)**.

#### GDM

The analysis of the BsmI SNP, encompassing more than seven models, included data from 6, 7, or 8 studies from various countries, as shown in Table 3. The pooled analysis demonstrated no significant association between the BsmI polymorphism and GDM across the different genotype models. Additionally, bias analysis confirmed the absence of publication bias.

*The results of ApaI (rs7975232) polymorphism in association with T1DM, T2DM and GDM*.

#### T1DM

An overall analysis of this SNP, incorporating data from 28 studies, is presented in Table 1. Of these, 7 studies were conducted in European countries, 15 in Asian countries, 4 in African countries, and 2 in South American countries. The forest plot in **Figure 8A-C** indicates that the pooled results revealed a significant negative and protective association between the ApaI gene polymorphism and T1DM across three genotype models: allelic (G vs. T) (OR = 0.85, 95% CI = 0.74–0.99, p = 0.031), the recessive model (GG vs. GT+TT) (OR = 0.74, 95% CI = 0.58–0.93, p = 0.011), and the homozygous model (GG vs. TT) (OR = 0.72, 95% CI = 0.55–0.93, p = 0.013). Bias analysis indicated no significant publication bias, as shown in Table 1. Additionally, subgroup analysis found no significant differences across ethnic groups in terms of susceptibility to T1DM for any of the seven genotype models.

**Figure 8.**
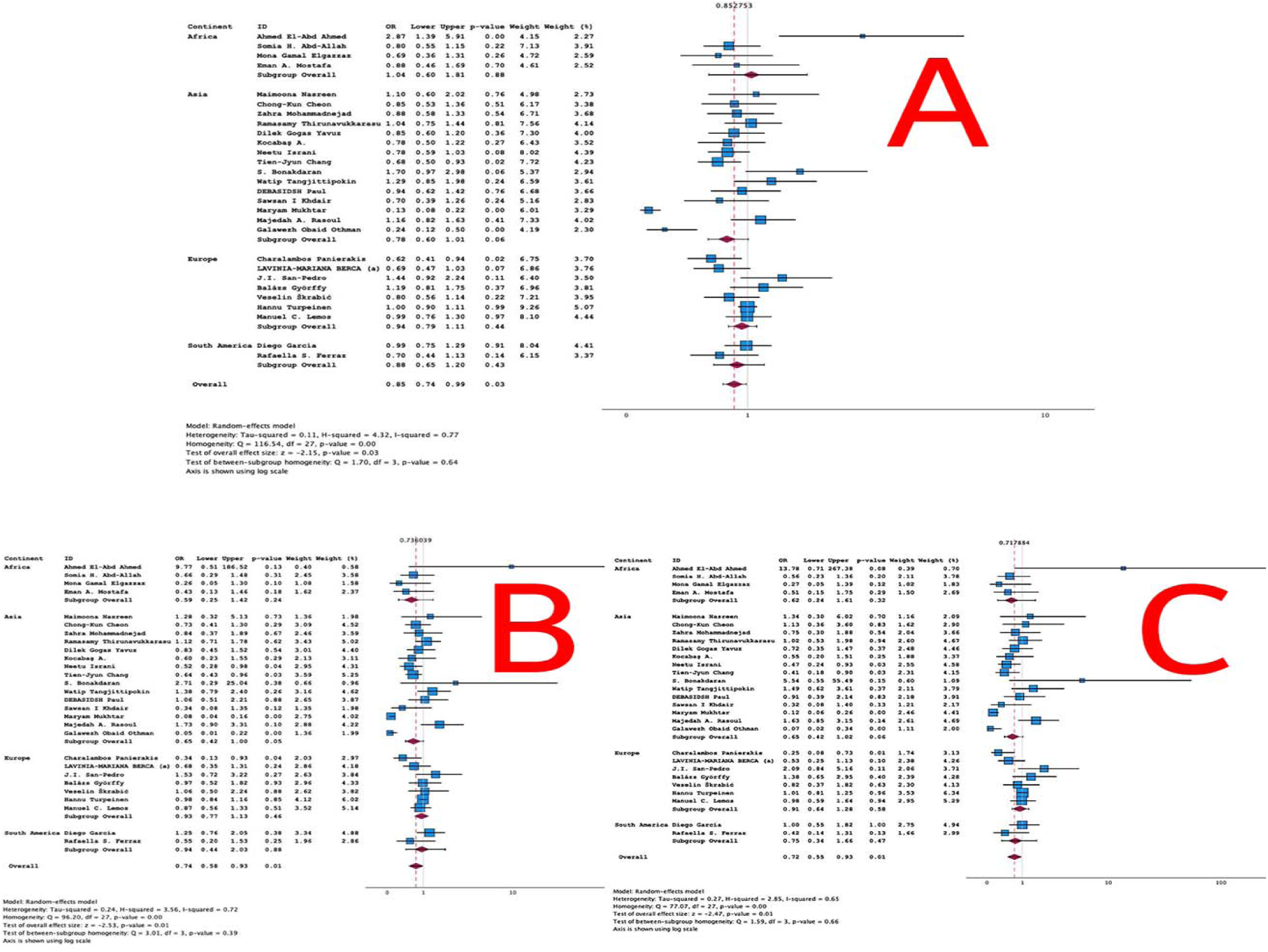
Forest plot of ApaI SNP in T1DM; A: Allelic (G vs. T) model, B: Recessive model (GG vs. GT+TT) model, C: Homozygou (GG vs. TT) model

#### T2DM

An extensive analysis of the ApaI SNP was conducted, utilizing data from 37 studies, as outlined in Table 2. These included 3 studies from European countries, 25 from Asian countries, 5 from African nations, and 3 from North American countries. The pooled analysis found no significant association between the ApaI gene polymorphism and T2DM across all genotype models, including the dominant model. However, bias analysis revealed significant publication bias in the allelic (G vs. T), recessive (GG vs. GT+TT), and (GG vs. GT) models, with missing studies in each case. Furthermore, subgroup analysis by ethnicity indicated reduced susceptibility to T2DM in the African population under the recessive model (GG vs. GT+TT, see S1, Figure 20).

#### GDM

The analysis of the ApaI SNP, covering more than seven models, included data from five studies representing various countries, as shown in Table 3. Bias analysis revealed significant bias in the allelic (G vs. T) model, with 2 missing studies on the left side of the funnel plot and imputing these studies would not alter the results. The pooled analysis found no significant association between the ApaI gene polymorphism and GDM across the different genotype models.

### Subgroup analysis between T1DM, T2DM and GDM

A subgroup analysis was performed to assess variations in four VDR gene polymorphisms among different diabetes types—T1DM, T2DM, and GDM. Notably, in the BsmI dominant C, a significant association emerged across the diabetes types, with T2DM showing an effect size of ES = 0.317 (p = 0.017), suggesting a positive association with increased susceptibility to T2DM. Likewise, in the GA vs. AA comparison, a significant effect size of ES = 0.330 (p = 0.008) was observed.

### Meta-regression analyses

We conducted a meta-regression analysis to identify the primary contributors to heterogeneity within the VDR gene polymorphism data related to diabetes mellitus, as detailed in S1, Table 3. The results indicated that age, ethnicity, and duration of illness (DOI) were significant determinants of variability in the FokI (TC vs. CC) and ApaI (G vs. T) gene models exclusively in T1DM. In contrast, no significant factors were identified to account for the heterogeneity observed in T2DM and GDM.

## Discussion

### Association between FokI (rs2228570) polymorphism and Type 1, Type 2 and GDM

The initial findings of this study indicate that the FokI (rs2228570) polymorphism, when analyzed under the allelic model (T vs. C) and the heterozygous model (TC vs. CC) across mixed ethnicities, demonstrates a significant association with T1DM in comparison to controls, suggesting a potential trend towards reduced susceptibility to T1DM in carriers of the T allele. Subgroup analyses further revealed significant associations across various ethnic groups; for instance, the TT vs. TC and TC vs. CC models showed notable differences between Asian individuals with T1DM and controls, indicating that these genotypic configurations may confer a decreased susceptibility to T1DM within the Asian population. In T2DM, analysis of the FokI polymorphism across mixed ethnicities (overall) revealed no significant differences between T2DM patients and controls. However, subgroup analyses identified significant associations within specific populations. In the African population, significant differences emerged under the allelic model (T vs. C), recessive model (TT vs. TC+CC), dominant model (TT+TC vs. CC), and homozygous model (TT vs. CC), suggesting an increased susceptibility to T2DM associated with the TT genotype. Conversely, in the Asian population, the heterozygous model (TT vs. TC) demonstrated a significant difference between T2DM patients and controls, implying a potential protective effect against T2DM. In GDM, no significant associations were observed across any genetic models when analyzed under mixed ethnicity.

Recently, a meta-analysis by Zhai et. al indicated no significant overall association between VDR polymorphisms and T1DM risk in the general population. However, subgroup analysis based on ethnicity revealed important insights FokI polymorphism: Associated with a decreased risk of T1DM in European populations, while African populations showed an increased risk under all genotype models (Zhai et al., 2020b). Another small-scale meta-analysis based on 7 studies indicated that no significant associations were found between FokI polymorphism and T1DM risk (Shahmoradi et al., 2021). However, previous studies did not incorporate all available studies examining various genetic variants in their analyses, potentially limiting the comprehensiveness of their findings. In contrast, our study pooled the overall odds ratios from 38 studies, enabling a more extensive and inclusive analysis. Furthermore, we conducted subgroup analyses based on ethnicity, which enhances the robustness and generalizability of our results by accounting for potential ethnic-specific genetic variations.

The FokI (rs2228570) polymorphism within the VDR gene, located on chromosome 12q13, holds significant implications for diabetes pathogenesis through its influence on immune responses and metabolic pathways (Apaydın et al., 2019b; Moura et al., 2024). This variant modifies the VDR protein structure through substitution nucleotides of the start codon, potentially impacting its interaction with nuclear co-regulators and altering transcriptional activity of vitamin D-responsive genes (Alimirah, Peng, Murillo, & Mehta, 2011; Li et al., 2012). These alterations can affect pathways linked to immune regulation, beta-cell function, and insulin sensitivity, contributing to diabetes susceptibility. Studies exploring the direct impact of FokI on these pathways, particularly in the context of immune cell behavior and glucose metabolism, (Surendar Aravindhan et al., 2021; van Etten et al., 2007; Zhai et al., 2020a)

Despite extensive research, the association between FokI and T1DM presents varied results across populations. While Shahmoradi et al. (2021) found no overall association (Shahmoradi et al., 2021), Zhai et al. (2020) reported a protective effect of the T allele in European cohorts and an increased risk in African populations (Zhai et al., 2020a). These discrepancies may be attributed to differences in study designs, sample sizes, and regional factors such as UV exposure and dietary vitamin D intake. Future studies should standardize methodologies and include broader sample sizes to better understand these associations, controlling for confounding variables such as lifestyle and socioeconomic status.

For T2DM, which is characterized by insulin resistance and beta-cell dysfunction, significant associations with FokI have been reported, particularly in Asian populations where the T allele is linked to increased risk (S Aravindhan et al., 2021). Contrary to previous findings, our results indicate a significant association between the presence of the T allele and a reduced risk of T2DM. This discrepancy highlights the need for additional studies to further explore the relationship between specific alleles and T2DM risk. Genetic predispositions may be enhanced by environmental and lifestyle factors, such as dietary habits and physical activity levels. Examining the role of FokI in combination with other polymorphisms and metabolic risk factors would help clarify its contribution to T2DM susceptibility. Longitudinal studies and genome-wide interaction analyses could provide further insights into these complex relationships.

In GDM, Liu (2021) found FokI significantly associated with increased risk, particularly in Caucasian populations (S. Liu, 2021a). This may relate to vitamin D’s critical role in insulin secretion and beta-cell function during pregnancy potentially modified by hormonal changes. In our study, we observed that, within the African population, the allelic, dominant, homozygous, and heterozygous models are associated with a lower risk of GDM. In contrast, in the Asian population, the allelic model is associated with an increased risk of GDM. Future research should focus on the interplay between FokI and pregnancy-specific factors to better understand its impact on GDM. Concrete examples, such as intervention trials assessing vitamin D supplementation based on FokI genotypes, could inform personalized prevention strategies.

### Association between TaqI (rs731236) polymorphism and Type 1, Type 2, and GDM

The second key finding in the present study showed that TaqI (rs731236) polymorphism has no association between TaqI and the risk of T1DM when all ethnic groups are combined. However, subgroup analysis detected that recessive model (CC vs. CT+TT) and homozygous (CC vs. TT) in south American populations are significantly different among T1DM and controls suggesting an associated with increasing risk to T1DM. Likewise in T2DM, these models namely dominant model (CC+CT vs. TT) and heterozygous (CT vs. TT) are associated with increasing risk to T2DM in African population. No significant difference in all genetic models of TaqI SNPs between GDM patients and controls when all ethnicities are combined suggesting no association between this SNPs and the risk of GDM. It should be noted that we couldn’t perform subgroups analysis due to lack of studies for each ethnicity.

The TaqI (rs731236) polymorphism, located in the 3’ UTR of the VDR gene, affects mRNA stability and VDR expression, which can influence immune responses and insulin signaling (Salehizadeh et al., 2024; Zhai et al., 2020a). While Zhai et al. found no significant association with T1DM (Zhai et al., 2020a), population-specific data suggest that TaqI may interact with other immune-related genes, such as HLA-DQ (Cullen, Middleton, & Savage, 1990). This supports the need for multi-gene analyses to elucidate its potential role in T1DM risk. Ethnic variability, particularly trends indicating increased risk in South American cohorts, highlights the importance of exploring region-specific environmental interactions, including sun exposure and dietary habits (Iscovich, 1998).

Aravindhan et al. found in T2DM, TaqI does not show a strong association overall (Surendar Aravindhan et al., 2021), though studies in African populations have linked certain TaqI genotypes to increased risk (S. Z. AL-Darraji, H. F. AL-Azzawie, & A. M. AL-Kharsani, 2017). Integrating findings from gene-environment interaction studies and epigenetic analyses could reveal how TaqI modulates glucose homeostasis under different environmental conditions. However, no significant associations were found with the TaqI polymorphism for these complications (Song et al., 2019). Additionally, Liu et al, found no significant overall association between GDM and TaqI, although pregnancy-related interactions between TaqI and maternal vitamin D levels warrant further study (Apaydın et al., 2019b; S. Liu, 2021b). Larger, multi-ethnic cohort studies that include vitamin D status and hormonal profiles (Fu et al., 2024).

### BsmI (rs1544410) polymorphism in Type 1, Type 2, and Gestational Diabetes Mellitus

The third major finding of the current study indicates that BsmI SNPs under overdominant model (GA vs. GG+AA) and (GG vs. GA) models when all ethnicities are combined significantly different among T1DM and controls suggesting an increasing risk for T1DM when (GA vs. GG+AA) is present and decreasing risk for T1DM when (GG vs. GA) model is present. However, group analyses based on ethnicity show that the increased frequency of allelic (G vs. A) and homozygous (GG vs. AA) model in African population may be associated with increased risk to T1DM. Likewise increasing frequency of dominant model (GG+GA vs. AA) and heterozygous (GA vs. AA) gene models in European population and overdominant (GA vs. GG+AA) model in Asian population maybe increasing risk to T1DM. However, in south American population, the increasing frequency of allelic (G vs. A), recessive model (GG vs. GA+AA), dominant model (GG+GA vs. AA), homozygous (GG vs. AA) and heterozygous (GA vs. AA) model may be associated with decreased risk to T1DM.

The BsmI (rs1544410) polymorphism, also located in the 3’ UTR, is believed to impact VDR expression and consequently affect immune and metabolic pathways. While Shahmoradi et al. observed no overall association with T1DM, specific genetic models suggested nuanced risk variations (Zhai et al., 2020b). The observed lowered susceptibility to T1DM in American cohorts versus inconsistent results in European and Asian groups could stem from genetic linkage differences and varying environmental exposures. Including visual data representation, such as allele frequency distributions and region-specific odds ratios, would enhance interpretation and applicability of these findings (Ji, Hemminki, Sundquist, & Sundquist, 2010).

Aravindhan et al. indicates that in T2DM, BsmI has been associated with risk under heterozygote model (Surendar Aravindhan et al., 2021). Future research should focus on how BsmI modulates inflammatory pathways, as chronic inflammation is a known contributor to T2DM. Functional studies exploring BsmI’s role in immune cell regulation and insulin signaling could offer valuable insights. For GDM, regional differences in BsmI’s effect have been noted, with increased risk observed in specific populations (S. Liu, 2021b). Investigating the interaction between BsmI, maternal vitamin D levels, and pregnancy-related hormonal changes would clarify its role in GDM. Including environmental data, such as sun exposure and dietary vitamin D intake, could further contextualize these findings.

### ApaI (rs7975232) polymorphism in Type 1, Type 2, and Gestational Diabetes Mellitus

The fourth major finding of the present study shows that ApaI (rs7975232) polymorphism under allelic (G vs. T), recessive model (GG vs. GT+TT) and homozygous (GG vs. TT) models are significantly increased in T1DM compared to controls which associated with decreased risk to T1DM in overall genetic model where the ethnicities are combined. Subgroup analysis showed no significant differences among different ethnicities. In T2DM, there is no significant difference in all models when all ethnicities are combined compared to controls. However, subgroup analysis showed that in African population, the recessive model (GG vs. GT+TT) significantly different between T2DM and controls and associated with decreased risk to T2DM. In GDM, there is no significant differences in all gene models.

The ApaI (rs7975232) polymorphism, located in intron 8, influences VDR expression, impacting immune function and glucose metabolism. While Shahmoradi et al. reported no overall association with T1DM, allelic models showed protective effects, particularly in African cohorts (Shahmoradi et al., 2021). Functional genomic studies should explore how ApaI modulates VDR gene expression under different environmental conditions. For T2DM, potential protective effects of ApaI in certain regions have been reported (Surendar Aravindhan et al., 2021), possibly due to its role in enhancing VDR expression. Research focusing on cumulative effects of ApaI with other polymorphisms could elucidate how it impacts metabolic pathways. For GDM, Liu et al highlighted an increased risk linked to ApaI in specific populations (S. Liu, 2021a). Longitudinal studies that include pregnancy-related variables and epigenetic factors would help determine how these interactions influence GDM risk.

Clinical and research implications genotyping for VDR polymorphisms, particularly FokI, TaqI, BsmI, and ApaI, could suggest personalized diabetes prevention and treatment strategies. Integrating genetic data with environmental factors in multi-gene panels could offer a comprehensive view of diabetes risk. Standardizing study designs and including diverse populations with controlled confounding factors will be crucial for advancing this research. Implementing visual tools, such as allele distribution charts and regional odds ratio plots, would improve data interpretation and clinical applicability. The understanding of VDR polymorphisms as part of a complex genetic landscape influencing diabetes highlights the importance of multifactorial approaches. Combining genetic, environmental, and epigenetic data in future studies could bridge research findings with public health strategies and personalized medical practices.

## Limitations

This study’s findings are limited by data heterogeneity, inconsistent control of confounding variables, and reliance on cross-sectional and case-control designs, which hinder causal inferences. The lack of functional studies and limited representation of diverse populations further restricts generalizability. Future research should prioritize longitudinal studies, diverse cohorts, and integrative approaches to better understand the role of VDR polymorphisms in diabetes.

## Conclusions

As summarized in **Figure 9**, this study identifies key associations between VDR gene polymorphisms and diabetes susceptibility. The G allele of BsmI significantly increases T2DM risk, while the T allele of FokI provides protection against T1DM. These associations vary across populations, reflecting the interplay of genetic and environmental factors. The findings underscore the need for standardized methodologies and large, multi-ethnic studies to address inconsistencies. Future research should prioritize functional analyses to elucidate mechanisms and integrate genetic and environmental data to develop personalized prevention and treatment strategies.

**Figure 9.**
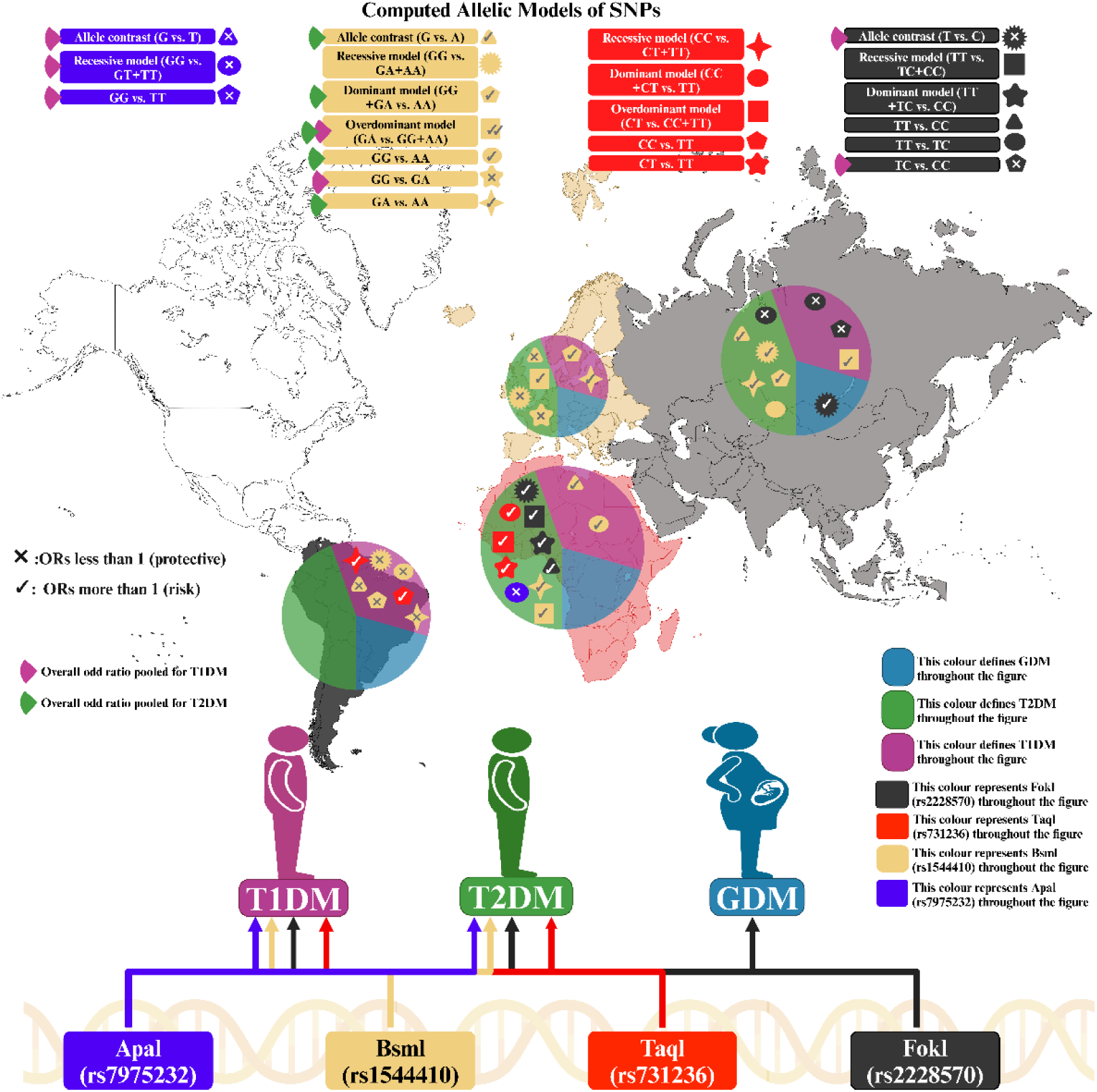
Summary of the associations of four VDR gene polymorphisms (FokI, TaqI, BsmI, and ApaI) with type 1 diabetes mellitus (T1DM), type 2 diabetes mellitus (T2DM), and gestational diabetes mellitus (GDM) across different genetic models and ethnic groups.

## Data Availability

The corresponding author (AA) will consider reasonable requests for access to the dataset (Excel file) utilized in this meta-analysis. Access will be granted after all authors have completed their planned analyses and fully utilized the data.

## Ethical approval and consent to participate

Not applicable.

## Consent for publication

Not applicable.

## Author’s contributions

HAM and AFA developed the study design, with HAM, AKA, HAA, and AFA working together to collect the data. HAM and AFA carried out the statistical analysis. All authors collaborated in writing the paper and gave their full approval for submitting the final version.

## Acknowledgments

Not applicable.

