## supplementary file 1 for "Vitamin D Receptor Gene Polymorphisms in Type 1, Type 2, and Gestational Diabetes Mellitus: A Comprehensive Meta-Analysis and Meta-Regression of 154 Studies"

**S1, Table 1.** Search sentences and terms were used in each database.

| **Database Name** | **Search Sentence** | **No. of Articles** |
| --- | --- | --- |
| **PubMed/Medline** | "FokI polymorphism" AND "Vitamin D receptor" AND ("Type 1 Diabetes Mellitus" OR "T1DM" OR "Type 2 Diabetes Mellitus" OR "T2DM" OR "Gestational Diabetes Mellitus" OR "GDM") | **29** |
|  | "BsmI polymorphism" AND "Vitamin D receptor" AND ("Type 1 Diabetes Mellitus" OR "T1DM" OR "Type 2 Diabetes Mellitus" OR "T2DM" OR "Gestational Diabetes Mellitus" OR "GDM") | **17** |
|  | "TaqI polymorphism" AND "Vitamin D receptor" AND ("Type 1 Diabetes Mellitus" OR "T1DM" OR "Type 2 Diabetes Mellitus" OR "T2DM" OR "Gestational Diabetes Mellitus" OR "GDM") | **8** |
|  | "ApaI polymorphism" AND "Vitamin D receptor" AND ("Type 1 Diabetes Mellitus" OR "T1DM" OR "Type 2 Diabetes Mellitus" OR "T2DM" OR "Gestational Diabetes Mellitus" OR "GDM") | **7** |
|  | "Vitamin D receptor" AND ("FokI" OR "BsmI" OR "TaqI" OR "ApaI") AND ("Type 1 Diabetes Mellitus" OR "T1DM" OR "Type 2 Diabetes Mellitus" OR "T2DM" OR "Gestational Diabetes Mellitus" OR "GDM") | **137** |
| **Google Scholar** | "FokI polymorphism" AND "Vitamin D receptor" AND ("Type 1 Diabetes" OR "T1DM" OR "Type 2 Diabetes" OR "T2DM" OR "Gestational Diabetes" OR "GDM") | **1200** |
|  | "BsmI polymorphism" AND "Vitamin D receptor" AND ("Type 1 Diabetes" OR "T1DM" OR "Type 2 Diabetes" OR "T2DM" OR "Gestational Diabetes" OR "GDM") | **1020** |
|  | "TaqI polymorphism" AND "Vitamin D receptor" AND ("Type 1 Diabetes" OR "T1DM" OR "Type 2 Diabetes" OR "T2DM" OR "Gestational Diabetes" OR "GDM") | **665** |
|  | "ApaI polymorphism" AND "Vitamin D receptor" AND ("Type 1 Diabetes" OR "T1DM" OR "Type 2 Diabetes" OR "T2DM" OR "Gestational Diabetes" OR "GDM") | **420** |
|  | "Vitamin D receptor polymorphism" AND ("FokI" OR "BsmI" OR "TaqI" OR "ApaI") AND ("Type 1 Diabetes" OR "T1DM" OR "Type 2 Diabetes" OR "T2DM" OR "Gestational Diabetes" OR "GDM") | **845** |
| **SciFinder** | FokI polymorphism AND Vitamin D receptor AND ("Type 1 Diabetes Mellitus" OR T1DM OR "Type 2 Diabetes Mellitus" OR T2DM OR "Gestational Diabetes Mellitus" OR GDM) | **209** |
|  | BsmI polymorphism AND Vitamin D receptor AND ("Type 1 Diabetes Mellitus" OR T1DM OR "Type 2 Diabetes Mellitus" OR T2DM OR "Gestational Diabetes Mellitus" OR GDM) | **205** |
|  | TaqI polymorphism AND Vitamin D receptor AND ("Type 1 Diabetes Mellitus" OR T1DM OR "Type 2 Diabetes Mellitus" OR T2DM OR "Gestational Diabetes Mellitus" OR GDM) | **206** |
|  | ApaI polymorphism AND Vitamin D receptor AND ("Type 1 Diabetes Mellitus" OR T1DM OR "Type 2 Diabetes Mellitus" OR T2DM OR "Gestational Diabetes Mellitus" OR GDM) | **206** |
|  | Vitamin D receptor polymorphism AND (FokI OR BsmI OR TaqI OR ApaI) AND ("Type 1 Diabetes Mellitus" OR T1DM OR "Type 2 Diabetes Mellitus" OR T2DM OR "Gestational Diabetes Mellitus" OR GDM) | **151** |

**S1, Table 2.** Newcastle-Ottawa Quality Assessment Form for Case-Control.

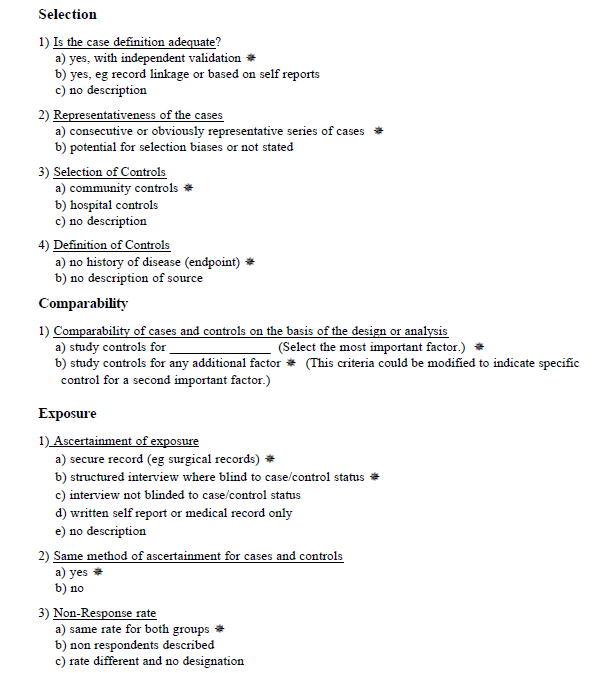

**S1, Table 3.** Results of Meta-Regression.

| **Gene Model** | **Explanatory**  **Variable** | **Q** | **df** | **p-value** | **τ²** | **I²** | **H²** | **R² (%)** | **t** | **P-value** |
| --- | --- | --- | --- | --- | --- | --- | --- | --- | --- | --- |
| **T1DM** | | | | | | | | | | |
| Allelic FokI (TC vs CC) & | Age | 66.502 | 30 | <0.001 | 0.086 | 43.8 | 1.780 | 39 | -2.772 | 0.009 |
|  | Ethnicity | 73.868 | 35 | 0.000 | 0.060 | 40.5 | 1.682 | 36 | 2.380 | 0.023 |
|  | DOI | 16.45 | 13 | 0.225 | 0.033 | 24.3 | 1.321 | 68.2 | -2.890 | 0.013 |
| Allelic ApaI G vs T & | DOI | 11.36 | 10 | 0.330 | 0.003 | 5.3 | 1.05 | 87.8 | -2.758 | 0.020 |

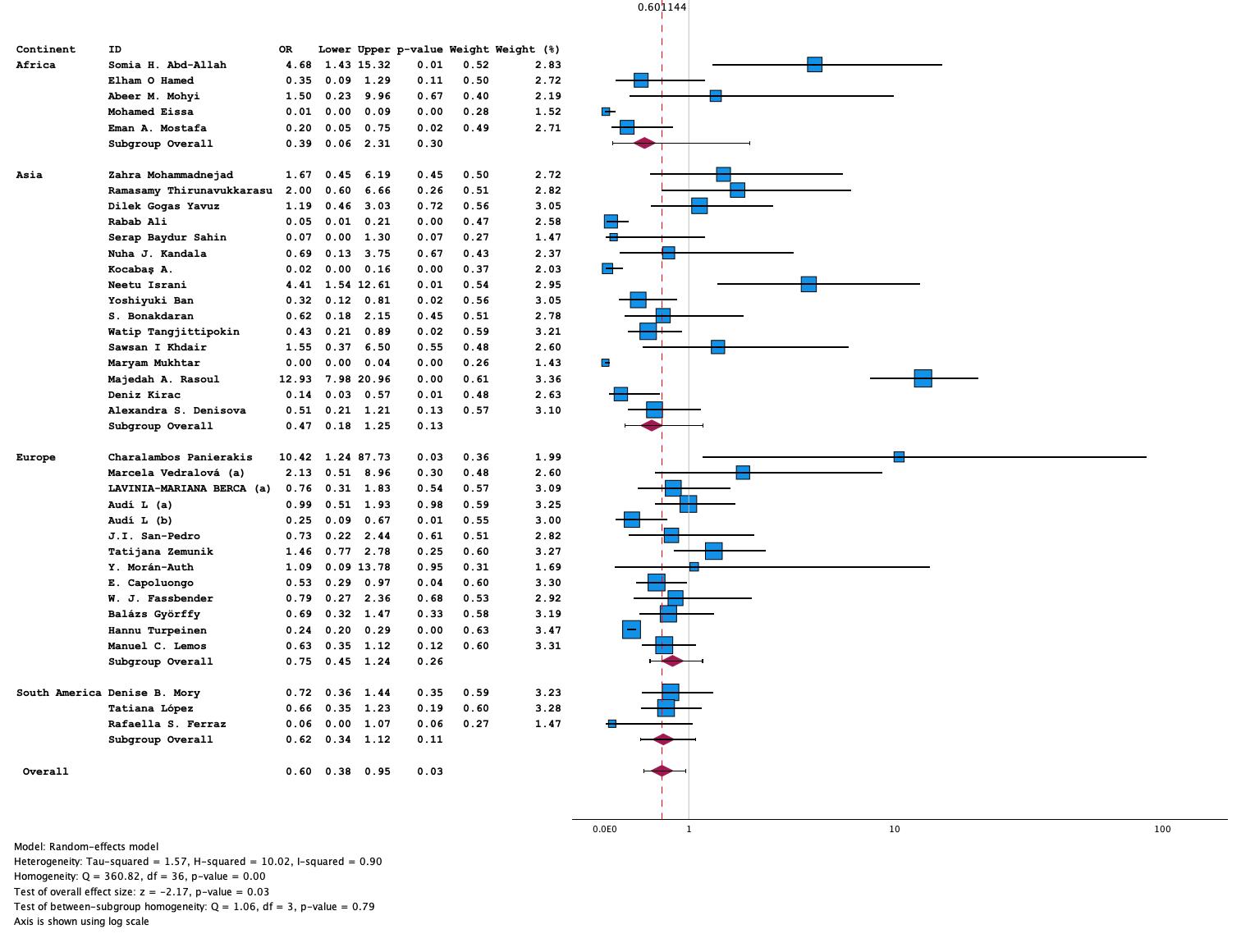
**S1. Figure 1** Forest plot of FokI SNP (TT vs TC) model in T1DM

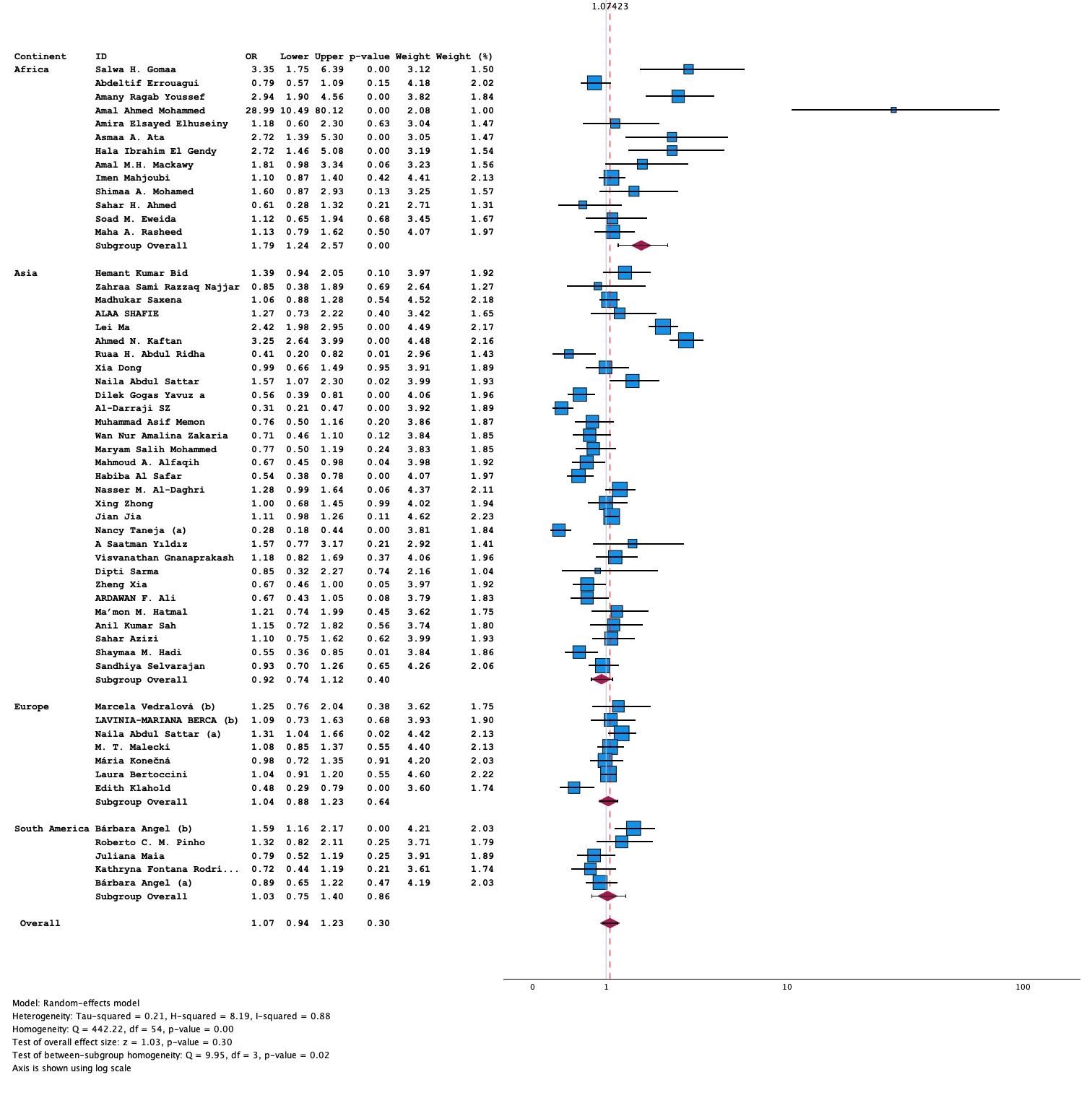
**S1. Figure 2** Forest plot of FokI SNP (T vs C) in T2DM

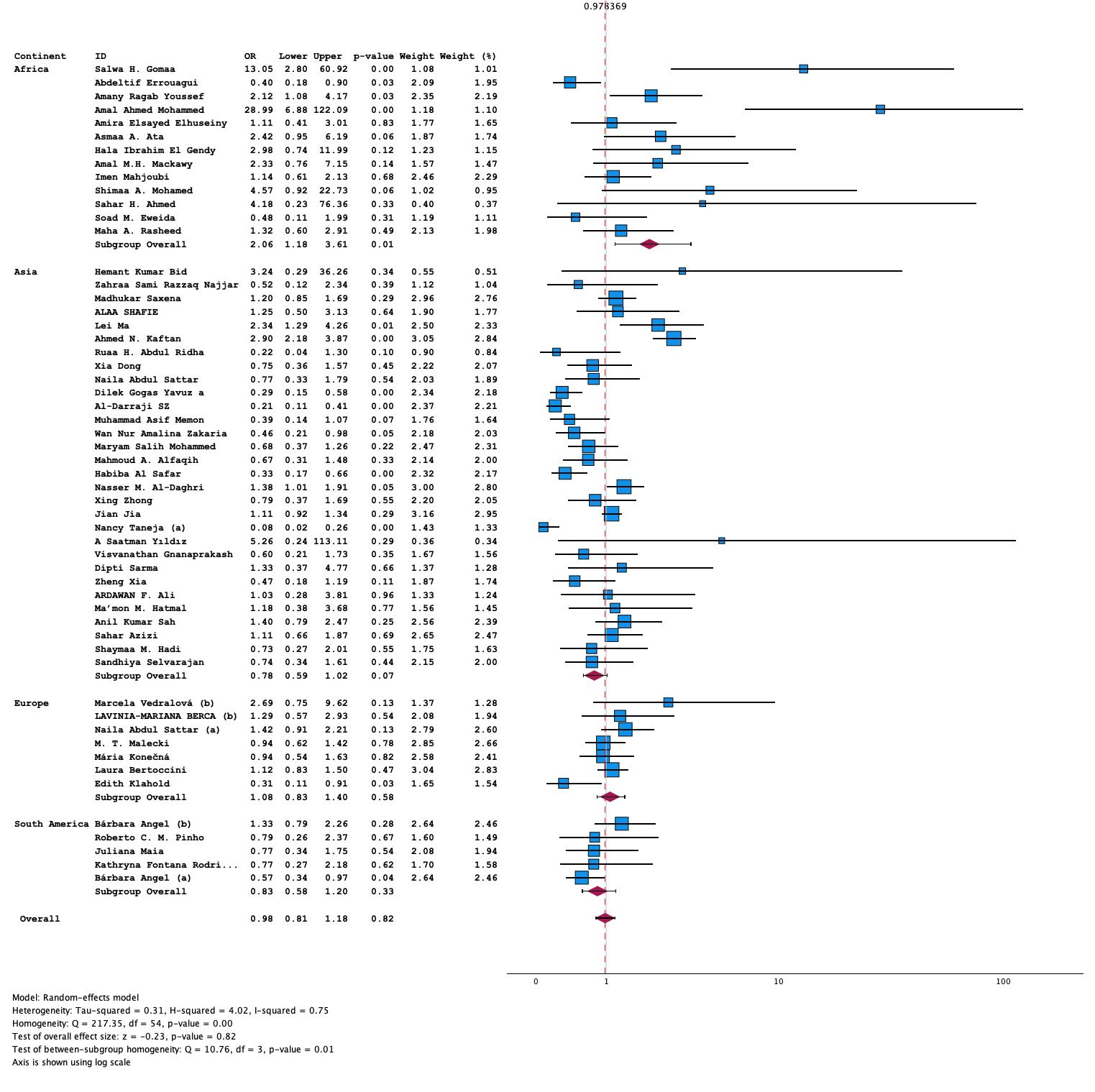
**S1. Figure 3** Forest plot of FokI SNP Recessive (TT vs. TC+CC) model in T2DM

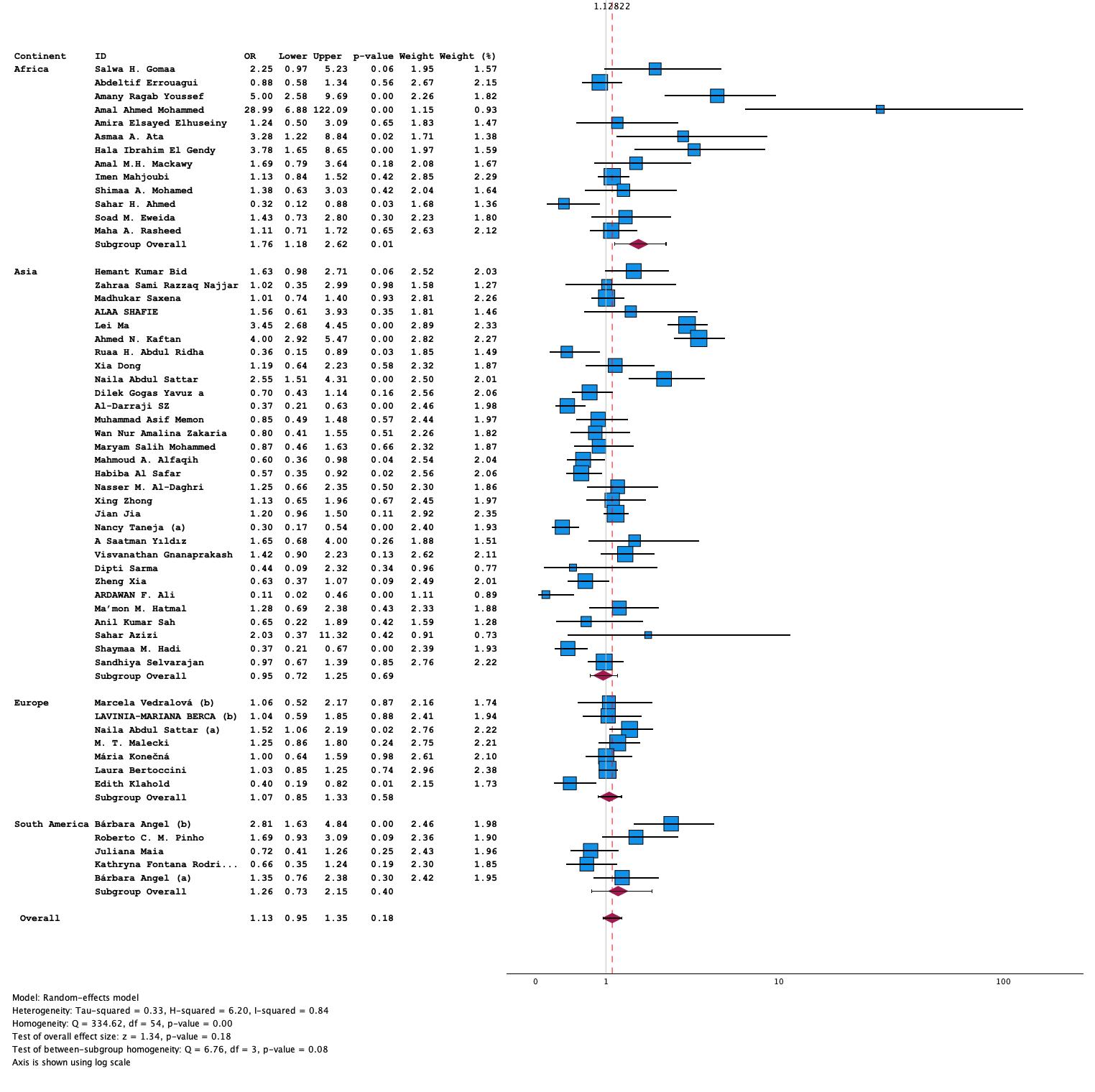
**S1. Figure 4** Forest plot of FokI SNP Dominant (TT+TC vs. CC) model in T2DM

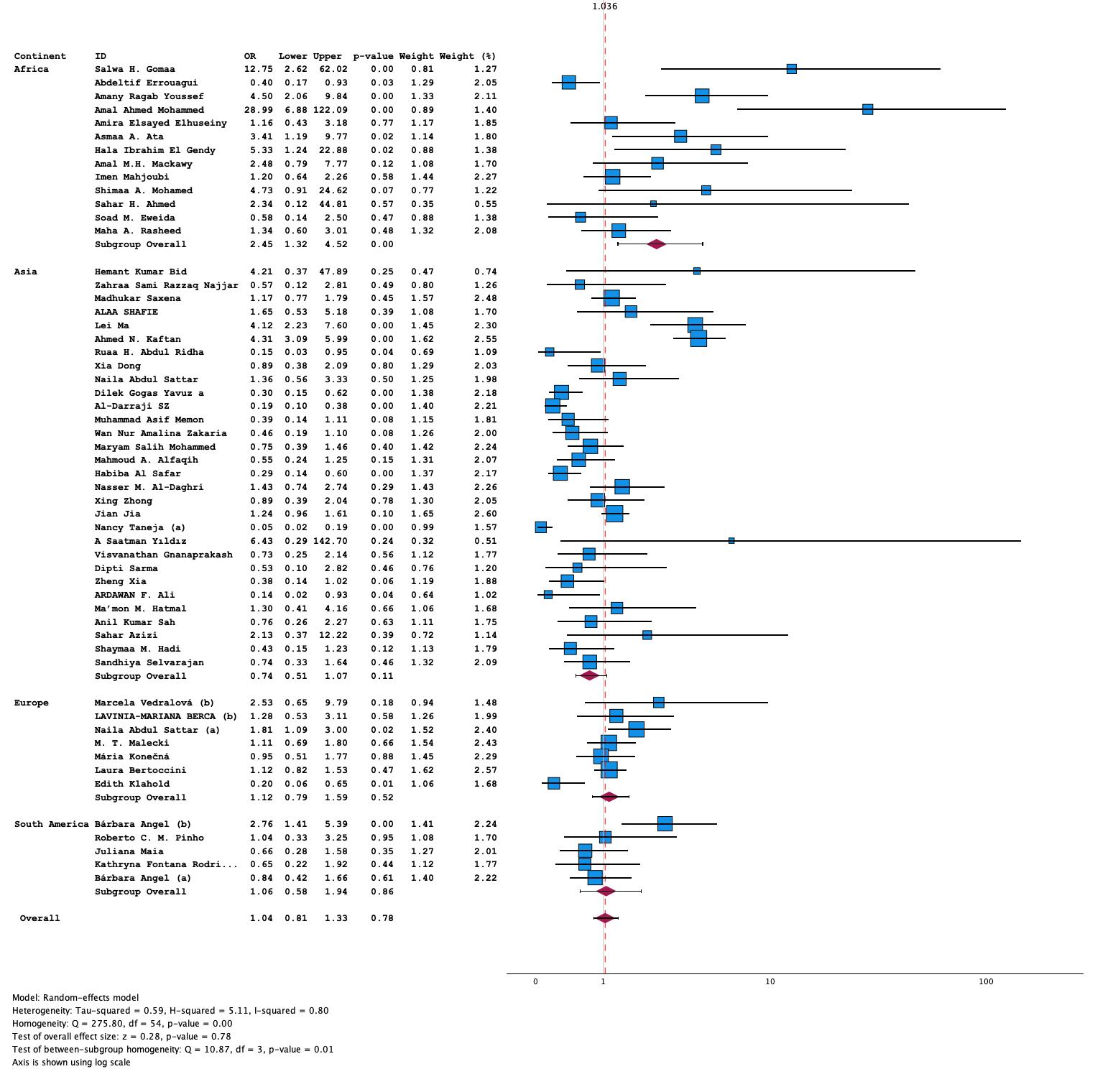
**S1. Figure 5** Forest plot of FokI SNP (TT vs CC) model in T2DM

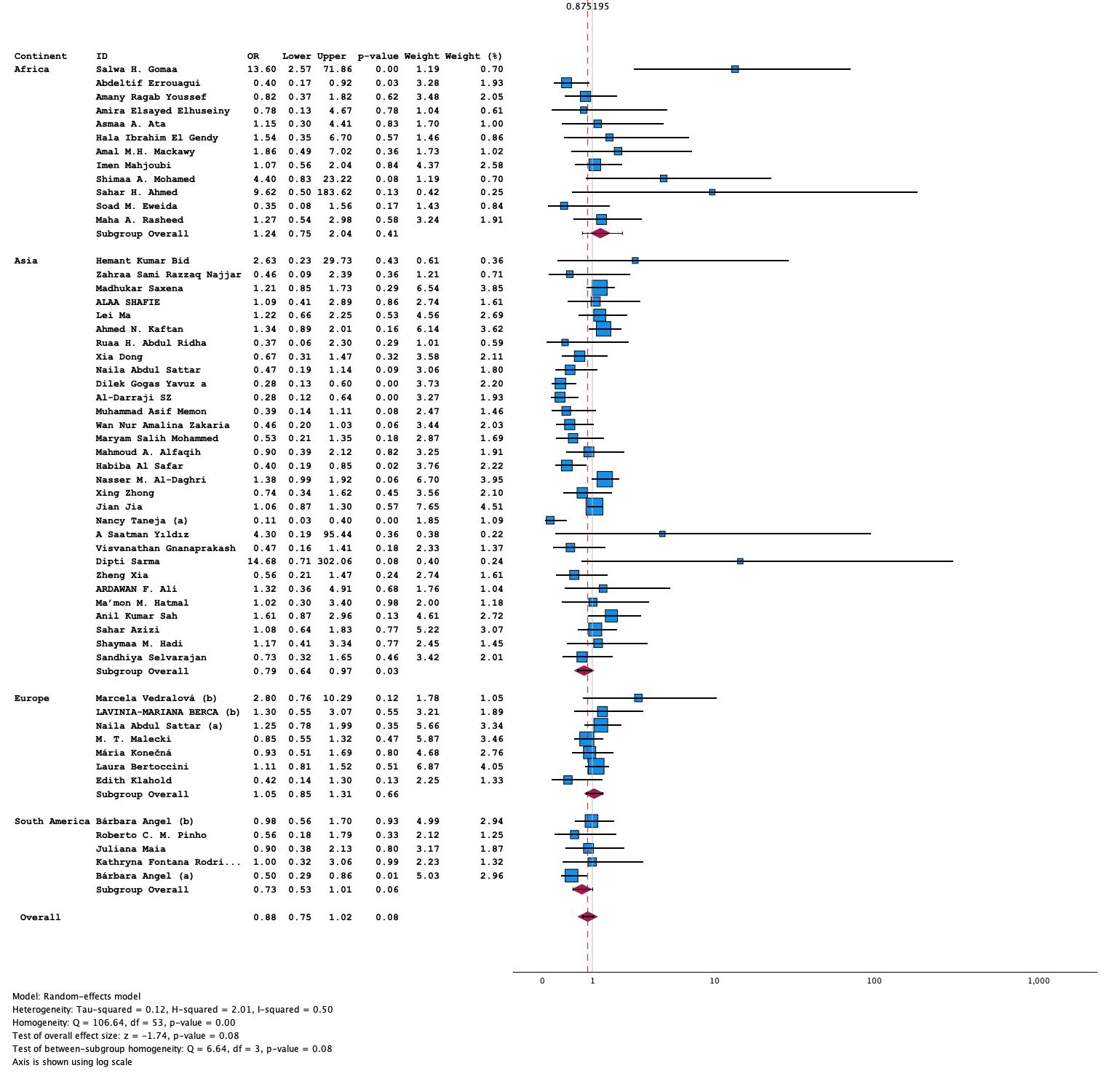
**S1. Figure 6** Forest plot of FokI SNP (TT vs TC) model in T2DM

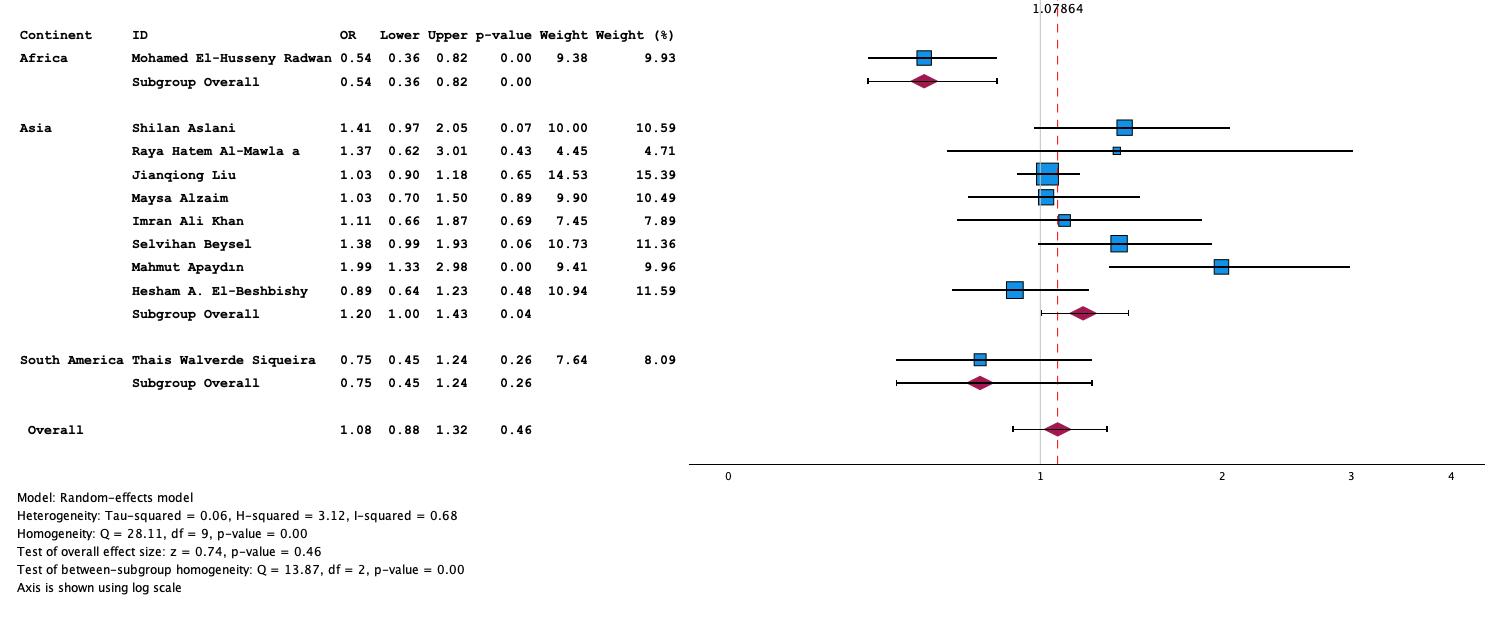

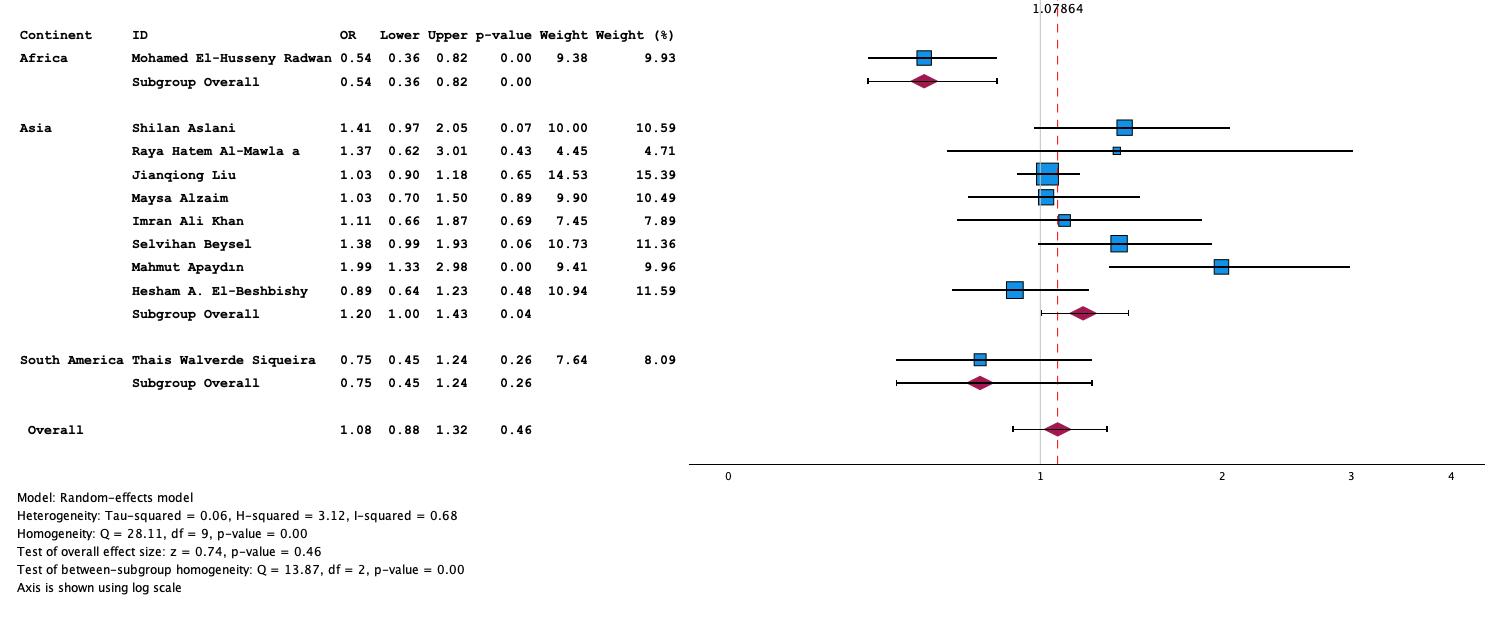

S1. Figure 7 Forest plot of FokI SNP (T vs C) model in GDM

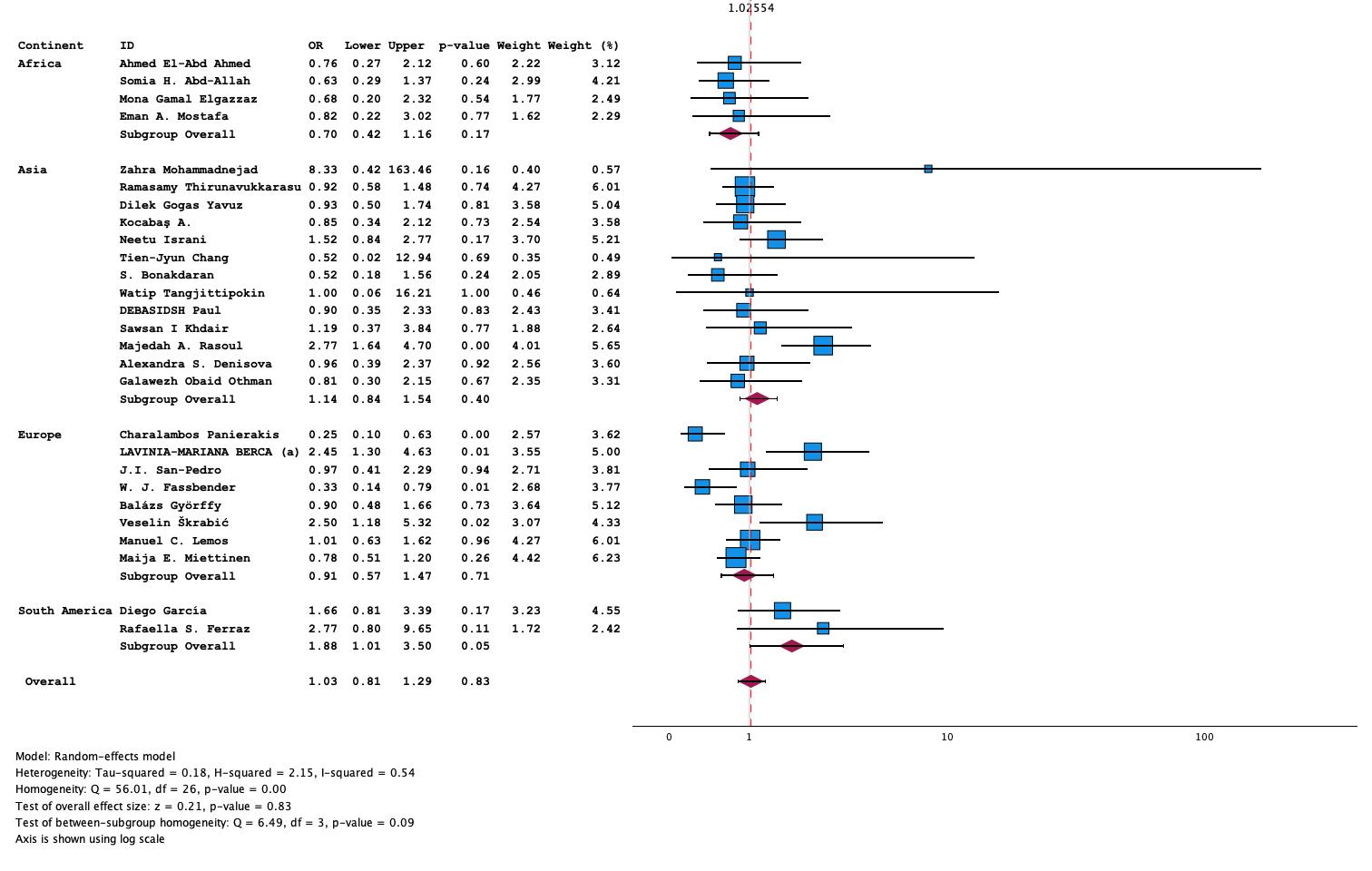

S1. Figure 8 Forest plot of TaqI SNP Recessive (CC vs. CT+TT) model in T1DM

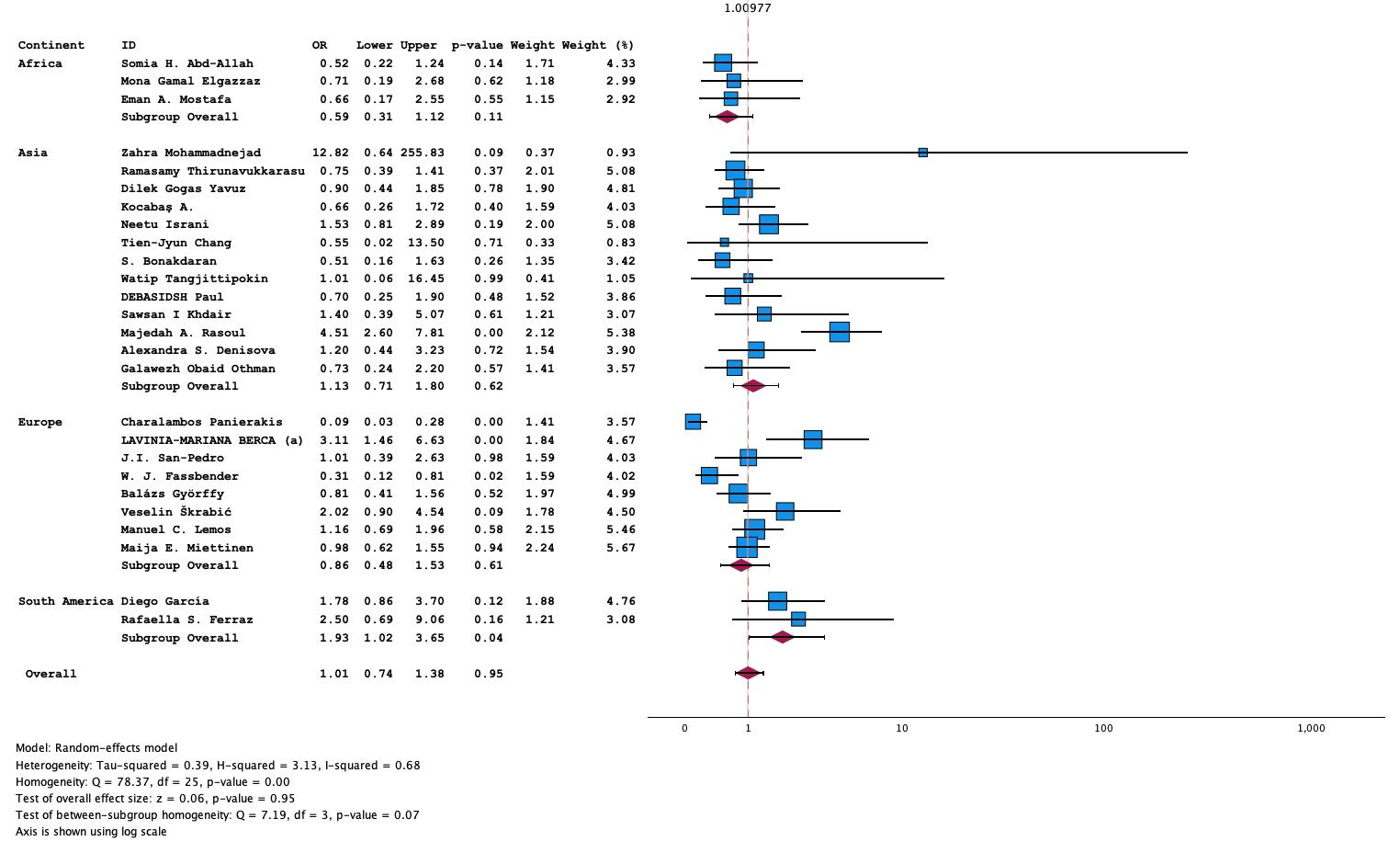
S1. Figure 9 Forest plot TaqI SNP homozygous (CC vs TT) model in T1DM

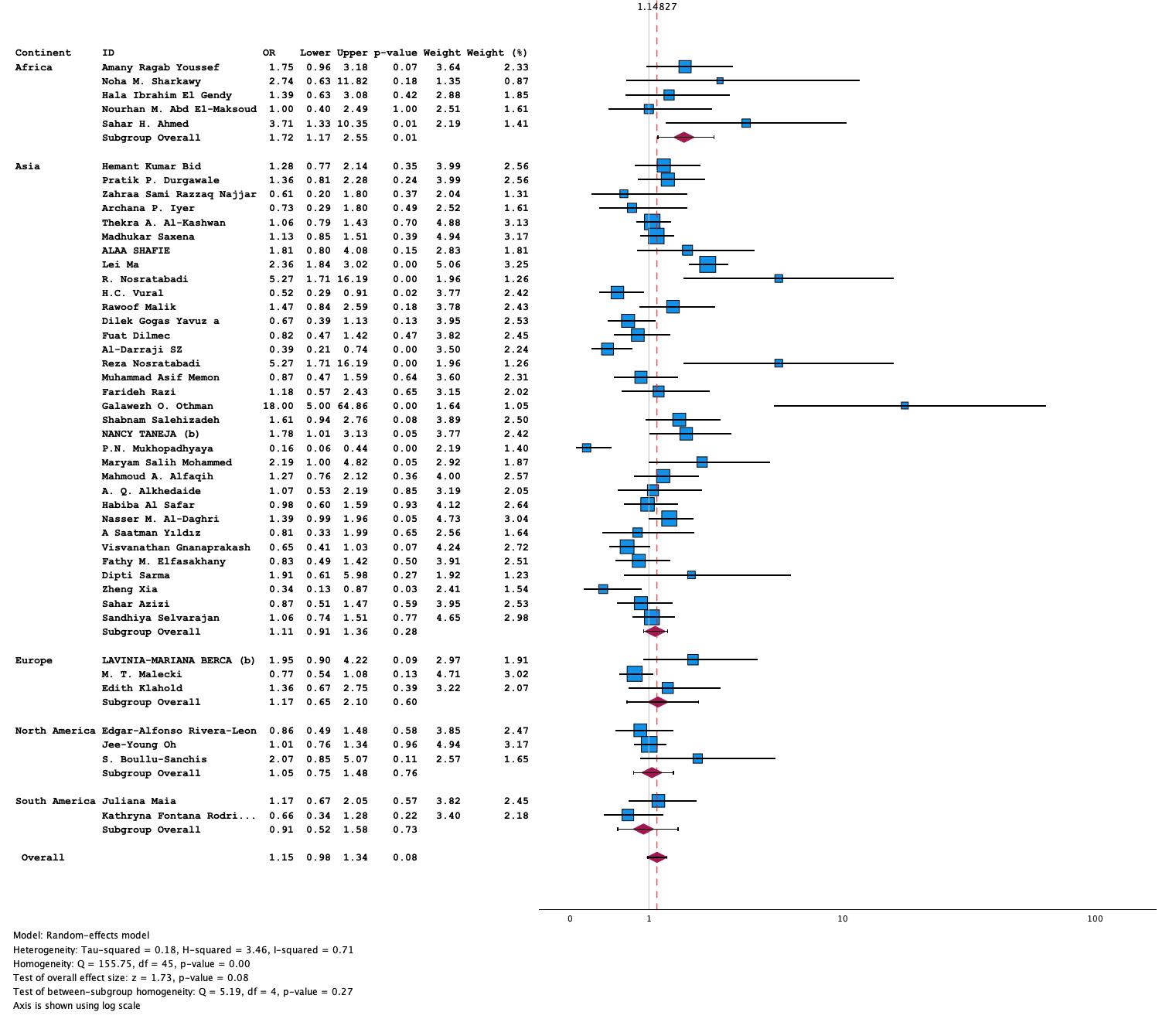

S1. Figure 10 Forest plot of TaqI SNP Dominant (CC+CT vs. TT) model in T2DM

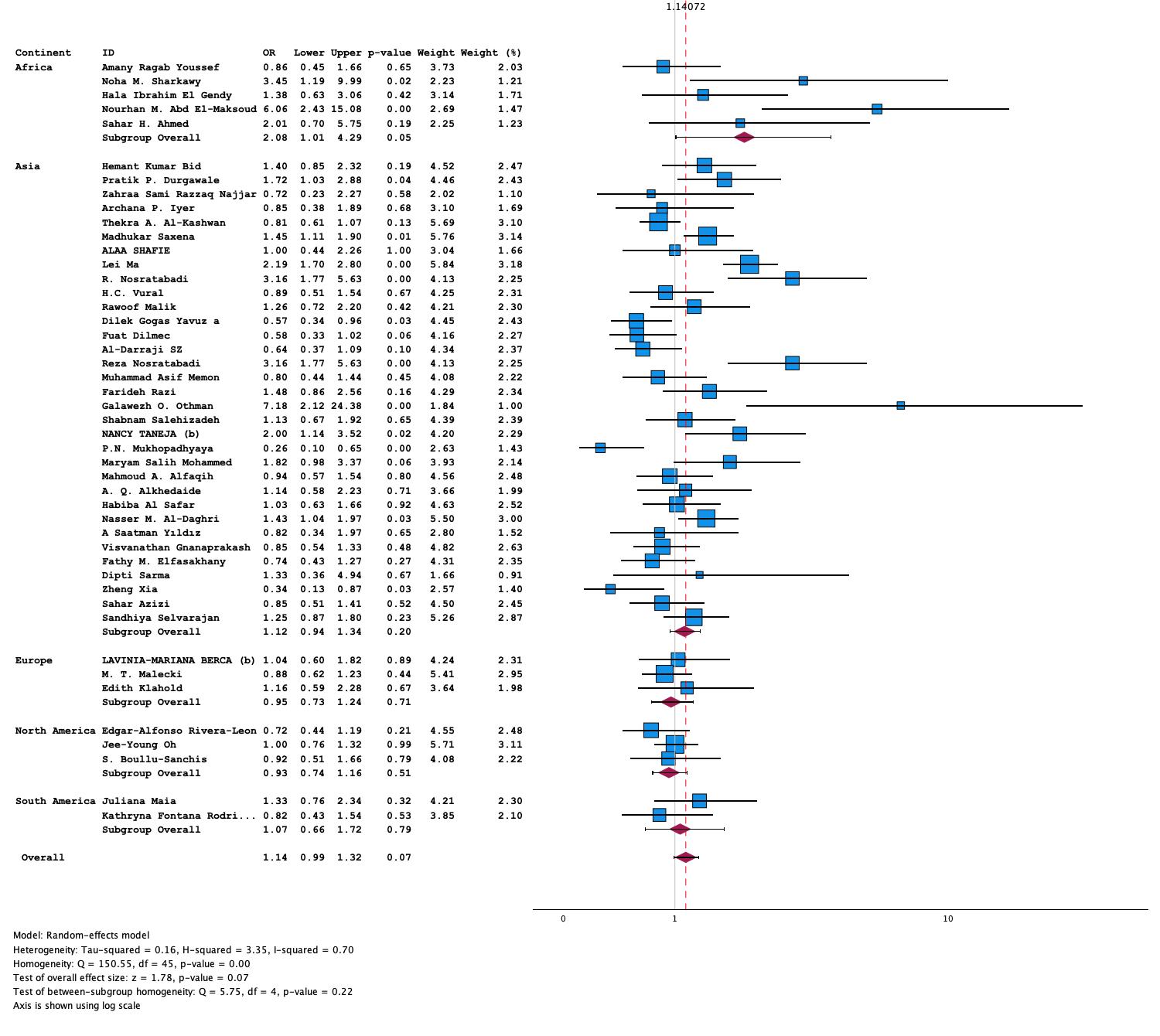
S1. Figure 11 Forest plot of TaqI OverDominant (CT vs. CC+TT) model in T2DM

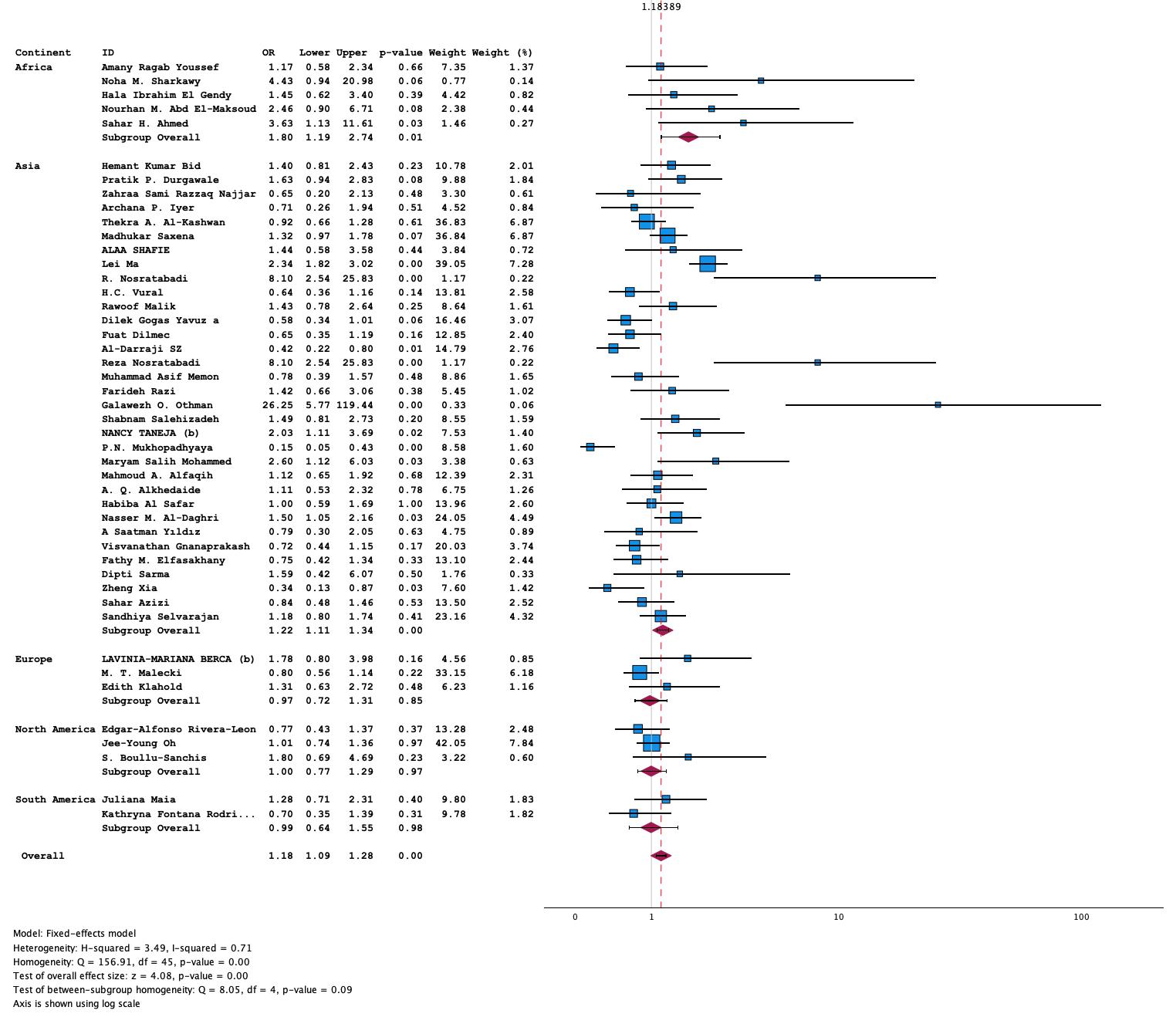

S1. Figure 12 Forest plot of TaqI heterozygous (CT vs. TT) model in T2DM

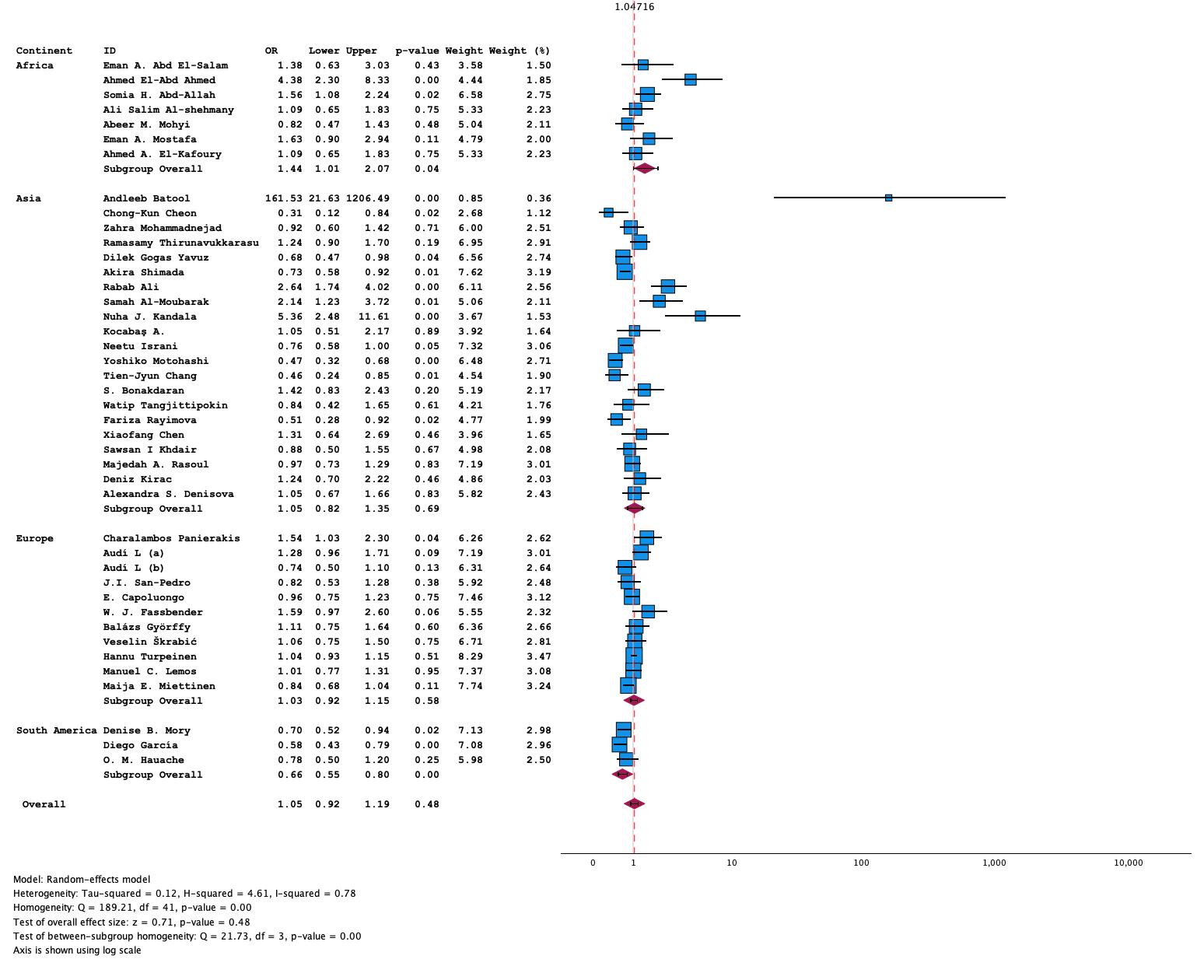

S1. Figure 13 Forest plot of BsmI SNP allelic (G vs A) model in T1DM

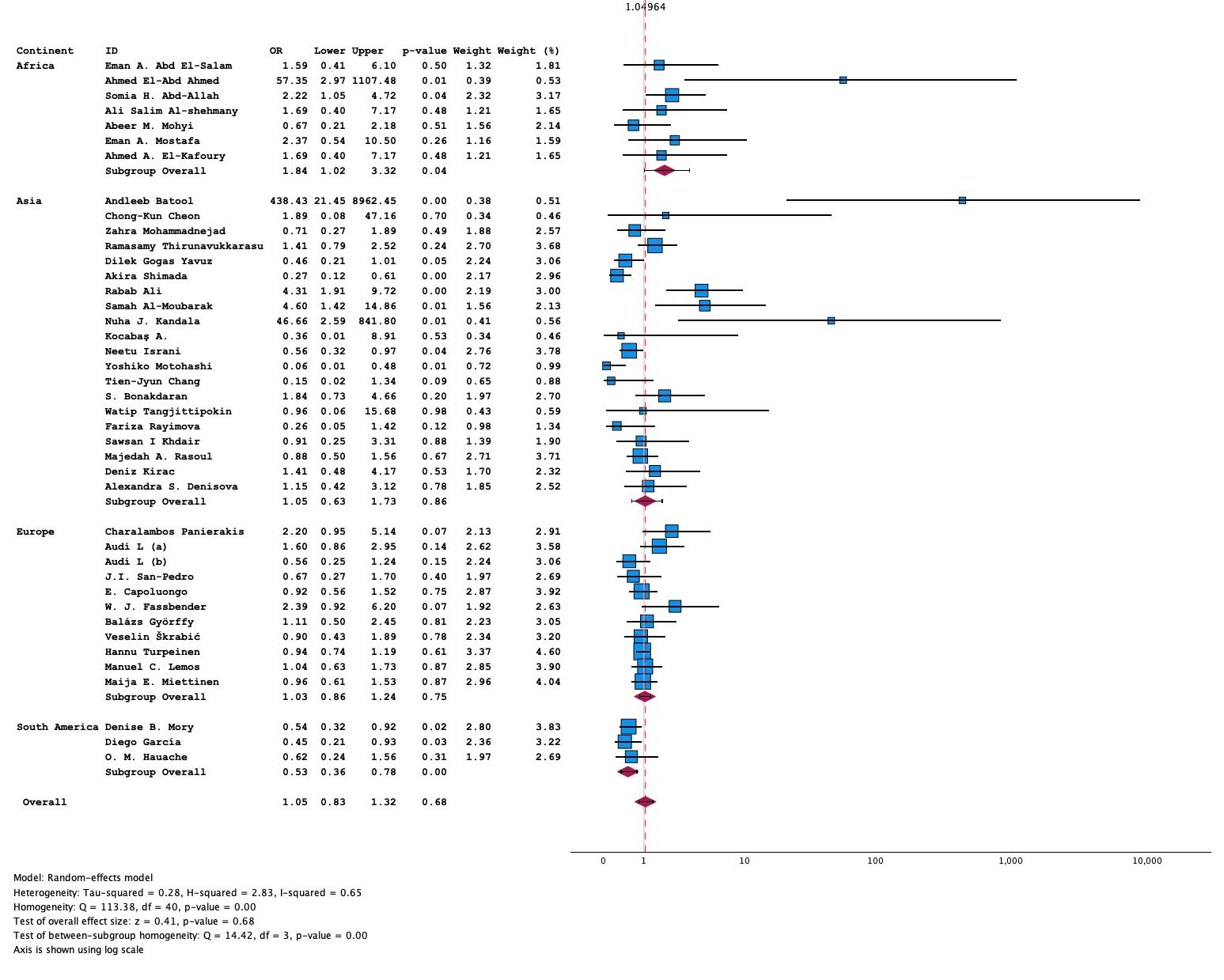
S1. Figure 14 Forest plot of BsmI Homozygous (GG vs AA) model in T1DM

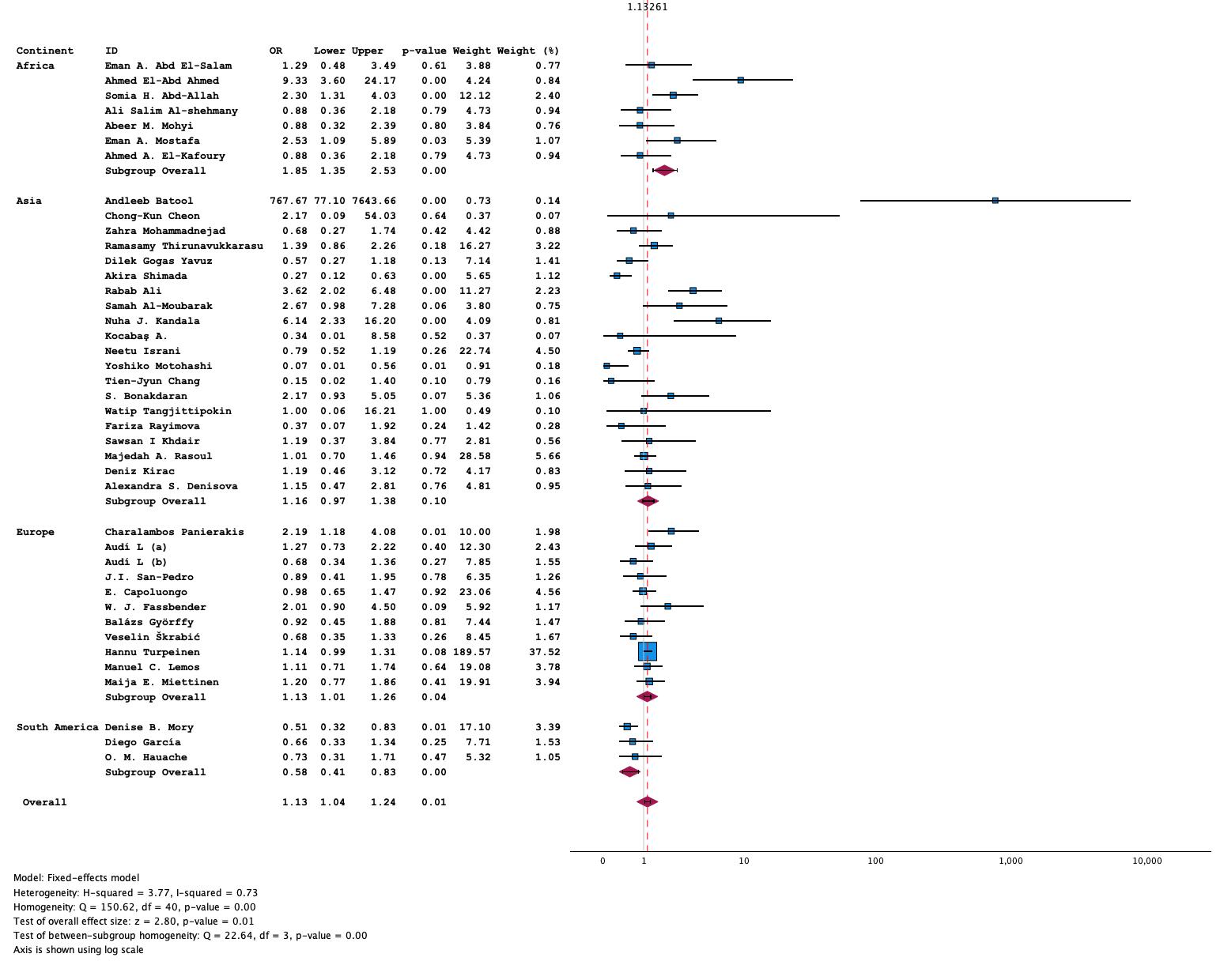
S1. Figure 15 Forest plot of BsmI SNP dominant (GG+GA vs. AA) model in T1DM

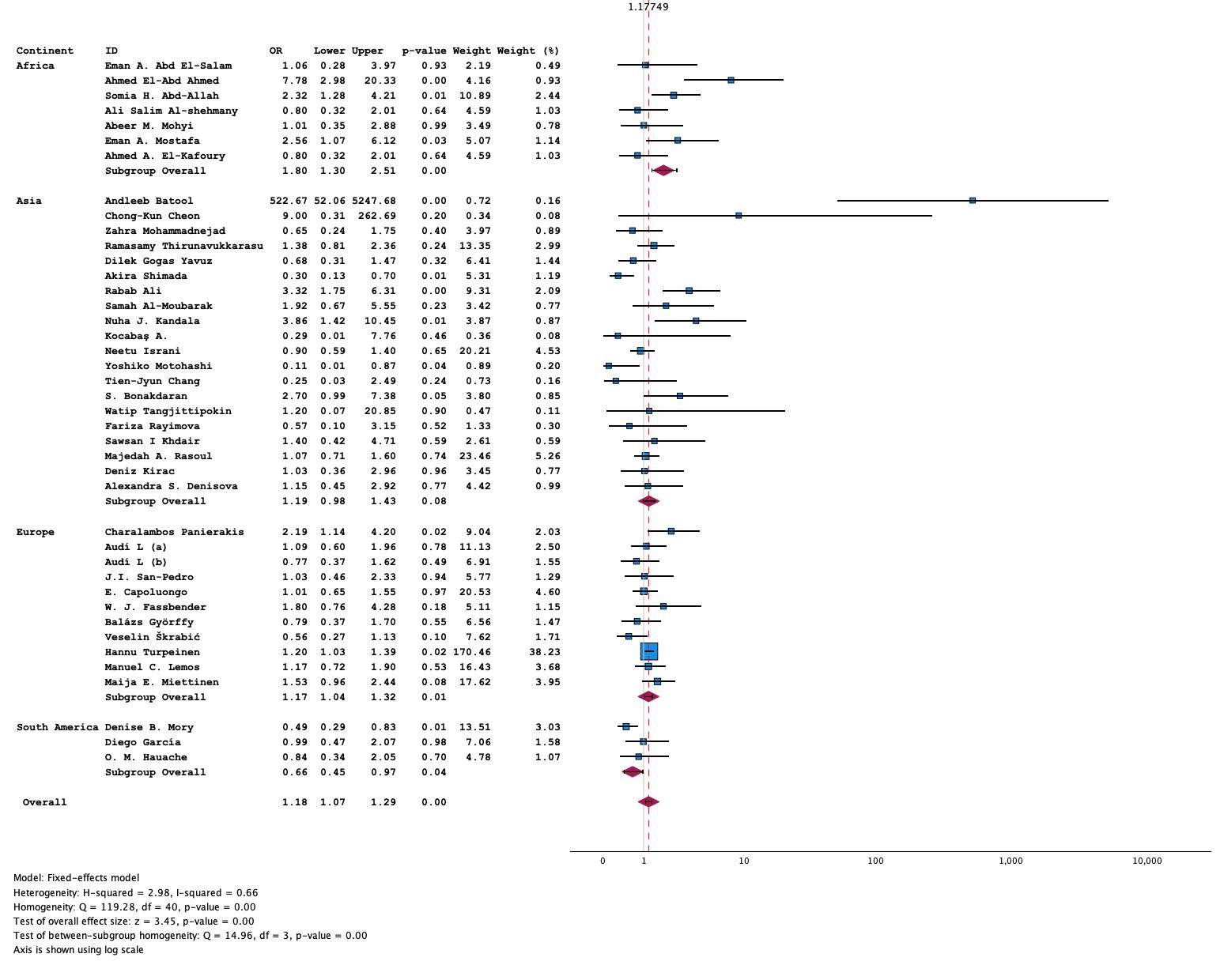

S1. Figure 16 Forest Plot of BsmI SNP heterozygous (GA vs AA) model in T1DM

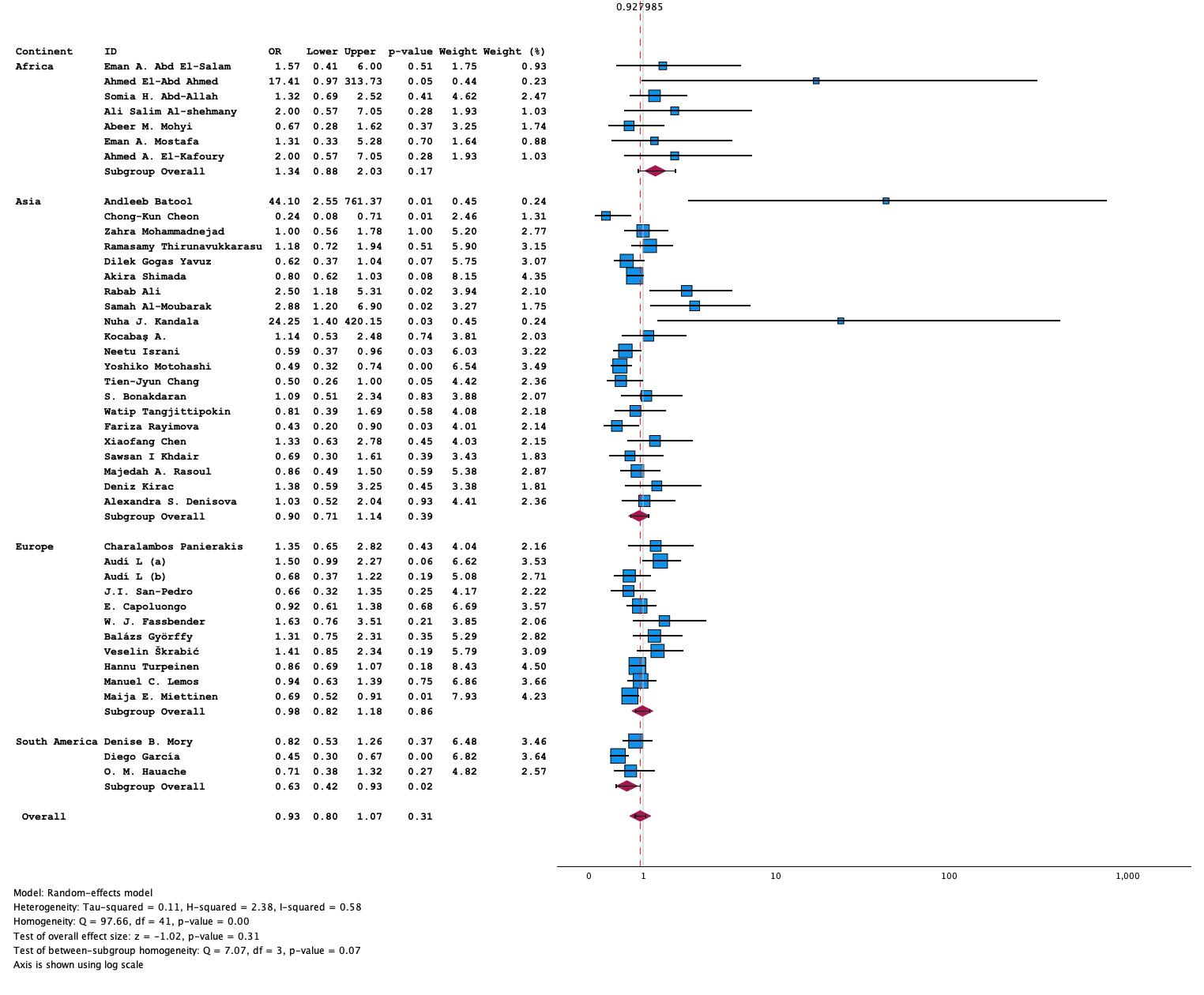
S1. Figure 17 Forest Plot of BsmI SNP Recessive (GG vs. GA+AA) model in T1DM

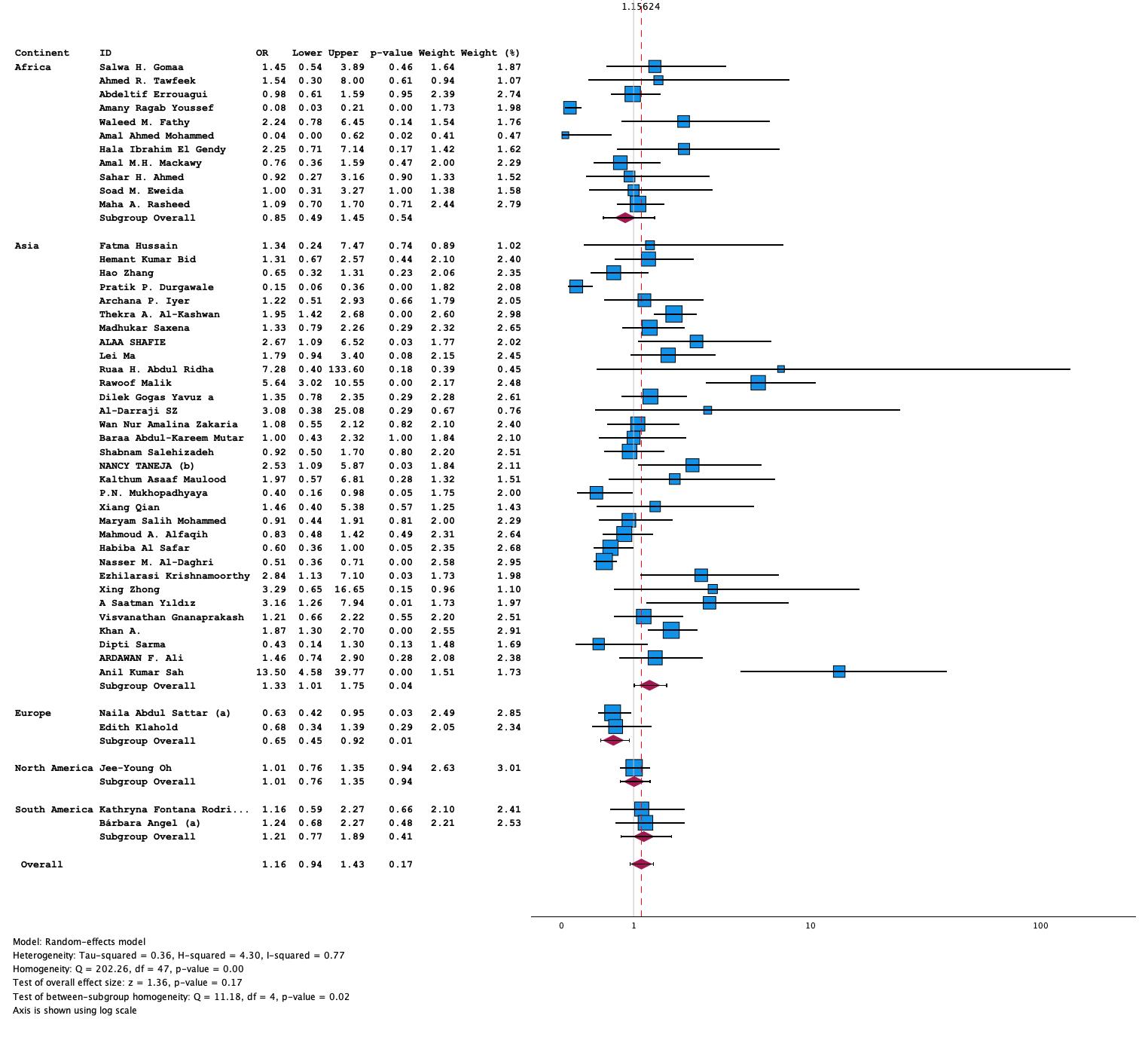
S1. Figure 18 Forest Plot of BsmI SNP Recessive (GG vs. GA+AA) in T2DM

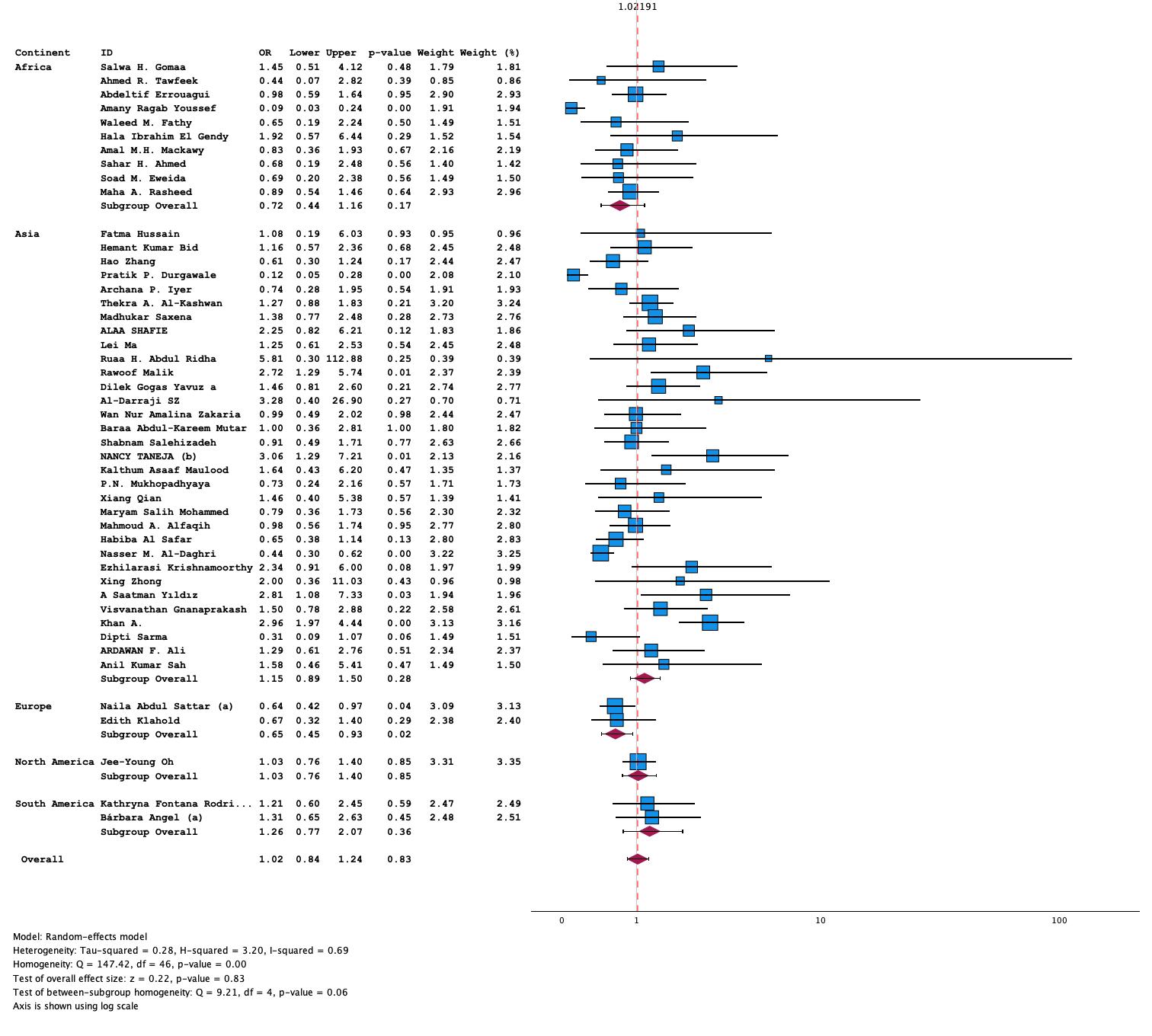
S1. Figure 19 Forest Plot BsmI SNP (GG vs GA) model in T2DM

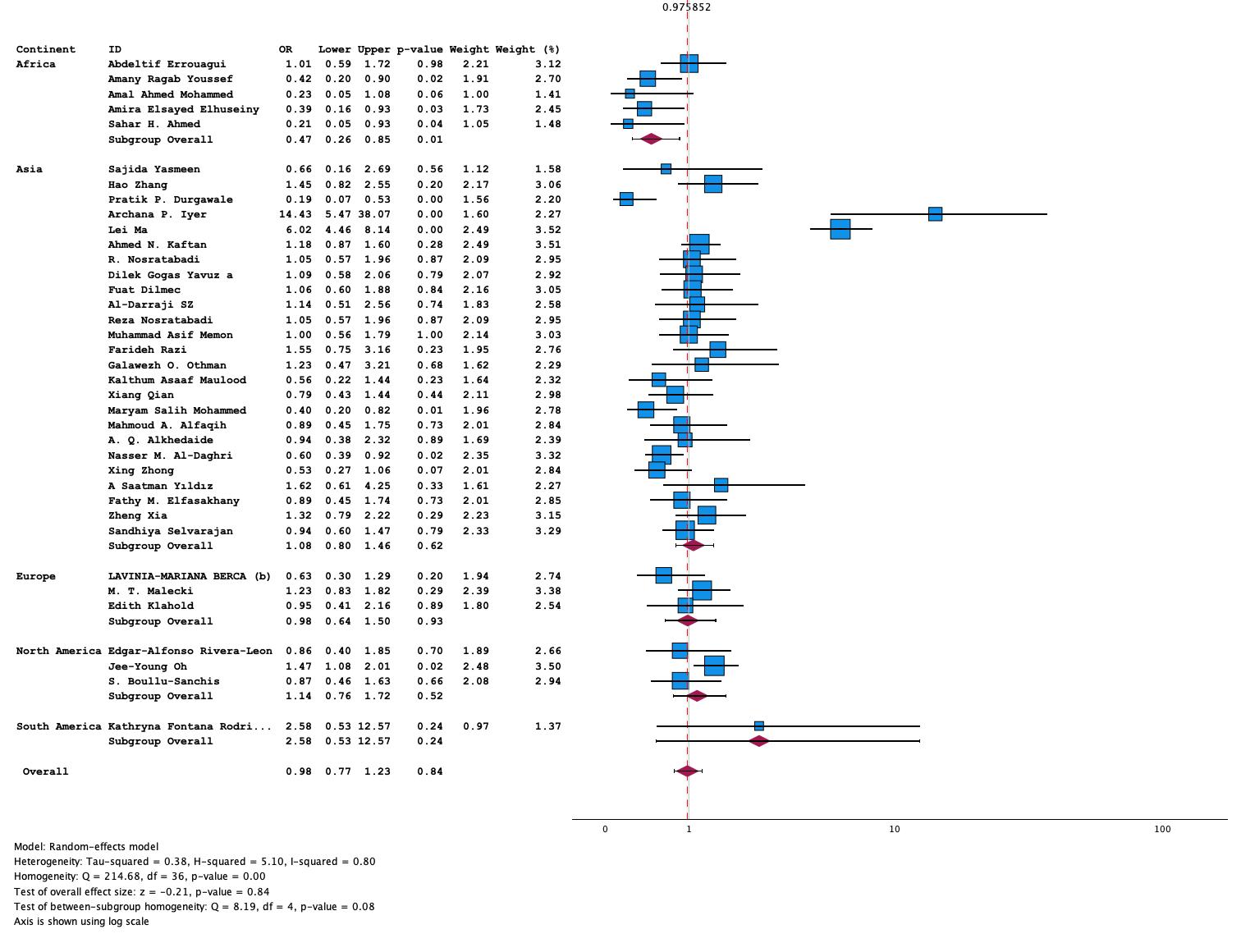

S1. Figure 20 Forest Plot of ApaI SNP Recessive (GG vs. GT+TT) model in T2DM
